# Time-varying reproduction numbers of COVID-19 in Georgia, USA, March 2-June 14, 2020

**DOI:** 10.1101/2020.05.19.20107219

**Authors:** Kamalich Muniz-Rodriguez, Gerardo Chowell, Jessica S. Schwind, Randall Ford, Sylvia K. Ofori, Chigozie A. Ogwara, Margaret R. Davies, Terrence Jacobs, Chi-Hin Cheung, Logan T. Cowan, Andrew R. Hansen, Isaac Chun-Hai Fung

## Abstract

In 2020, SARS-CoV-2 impacted Georgia, USA. Georgia announced state-wide shelter-in-place on April 2 and partially lifted restrictions on April 27. We analyzed daily incidence of confirmed COVID-19 cases by reporting date, March 2-June 14, in Georgia, Metro Atlanta, and Dougherty County and estimate the time-varying reproduction number, *R*_*t*_, using R package EpiEstim. The median *R*_*t*_ estimate in Georgia dropped from between 2 and 4 in mid-March, to <2 in late March, and around 1 from mid-April to mid-June. Regarding Metro Atlanta, *R*_*t*_ fluctuated above 1.5 in March and around 1 since April. In Dougherty County, the median *R*_*t*_ declined from around 2 in late March to 0.32 on April 26. In Spring 2020, SARS-CoV-2 transmission in Georgia declined likely because of social distancing measures. However, as restrictions were relaxed in late April, community transmission continued with *R*_*t*_ fluctuating around 1 across Georgia, Metro Atlanta, and Dougherty County as of mid-June.

## INTRODUCTION

In 2020, the pandemic of coronavirus disease 2019 (COVID-19), caused by severe acute respiratory syndrome coronavirus 2 (SARS-CoV-2), impacted the state of Georgia (GA) as in other jurisdictions within the United States. Within Georgia, Metro Atlanta counties have been the hardest hit by the virus with thousands of confirmed cases cumulatively: 15,221 in Fulton, 14,801 in Gwinnett, and 10,767 in Dekalb as of July 24, 2020 (Georgia Department of Public Health 2020). Dougherty County, with Albany as the county seat, was a COVID-19 hotspot in southeast GA and reported a large number of cases (as of July 24, 2020: cumulative number = 2,428, with incidence rate, IR=2,701 per 100,000 individuals) (Georgia Department of Public Health 2020). In GA, every county government had the power to impose preventive measures to reduce viral transmission as they see fit, before the state imposed a state-wide emergency that overrode autonomy of county governments (Table 1). On March 23, 2020, the GA State Government issued an executive order requesting citizens with underlying conditions and those with a COVID-19 diagnosis to shelter-in-place (State of Georgia Government 2020a). Certain businesses were to remain closed and no more than 10 individuals could gather in a location without maintaining a distance of at least six feet. The order also called for restaurants to offer only curbside pick-ups or deliveries (State of Georgia Government 2020a). On April 2, 2020, a state-wide shelter-in-place ordinance was enacted by the Governor allowing only essential services to operate (implemented on April 3) (State of Georgia Government 2020c). The GA State Government announced on April 27, 2020 during a press conference that services, such as beauty salons, barber shops, stores, and restaurants, can reopen if they follow pertinent social distancing measures specified by the state (State of Georgia Government 2020b). As the COVID-19 epidemic in GA continues, it is important to quantify the epidemiologic characteristics of COVID-19 so that we may formulate policies and implement interventions to minimize transmission and mortality.

**Table 1.**
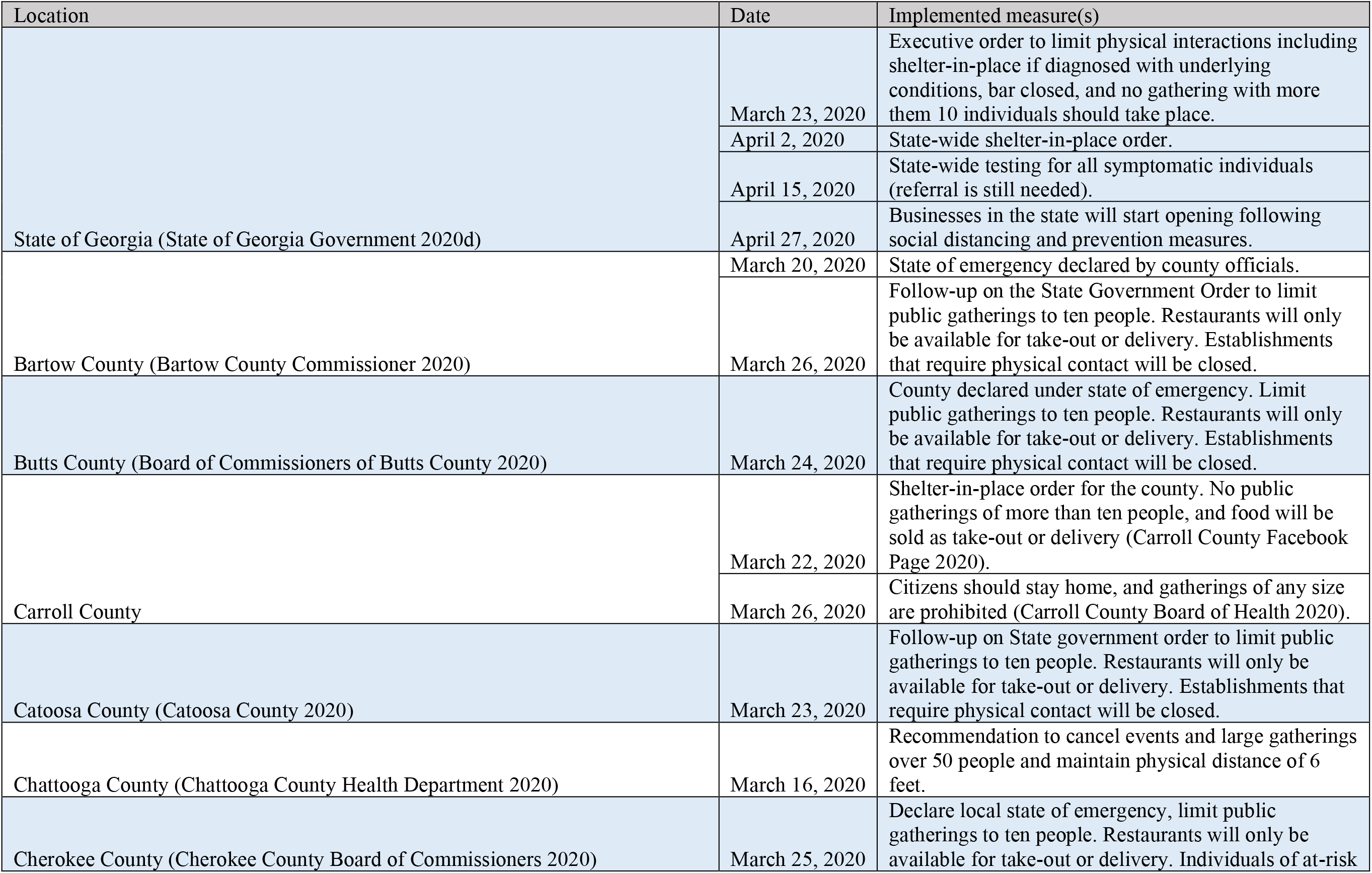
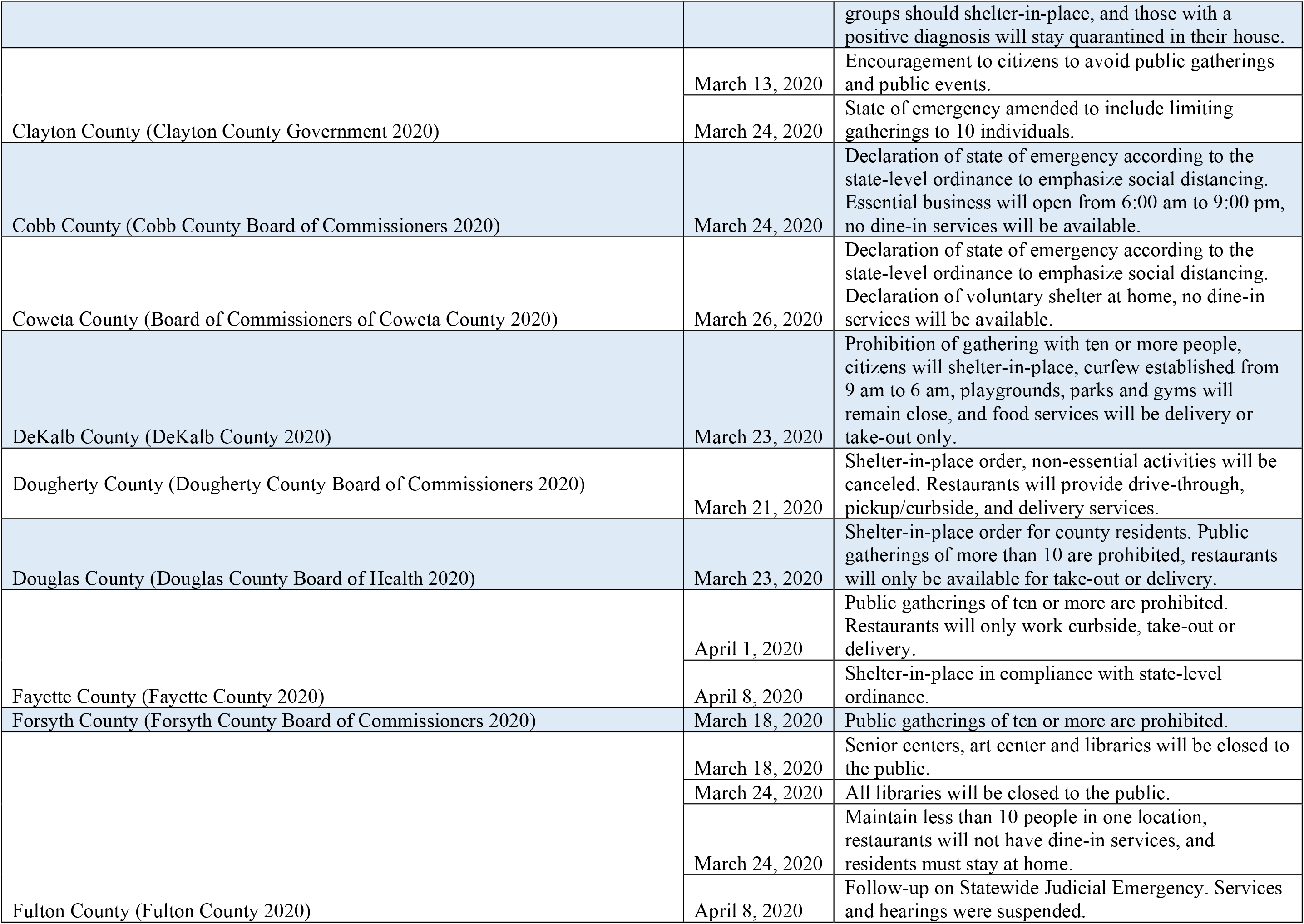
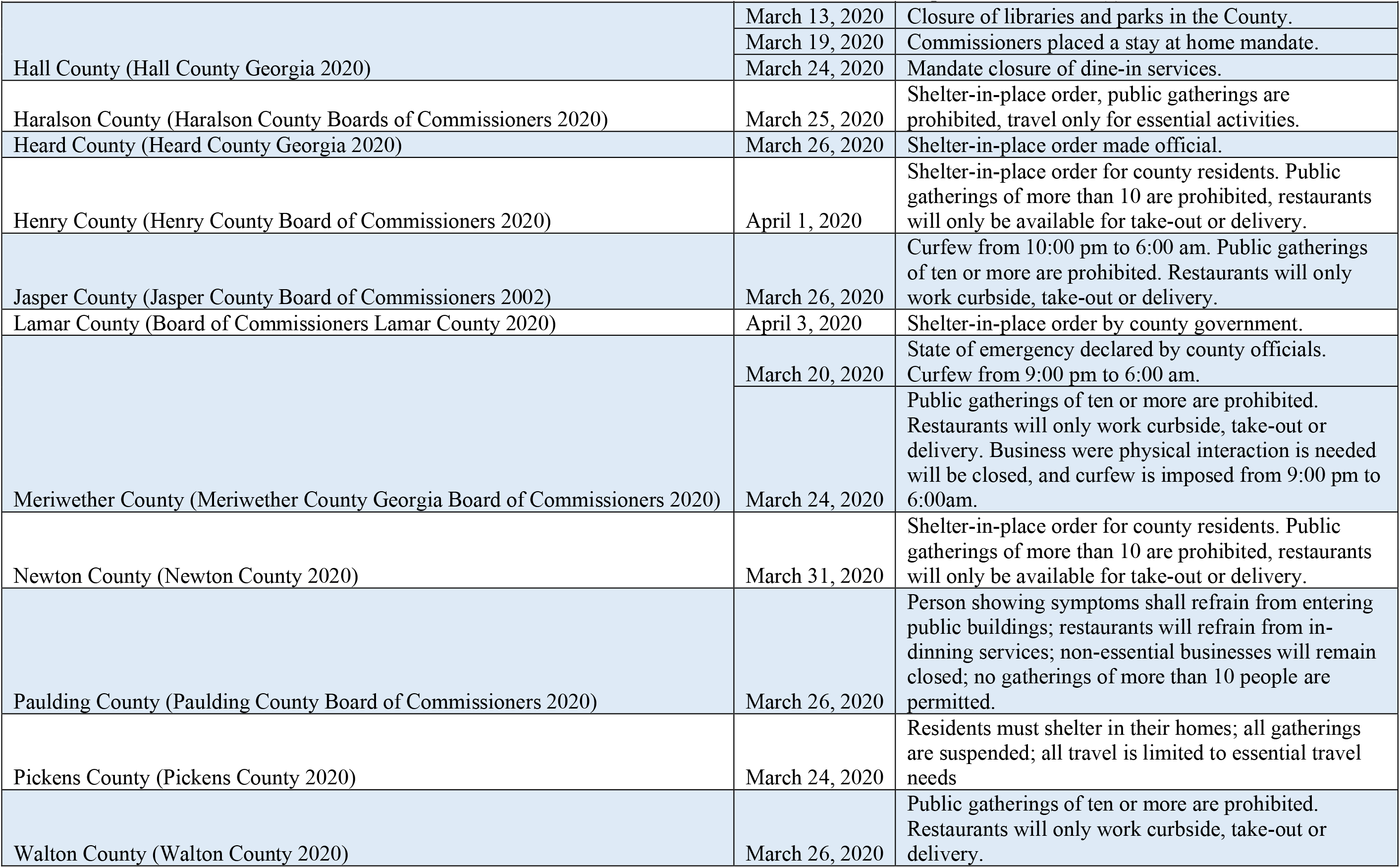
Control measures announced and implemented by state and local government agencies in the state of Georgia, in Metro Atlanta counties and Dougherty County in March-April, 2020. Note that most of these control measures began to be implemented the day after the announcement. For example, Georgia’s state-wide shelter-in-place was announced on April 2, 2020 to be implemented on April 3, 2020.

To characterize the transmission potential of an epidemic, it is necessary to calculate the reproduction number based on the trajectory of the incidence curve (Vynnycky and White 2010). The basic reproduction number, *R*_*0*_, is the average number of secondary cases that one primary case can generate in a completely susceptible population in the absence of behavioral changes or public health interventions (Vynnycky and White 2010). The estimated values of the *R*_*0*_ for SARS-CoV-2 vary across geographic locations. An early study of the epidemic in Wuhan reported a *R*_*0*_ of 2.2, assuming serial interval of 7.5 (Li et al. 2020). A more recent study of the epidemic in China, adjusted for the changing case definition, estimated *R*_*0*_ of 1.8-2.0 (assuming serial interval of 7.5) or 1.4-1.5 (assuming serial interval of 4.7) (Tsang et al. 2020). Assuming serial interval of 4.4, our analysis of confirmed COVID-19 cases in Iran estimated the mean *R*_*0*_ as 3.5 or 4.4, depending on the statistical method chosen (Muniz-Rodriguez et al. 2020).

In contrast, the time-varying reproduction number, *R*_*t*_, is a time-dependent estimate of the secondary cases that arise from one case at time *t*, when depletion of the susceptible population, behavioral changes, and measures to control transmission of disease have taken place (Cori et al. 2013; Thompson et al. 2019). As with *R*_*0*_, if *R*_*t*_>1, it indicates there is sustainable transmission in the population. When *R*_*t*_<1, disease transmission cannot be sustained, and it is used as an indication of the effectiveness of infection control measures (Vynnycky and White 2010; Thompson et al. 2019).

Various statistical methods have been proposed to estimate *R*_*t*_. Their strengths and weaknesses have been recently assessed by researchers who compared the performance of different methods using synthetic epidemic data (Gostic et al. 2020; O’Driscoll et al. 2020), and observed COVID-19 incidence data (Roosa et al. 2020). In this paper, we chose to use an oft-used method, known as the instantaneous reproduction number method, as implemented in the R package EpiEstim (Cori et al. 2013; Thompson et al. 2019). This Bayesian method provides an estimate of the average *R*_*t*_ over a short time window specified by the user (in this paper both a 7-day window and a 14-day window that end at time *t*). It treats the fluctuation in incidence data as signals of an increasing or decreasing reproduction number. This method has been used to estimate COVID-19 *R*_*t*_ in jurisdictions such as mainland China (Leung et al. 2020), Hong Kong (Cowling et al. 2020), Iran (Najafi et al. 2020), South Korea (Zhuang et al. 2020), Italy (Moirano et al. 2020), Nigeria (Adegboye et al. 2020), and Switzerland (Scire et al. 2020).

This study aimed to estimate *R*_*t*_ for COVID-19 in GA, Dougherty County, and Metro Atlanta counties, analyzing historical data from March 2 through June 14, 2020, as the state incrementally implemented and then relaxed social distancing interventions (Table 1).

## METHODS

This study uses historical data from the COVID-19 pandemic, March 2 — June 14, 2020, in the state of GA, Dougherty County, and all Metro Atlanta counties (Supplementary Materials Tables S1-S12). Metro Atlanta is defined by the United States Office of Management and Budget as the “Atlanta-Sandy Springs-Alpharetta, Georgia Metropolitan Statistical Area” (Office of Management and Budget 2015). The list of Metro Atlanta counties is provided in Table 2.

**Table 2.** Estimates for the time-varying reproduction number, *R*_*t*_, on June 14, 2020 for the state of Georgia, Dougherty County, and Metro Atlanta counties, using the instantaneous reproduction number method as implemented in the R package EpiEstim. The analysis used a serial interval following a gamma distribution with a mean of 4.60 days and a standard deviation of 5.55 days, and α = 0.05.

| Location | First reported case<br>(dd-mmm-yy) | EpiEstim (1-week window) |  | EpiEstim (2-week window) |  |
| --- | --- | --- | --- | --- | --- |
| | | Median $R_t$<br>(2.5%, 97.5% quantiles) | Mean $R_t$ (Standard<br>Deviation) | Median $R_t$<br>(2.5%, 97.5% quantiles) | Mean $R_t$ (Standard<br>Deviation) |
| Georgia | 2-Mar-20 | 1.14 (1.11, 1.17) | 1.14 (0.02) | 1.07 (1.05, 1.09) | 1.07 (0.01) |
| Dougherty | 15-Mar-20 | 1.03 (0.72, 1.41) | 1.04 (0.18) | 0.93 (0.72, 1.17) | 0.93 (0.11) |
| Metro Atlanta | 2-Mar-20 | 1.02 (0.98, 1.06) | 1.02 (0.02) | 0.98 (0.95, 1.01) | 0.98 (0.01) |
| Bartow | 11-Mar-20 | 0.73 (0.46, 1.08) | 0.74 (0.16) | 0.95 (0.74, 1.20) | 0.96 (0.12) |
| Butts | 22-Mar-20 | 0.47 (0.11, 1.27) | 0.53 (0.30) | 0.78 (0.46, 1.21) | 0.79 (0.19) |
| Carroll | 20-Mar-20 | 0.89 (0.63, 1.22) | 0.90 (0.15) | 0.96 (0.77, 1.17) | 0.96 (0.10) |
| Cherokee | 8-Mar-20 | 1.06 (0.83, 1.33) | 1.07 (0.13) | 0.87 (0.72, 1.03) | 0.87 (0.08) |
| Clayton | 15-Mar-20 | 1.06 (0.79, 1.16) | 1.07 (0.10) | 1.02 (0.89, 1.16) | 1.02 (0.07) |
| Cobb | 7-Mar-20 | 1.15 (1.03, 1.27) | 1.15 (0.06) | 1.03 (0.95, 1.12) | 1.03 (0.04) |
| Coweta | 14-Mar-20 | 1.00 (0.75, 1.31) | 1.01 (0.14) | 0.96 (0.78, 1.17) | 0.96 (0.10) |
| Dawson | 20-Mar-20 | 0.94 (0.36, 1.93) | 0.99 (0.41) | 0.86 (0.43, 1.53) | 0.89 (0.28) |
| Dekalb | 9-Mar-20 | 0.97 (0.86, 1.08) | 0.97 (0.06) | 0.89 (0.82, 0.96) | 0.89 (0.04) |
| Douglas | 20-Mar-20 | 1.10 (0.85, 1.40) | 1.11 (0.14) | 1.07 (0.88, 1.28) | 1.07 (0.10) |
| Fayette | 9-Mar-20 | 1.15 (0.68, 1.79) | 1.17 (0.28) | 1.08 (0.72, 1.54) | 1.09 (0.21) |
| Forsyth | 16-Mar-20 | 1.10 (0.82, 1.43) | 1.10 (0.16) | 1.00 (0.80, 1.23) | 1.00 (0.11) |
| Fulton | 2-Mar-20 | 0.94 (0.83, 1.05) | 0.94 (0.06) | 0.88 (0.81, 0.95) | 0.88 (0.04) |
| Gwinnett | 7-Mar-20 | 1.05 (0.97, 1.12) | 1.05 (0.04) | 1.05 (0.99, 1.10) | 1.05 (0.03) |
| Hall | 16-Mar-20 | 0.96 (0.81, 1.13) | 0.96 (0.08) | 0.99 (0.88, 1.11) | 0.99 (0.06) |
| Haralson | 26-Mar-20 | 0.90 (0.35, 1.85) | 0.95 (0.39) | 1.05 (0.60, 1.68) | 1.07 (0.28) |
| Heard | 20-Mar-20 | 1.44 (0.65, 2.71) | 1.50 (0.53) | 0.97 (0.48, 1.72) | 1.01 (0.32) |
| Henry | 13-Mar-20 | 1.16 (0.93, 1.43) | 1.17 (0.13) | 1.14 (0.97, 1.34) | 1.15 (0.09) |
| Jasper | 24-Mar-20 | 0.89 (0.42, 1.62) | 0.93 (0.31) | 1.07 (0.67, 1.60) | 1.08 (0.24) |
| Lamar | 20-Mar-20 | 0.68 (0.20, 1.63) | 0.74 (0.37) | 0.79 (0.43, 1.30) | 0.81 (0.22) |
| Meriwether | 24-Mar-20 | 1.07 (0.66, 1.61) | 1.08 (0.24) | 1.19 (0.86, 1.59) | 1.20 (0.19) |
| Morgan | 23-Mar-20 | 1.05 (0.15, 3.50) | 1.25 (0.89) | 1.10 (0.33, 2.63) | 1.20 (0.60) |
| Newton | 15-Mar-20 | 0.91 (0.61, 1.31) | 0.92 (0.18) | 0.92 (0.71, 1.17) | 0.93 (0.12) |
| Paulding | 16-Mar-20 | 1.15 (0.87, 1.49) | 1.16 (0.16) | 1.00 (0.79, 1.24) | 1.01 (0.11) |
| Pickens | 20-Mar-20 | 1.48 (0.83, 2.40) | 1.51 (0.40) | 1.29 (0.78, 1.99) | 1.32 (0.31) |
| Pike | 28-Mar-20 | 1.11 (0.52, 2.01) | 1.15 (0.38) | 1.24 (0.73, 1.96) | 1.27 (0.32) |
| Rockdale | 19-Mar-20 | 1.18 (0.82, 1.63) | 1.19 (0.21) | 1.13 (0.86, 1.45) | 1.13 (0.15) |
| Spalding | 20-Mar-20 | 1.04 (0.66, 1.56) | 1.06 (0.23) | 1.06 (0.77, 1.43) | 1.07 (0.17) |
| Walton | 24-Mar-20 | 0.81 (0.54, 1.17) | 0.82 (0.16) | 0.82 (0.63, 1.05) | 0.82 (0.11) |

Note. All data collection concluded on June 14, 2020.

### Data acquisition

We downloaded the cumulative data of confirmed cases from March 2 — June 14, 2020, for the entire state of GA and its counties from the New York Times (NYT) GitHub data repository (The New York Times 2020). Every jurisdiction in our study has a start date corresponding to the first reported case for the area, according to NYT (Table 2). The first case in GA was reported on March 2, 2020 (The New York Times 2020). Our cutoff point for all jurisdictions was June 14, 2020, 7 weeks after businesses in the state reopened on April 27, 2020, following social distancing and prevention measures (Table 1) (State of Georgia Government 2020b).

We verified the numbers with official statistical reports from the Georgia Department of Public Health (GA-DPH) (Georgia Department of Public Health 2020). If any inconsistencies were found, the numbers from the GA-DPH were used as the standard. To estimate *R*_*t*_, we transformed the cumulative numbers into daily number of cases reported to obtain daily SARS-CoV-2 incidence (Supplementary Materials Table S1-S12). We also searched the local government pages to verify if any control measures were established. Such information is presented in Table 1.

### Statistical analyses

#### The instantaneous reproduction number (EpiEstim package)

The *R*_*t*_ estimated by the R package EpiEstim is also known as an instantaneous reproduction number (Cori et al. 2013). This measure is defined by Cori et al. (2013) as the ratio between *I*_*t*_, the number of incident cases at time *t*, to *Λ*_*t*_, the total infectiousness of all the infected individuals at time *t*.

The latter is represented mathematically as 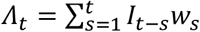, i.e., the sum of infection incidence up to time *t-1*, weighted by an infectivity function *w*_*s*_, which is a probability distribution describing the average infectiousness profile after infection and is usually represented by the serial interval distribution. Thus, the number of new cases at time *t* is Poisson-distributed with a mean of 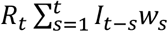. Conditional on the number of new cases in previous time points, *I*_*0*_, …, *I*_*t-1*_, and given the reproduction number *R*_*t*_, the likelihood of the incidence *I*_*t*_ is as follows: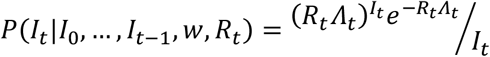 where the total infectiousness of infected individuals at time *t*,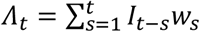.(Cori et al. 2013)

In other words,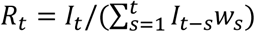.This can be interpreted as the average number of secondary cases that an infectious individual will infect at time *t*, if conditions remain the same at time *t*. As this formulation may result in a highly variable *R*_*t*_ estimate over time, the instantaneous reproduction number method as implemented in EpiEstim assumes that the *R*_*t*_ is constant over a specified time window of size *τ* ending at time *t*. This will allow the estimate to be less variable and more precise. Given that transmissibility is assumed constant over a time period, from (*t*-*τ*+1) to *t*, and is denoted by a reproduction number, *R*_*t,τ*_, the likelihood of the incidence during the time period, from *I*_*(t-τ+1)*_ to *I*_*t*_, conditional on incidence prior to the time period, from *I*_*0*_, to *I*_*(t-τ)*_, is as follows (Cori et al. 2013):

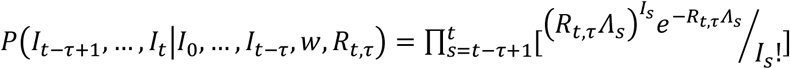

Here, we used a time window of 7 days (main analysis) and that of 14 days (sensitivity analysis) respectively.

Using a Bayesian framework with a Gamma-distributed prior for *R*_*t,τ*_, Cori et al. derived an analytical expression of the posterior distribution of *R*_*t*_ and thus estimated its median, the variance, and the 95% credible interval (Cori et al. 2013). In this paper, the data was analyzed using EpiEstim version 2.2-3 (Cori et al. 2013; Thompson et al. 2019) implemented in R Version 1.2.1335 Macintosh (R Core Team, Vienna, Austria).

#### Ethics

The Georgia Southern University Institutional Review Board made a non–human subjects determination for this project (H20364), under the G8 exemption category.

## RESULTS

Community transmission of SARS-CoV-2 remained ongoing in GA based on incidence data by date of report from March 2 – June 14, 2020. As of June 14, 2020, the median EpiEstim *R*_*t*_ estimates were 1.14 (95% credible interval, CrI, 1.11, 1.17) for a 1-week-window and 1.07 (95% CrI, 1.05, 1.09) for a 2-week-window respectively. In Metro Atlanta, the transmission may have been under control, with *R*_*t*_ estimates being 1.02 (95% CrI, 0.98, 1.06) for a 1-week-window and 0.98 (95% CrI, 0.95, 1.01) for a 2-week-window. Likewise, in Dougherty County, the transmission with *R*_*t*_ estimates being 1.03 (95% CrI, 0.72, 1.41) for a 1-week-window and 0.93 (95% CrI, 0.72, 1.17) for a 2-week-window (Table 2).

The EpiEstim *R*_*t*_ estimates were sensitive to fluctuation in incident cases. As social distancing measures unfolded and then relaxed in GA during our study period (March 2 – June 14, 2020), the median EpiEstim *R*_*t*_ estimate (1-week window) in Georgia dropped from between 2 and 4 in mid-March, to <2 in late March and early April. The median *R*_*t*_ dropped <1 for the first time on April 15. *R*_*t*_ fluctuated around 1 from mid-April to June 14. The 2-week window EpiEstim *R*_*t*_ estimate largely followed the trend of the 1-week window estimate, even though the 2-week estimate was smoother than the 1-week estimate when the *R*_*t*_ estimate fluctuated around 1 from late April to mid-June (Figure 1).

**Figure 1.**
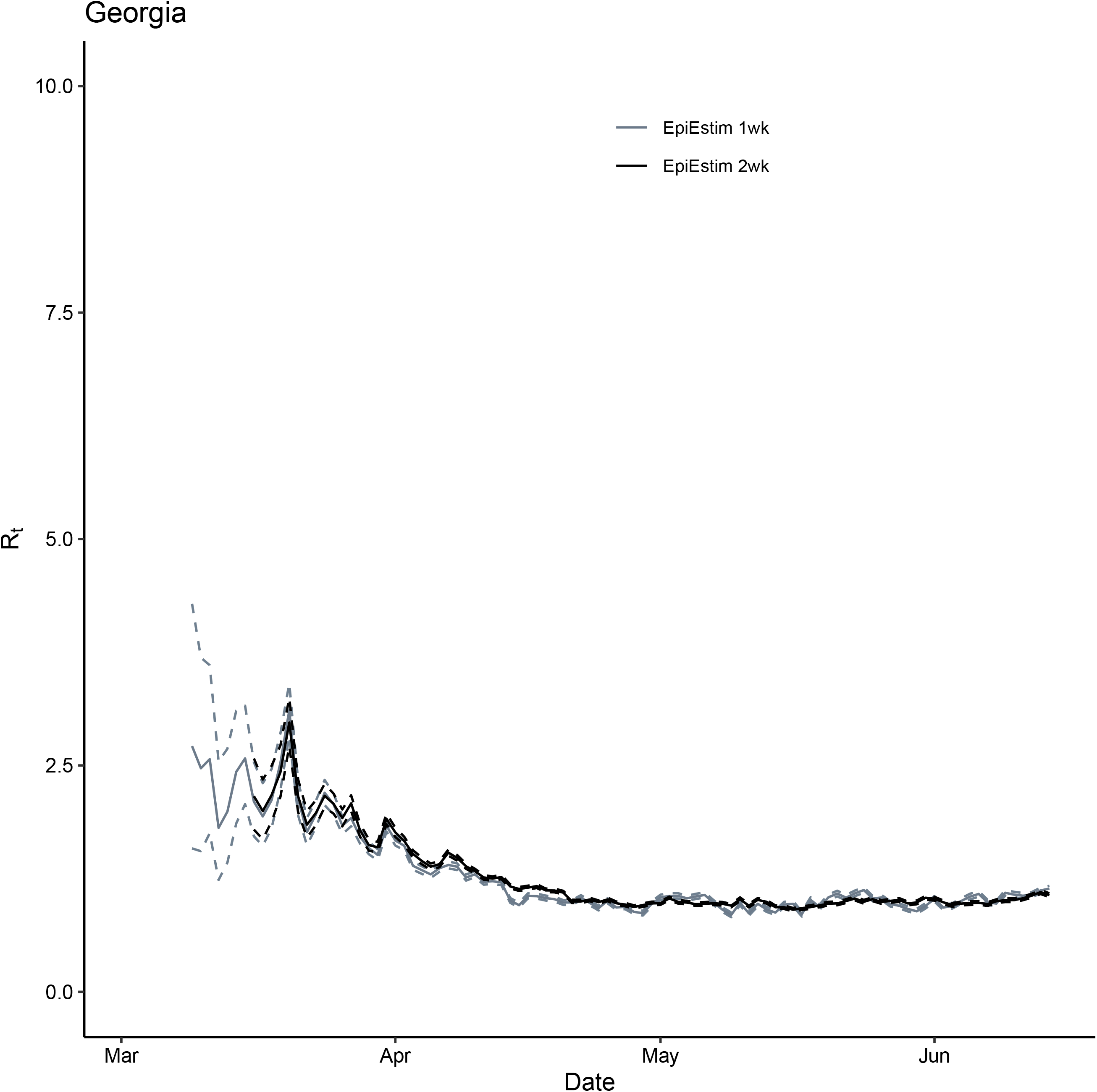
Comparison between *R*_*t*_ for Georgia, USA, March 2—June 14, 2020, estimated using the instantaneous reproduction number method implemented in EpiEstim package (grey: 1-week window and black: 2-week window). Solid lines represent the median estimates and the dashed line represent the 2.5 and 97.5 quantiles.

Regarding Metro Atlanta (Figure 2), EpiEstim *R*_*t*_ estimates (both 1-week and 2-week window) fluctuated above 1.5 in March and gradually decreased to around 1 in April and continued to fluctuate above and below 1. The *R*_*t*_ estimates for each of the Metro Atlanta counties are presented in Figures S1 to S29 in the Supplementary Materials.

**Figure 2.**
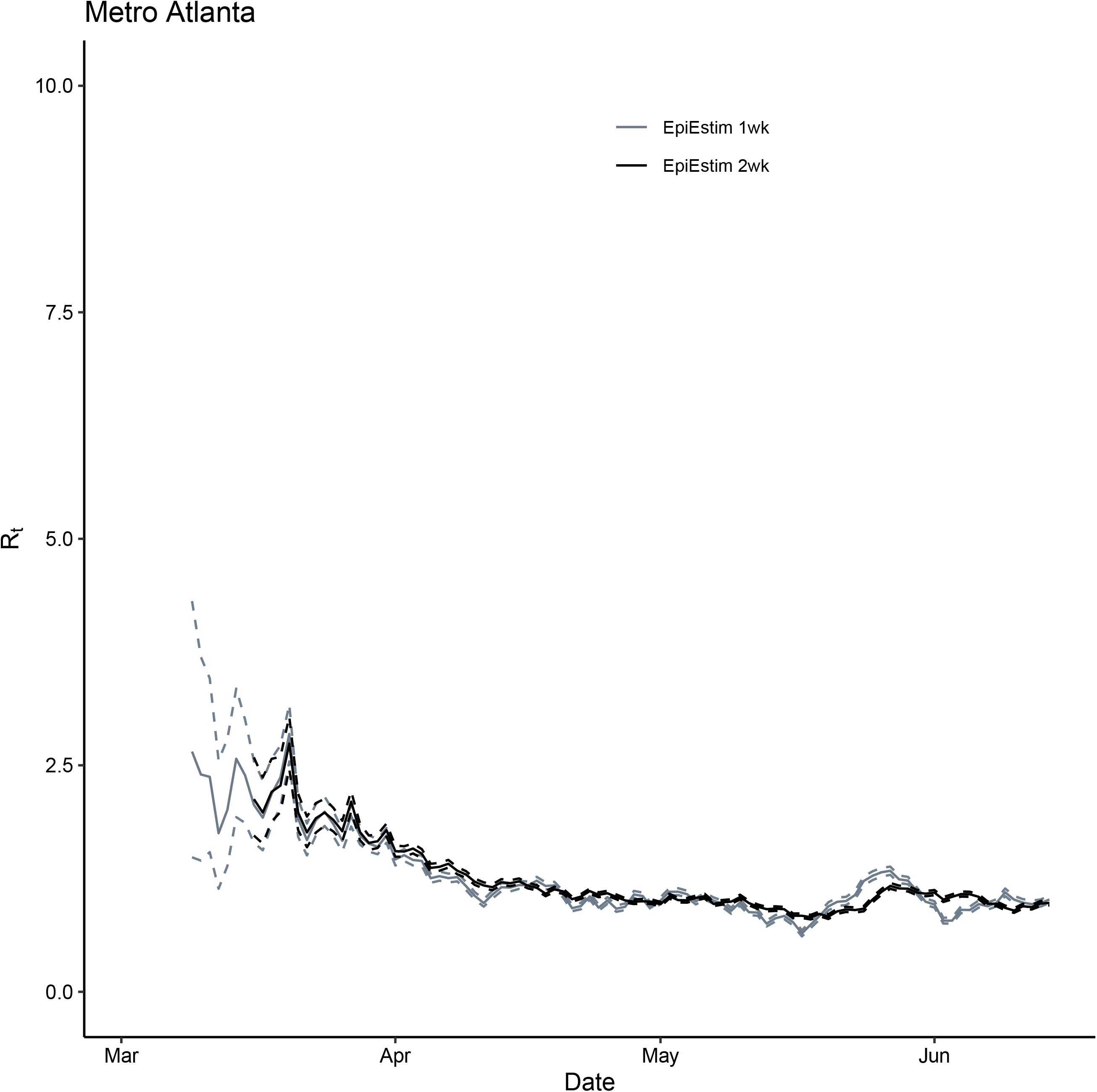
Comparison between *R*_*t*_ for Metro Atlanta, GA, March 2—June 14, 2020, estimated using the instantaneous reproduction number method implemented in EpiEstim package (grey: 1-week window and black: 2-week window). Solid lines represent the median estimates and the dashed line represent the 2.5 and 97.5 quantiles.

Regarding Dougherty County (Figure 3), we observed a speedy decline in EpiEstim *R*_*t*_ estimates (both 1-week and 2-week window) from around 2 in late March to a low of 0.32 on April 26. This finding was primarily driven by the early epidemic observed in Dougherty County, which were large clusters of cases infected via two funerals that happened to be superspreading events (Barry 2020).

**Figure 3.**
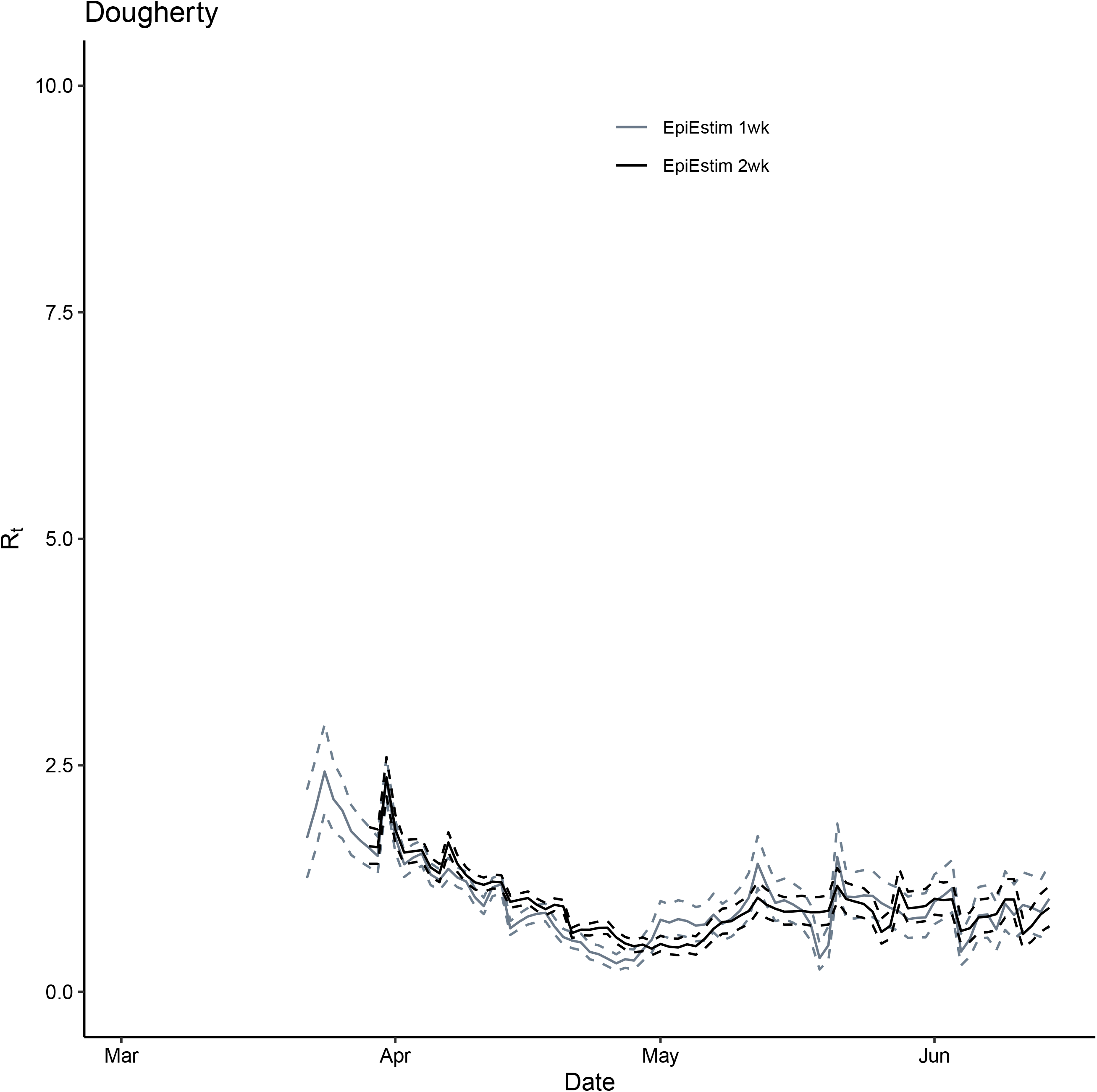
Comparison between *R*_*t*_ for Dougherty, GA, March 2—June 14, 2020, estimated using the instantaneous reproduction number method implemented in EpiEstim package (grey: 1-week window and black: 2-week window). Solid lines represent the median estimates and the dashed line represent the 2.5 and 97.5 quantiles.

## DISCUSSION

Community transmission of SARS-CoV-2 remained ongoing in GA as of June 14, 2020 (i.e., 101/2 weeks after statewide shelter-in-place order was first imposed on April 2 and 7 weeks after businesses reopened under new social distancing guidance) (Table 1). The 95% CrI of both the 1-week and 2-week window EpiEstim *R*_*t*_ estimates were >1.

On April 27, GA reopened some sectors of the economy, with specific guidelines pertinent to social distancing (State of Georgia Government 2020b). As the economy reopened and increased unprotected social mixing ensued, an increase in daily number of new confirmed cases was observed in June as SARS-CoV-2 transmission continued unabated (Georgia Department of Public Health 2020). Our study documented the decrease in *R*_*t*_ following social distancing interventions in GA and provided further evidence that social distancing measures remained important to keep COVID-19 under control.

Furthermore, many residents in both rural and urban GA are medically vulnerable. A recent analysis by The Surgo Foundation estimated the COVID-19 community vulnerability index (CCVI) for Dougherty County, by combining epidemiologic risk factors for infection and sociodemographic factors, at very high levels (CCVI = 0.87) when compared with counties in Metro Atlanta (Fulton county’s CCVI = 0.42) (Surgo Foundation 2020). Although the EpiEstim *R*_*t*_ estimates for Dougherty County were <1 from April 14 to April 30, *R*_*t*_ rebounded since late April, and it has been fluctuating with 95% CrI including 1 most of the time from early May to mid-June (Figure 3). This corresponded to the partial reopening of the GA economy. The relaxation of social distancing measures should be implemented with an abundance of caution due to the population’s vulnerability. Another important factor for consideration is access to healthcare and surge capacity in hospitals, especially in rural GA. The healthcare system in Dougherty County was heavily impacted by the surge of COVID-19 patients driven by superspreading events (Barry 2020).

While economic factors are legitimate concerns for decisionmakers, reopening the economy too soon led to the resurgence of COVID-19 cases in GA as observed in June. Further research into the spatiotemporal variation of SARS-CoV-2 transmission potential and its association with economic and medical vulnerability will shed light on the disease and economic burden of COVID-19 in GA.

Our study estimated *R*_*t*_ using the instantaneous reproduction number method implemented in the R package EpiEstim (Cori et al. 2013; Thompson et al. 2019). The EpiEstim estimate is sensitive to fluctuation in daily incident case counts as the instantaneous reproduction number method treats such changes as meaningful signals reflecting genuine increases or decreases in transmission potential. The instantaneous reproduction number method in the EpiEstim package can be used if the purpose is to identify time-dependent changes in the *R*_*t*_ estimate that reflects the implementation or relaxation of social distancing measures over time.

Regarding the time window chosen for EpiEstim, we used a window of 7 days in our main analysis and 14 days in our sensitivity analysis. We did not use a window of <7 days, because a weekend effect was observed in the data, i.e., the daily number of cases reported in the weekend was consistently smaller than those in the weekdays before or after the weekend. The 2-week window EpiEstim estimate lagged behind the 1-week window EpiEstim estimate when the *R*_*t*_ estimate fluctuates around 1. This result can be explained because the 2-week estimate was an average of 14 days and did not reflect the immediate change in transmission potential.

### Limitations

Our study is limited by several factors. First, we used the NYT dataset; data was recorded by reported date and not by day of symptom onset. Given the time difference between the transmission events and subsequent symptom onset, testing, and case reporting, the *R*_*t*_ estimates should reflect transmission that had happened several days ago (the sum of the incubation period and the delay to testing and case reporting). Second, our data does not differentiate between imported and community transmission cases. Third, cases may be underreported due to limited testing capacity, or they may be mild or asymptomatic cases. Fourth, our analysis is right-censored by June 14, 2020. Future studies can further extend the analysis as the pandemic progresses. Fifth, in addition to the two methods utilized here, there are other statistical methods that estimate *R*_*t*_ (Gostic et al. 2020; O’Driscoll et al. 2020), e.g., the case reproduction number method as proposed by Walling and Teunis (2004) but they are out-of-scope for this paper.

## CONCLUSION

In Spring 2020, social distancing measures reduced the transmission of SARS-CoV-2 in GA. However, the epidemic has not yet been suppressed. *R*_*t*_ was fluctuating around 1 for GA, Metro Atlanta and Dougherty County since GA economy reopened in late April 2020. Government agencies should carefully consider the next steps of their COVID-19 plans for their communities considering ongoing transmission across GA.

## Data Availability

All data used in the analysis is presented in the Supplementary Materials.

## Acknowledgment

G.C. received support from NSF grant 1414374 as part of the joint NSF-NIH-USDA Ecology and Evolution of Infectious Diseases program. I.C.-H.F. received salary support from the Centers for Disease Control and Prevention (CDC) (19IPA1908208). This article is not part of I.C.-H.F’s CDC-sponsored projects. We would like to acknowledge Bryan O. Sepulveda-Bahamundi, M.S. for his contribution in data collection for this project.

## Online Supplementary Materials

**Table S1.**
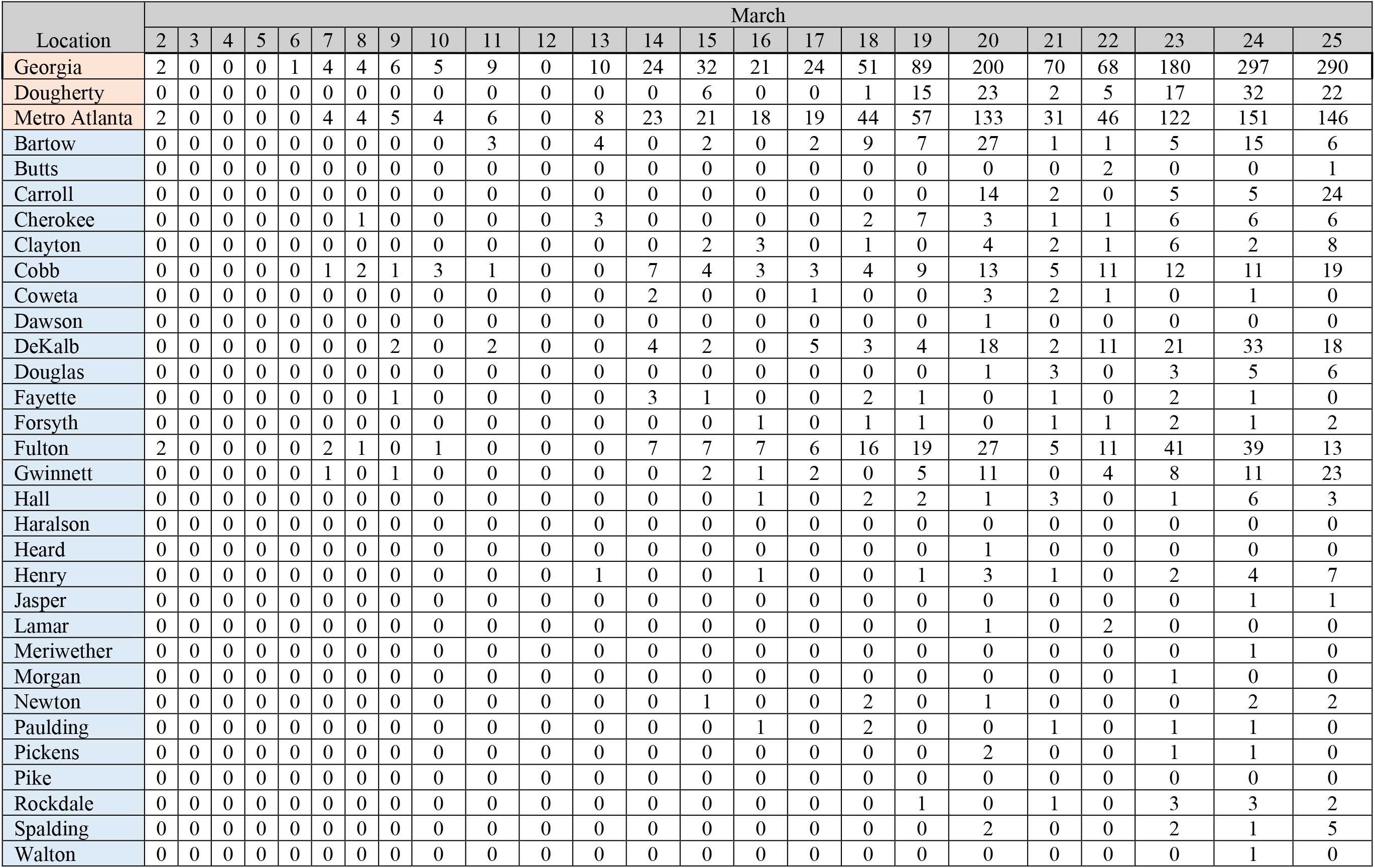
Daily number of COVID-19 cases reported in the state of Georgia, Dougherty County and Metro Atlanta counties, March 2—25, 2020.

**Table S2.** Daily number of COVID-19 cases reported in the state of Georgia, Dougherty County and Metro Atlanta counties, from March 26— April 18, 2020.

| Location | March |  |  |  |  |  | April |  |  |  |  |  |  |  |  |  |  |  |  |  |  |  |  |  |
| --- | --- | --- | --- | --- | --- | --- | --- | --- | --- | --- | --- | --- | --- | --- | --- | --- | --- | --- | --- | --- | --- | --- | --- | --- |
|  | 26 | 27 | 28 | 29 | 30 | 31 | 1 | 2 | 3 | 4 | 5 | 6 | 7 | 8 | 9 | 10 | 11 | 12 | 13 | 14 | 15 | 16 | 17 | 18 |
| Georgia | 256 | 555 | 249 | 236 | 349 | 1085 | 631 | 696 | 523 | 416 | 359 | 816 | 1598 | 1048 | 681 | 974 | 402 | 191 | 1169 | 292 | 670 | 1061 | 1014 | 1183 |
| Dougherty | 41 | 39 | 21 | 23 | 31 | 188 | 24 | 31 | 86 | 78 | 3 | 34 | 251 | 28 | 41 | 30 | 30 | 76 | 67 | 52 | 23 | 38 | 27 | 24 |
| Metro Atlanta | 125 | 397 | 141 | 182 | 283 | 483 | 154 | 370 | 556 | 355 | 84 | 430 | 576 | 314 | 302 | 380 | 167 | 241 | 642 | 674 | 483 | 427 | 642 | 191 |
| Bartow | 11 | 14 | 10 | 2 | 6 | 12 | 10 | 6 | 6 | 1 | 0 | 22 | 9 | 4 | 8 | 5 | 3 | 2 | 10 | 7 | 7 | 3 | 5 | 1 |
| Butts | 0 | 0 | 2 | 0 | 1 | 2 | 0 | 1 | 0 | 1 | 2 | 5 | 1 | 5 | 2 | 2 | 1 | 1 | 1 | 5 | 1 | 2 | 2 | 1 |
| Carroll | 2 | 9 | 3 | 6 | 27 | 26 | 10 | 6 | 8 | 11 | 0 | 5 | 20 | 1 | 11 | 5 | 4 | 6 | 19 | 6 | 7 | 16 | 3 | 10 |
| Cherokee | 8 | 6 | 4 | 6 | 9 | 9 | 7 | 9 | 20 | 6 | 4 | 17 | 6 | 8 | 12 | 10 | 9 | 1 | 17 | 15 | 13 | 13 | 29 | 11 |
| Clayton | 8 | 16 | 4 | 2 | 3 | 50 | 16 | 37 | 41 | 29 | 3 | 16 | 24 | 11 | 9 | 30 | 11 | 33 | 24 | 39 | 27 | 11 | 26 | 6 |
| Cobb | 10 | 44 | 22 | 43 | 22 | 37 | 17 | 37 | 81 | 31 | 21 | 43 | 49 | 42 | 45 | 28 | 24 | 23 | 88 | 79 | 55 | 64 | 71 | 19 |
| Coweta | 4 | 5 | 1 | 3 | 14 | 4 | 1 | 6 | 8 | 8 | 3 | 9 | 4 | 2 | 5 | 13 | 1 | 5 | 15 | 14 | 3 | 4 | 11 | 7 |
| Dawson | 2 | 0 | 0 | 1 | 3 | 3 | 1 | 0 | 2 | 3 | 0 | 2 | 2 | 3 | 1 | 1 | 0 | 1 | 6 | 4 | 1 | 2 | 3 | 1 |
| DeKalb | 12 | 82 | 27 | 27 | 21 | 66 | 0 | 49 | 74 | 56 | 10 | 51 | 73 | 59 | 34 | 60 | 22 | 45 | 113 | 138 | 81 | 35 | 106 | 42 |
| Douglas | 0 | 14 | 3 | 3 | 5 | 8 | 5 | 10 | 12 | 6 | 7 | 14 | 12 | 7 | 10 | 14 | 3 | 8 | 18 | 12 | 12 | 8 | 13 | 7 |
| Fayette | 2 | 8 | 4 | 1 | 15 | 6 | 0 | 7 | 6 | 5 | 1 | 7 | 7 | 9 | 3 | 0 | 6 | 1 | 6 | 9 | 7 | 4 | 8 | 2 |
| Forsyth | 5 | 6 | 1 | 6 | 8 | 11 | 3 | 3 | 8 | 10 | 2 | 12 | 14 | 0 | 10 | 10 | 9 | 5 | 10 | 10 | 4 | 12 | 16 | 12 |
| Fulton | 27 | 116 | 31 | 47 | 78 | 96 | 39 | 109 | 163 | 49 | 11 | 83 | 132 | 76 | 75 | 81 | 29 | 49 | 140 | 177 | 90 | 43 | 92 | 28 |
| Gwinnett | 10 | 42 | 10 | 14 | 33 | 64 | 15 | 46 | 50 | 47 | 10 | 45 | 85 | 46 | 32 | 51 | 12 | 20 | 65 | 49 | 70 | 32 | 120 | 22 |
| Hall | 3 | 8 | 1 | 2 | 1 | 33 | 4 | 1 | 9 | 36 | 0 | 21 | 77 | 18 | 14 | 26 | 3 | 6 | 37 | 44 | 39 | 124 | 79 | 2 |
| Haralson | 1 | 0 | 1 | 0 | 2 | 1 | 1 | 0 | 4 | 0 | 0 | 4 | 0 | 1 | 0 | 0 | 1 | 0 | 2 | 1 | 0 | 0 | 1 | 0 |
| Heard | 0 | 0 | 0 | 0 | 0 | 0 | 1 | 0 | 0 | 0 | 0 | 0 | 1 | 0 | 0 | 0 | 1 | 0 | 0 | 1 | 0 | 0 | 0 | 2 |
| Henry | 9 | 15 | 6 | 6 | 12 | 18 | 9 | 20 | 28 | 24 | 1 | 13 | 27 | 11 | 14 | 16 | 5 | 9 | 26 | 17 | 22 | 2 | 20 | 5 |
| Jasper | 0 | 0 | 0 | 0 | 0 | 0 | 0 | 0 | 0 | 2 | 0 | 0 | 2 | 0 | 0 | 0 | 0 | 2 | 1 | 0 | 1 | 0 | 4 | 1 |
| Lamar | 0 | 0 | 0 | 0 | 0 | 0 | 1 | 2 | 0 | 1 | 0 | 8 | 0 | 0 | 0 | 0 | 0 | 2 | 1 | 2 | 1 | 0 | 1 | 2 |
| Meriwether | 2 | 0 | 0 | 2 | 1 | 0 | 1 | 2 | 1 | 1 | 0 | 4 | 3 | 0 | 0 | 1 | 4 | 0 | 0 | 1 | 4 | 7 | 5 | 0 |
| Morgan | 0 | 0 | 0 | 0 | 0 | 1 | 0 | 0 | 0 | 0 | 0 | 4 | 4 | 2 | 1 | 1 | 0 | 0 | 1 | 0 | 2 | 2 | 0 | 1 |
| Newton | 4 | 3 | 0 | 3 | 4 | 9 | 3 | 3 | 5 | 10 | 3 | 10 | 2 | 0 | 3 | 7 | 1 | 4 | 11 | 7 | 13 | 2 | 6 | 0 |
| Paulding | 1 | 6 | 7 | 3 | 3 | 5 | 2 | 2 | 11 | 4 | 1 | 6 | 5 | 4 | 3 | 5 | 5 | 5 | 16 | 15 | 3 | 5 | 9 | 1 |
| Pickens | 0 | 0 | 1 | 0 | 1 | 1 | 0 | 2 | 0 | 0 | 0 | 0 | 1 | 0 | 0 | 0 | 1 | 0 | 0 | 2 | 1 | 1 | 1 | 2 |
| Pike | 0 | 0 | 1 | 1 | 0 | 0 | 0 | 0 | 2 | 1 | 0 | 4 | 1 | 1 | 1 | 3 | 1 | 0 | 3 | 3 | 3 | 3 | 1 | 1 |
| Rockdale | 3 | 3 | 1 | 1 | 11 | 16 | 2 | 10 | 10 | 8 | 2 | 5 | 3 | 0 | 0 | 4 | 3 | 5 | 4 | 5 | 1 | 4 | 6 | 2 |
| Spalding | 1 | 0 | 0 | 1 | 2 | 1 | 2 | 0 | 6 | 3 | 0 | 18 | 10 | 2 | 4 | 5 | 4 | 4 | 5 | 5 | 13 | 26 | 2 | 2 |
| Walton | 0 | 0 | 1 | 2 | 1 | 4 | 4 | 2 | 1 | 2 | 3 | 2 | 2 | 2 | 5 | 2 | 4 | 4 | 3 | 7 | 2 | 2 | 2 | 1 |

**Table S3.** Daily number of COVID-19 cases reported in the state of Georgia, Dougherty County and Metro Atlanta counties, April 19—May 2, 2020.

| Location | April |  |  |  |  |  |  |  |  |  |  |  | May |  |
| --- | --- | --- | --- | --- | --- | --- | --- | --- | --- | --- | --- | --- | --- | --- |
|  | 19 | 20 | 21 | 22 | 23 | 24 | 25 | 26 | 27 | 28 | 29 | 30 | 1 | 2 |
| Georgia | 605 | 828 | 742 | 910 | 806 | 670 | 650 | 234 | 770 | 378 | 693 | 1131 | 1005 | 832 |
| Dougherty | 16 | 11 | 20 | 0 | 0 | 9 | 5 | 0 | 10 | 13 | 7 | 6 | 25 | 3 |
| Metro Atlanta | 384 | 470 | 246 | 432 | 668 | 322 | 349 | 121 | 469 | 560 | 473 | 368 | 466 | 628 |
| Bartow | 4 | 5 | 1 | 3 | 5 | 9 | 10 | 3 | 3 | 17 | 3 | 6 | 8 | 1 |
| Butts | 9 | 30 | 1 | 44 | 0 | 0 | 0 | 1 | 0 | 4 | 4 | 11 | 6 | 0 |
| Carroll | 11 | 23 | 8 | 0 | 10 | 6 | 5 | 1 | 3 | 13 | 6 | 10 | 17 | 8 |
| Cherokee | 15 | 17 | 7 | 16 | 25 | 2 | 1 | 6 | 19 | 43 | 21 | 10 | 7 | 8 |
| Clayton | 15 | 20 | 7 | 2 | 67 | 13 | 12 | 0 | 8 | 23 | 21 | 23 | 17 | 44 |
| Cobb | 44 | 48 | 34 | 42 | 54 | 42 | 27 | 33 | 55 | 56 | 42 | 40 | 58 | 70 |
| Coweta | 4 | 5 | 0 | 0 | 8 | 10 | 0 | 1 | 2 | 1 | 4 | 0 | 16 | 2 |
| Dawson | 1 | 1 | 0 | 4 | 1 | 1 | 0 | 0 | 2 | 5 | 2 | 1 | 0 | 5 |
| DeKalb | 65 | 47 | 43 | 46 | 80 | 32 | 67 | 12 | 55 | 52 | 73 | 47 | 46 | 75 |
| Douglas | 4 | 10 | 3 | 13 | 14 | 3 | 0 | 2 | 6 | 16 | 11 | 8 | 10 | 6 |
| Fayette | 3 | 4 | 1 | 2 | 10 | 2 | 3 | 0 | 8 | 2 | 5 | 0 | 2 | 3 |
| Forsyth | 5 | 9 | 5 | 11 | 8 | 6 | 8 | 3 | 19 | 13 | 8 | 7 | 10 | 22 |
| Fulton | 66 | 67 | 8 | 49 | 181 | 64 | 43 | 2 | 135 | 73 | 32 | 36 | 59 | 48 |
| Gwinnett | 65 | 57 | 57 | 35 | 78 | 31 | 78 | 44 | 41 | 75 | 124 | 49 | 55 | 86 |
| Hall | 18 | 77 | 54 | 151 | 56 | 59 | 10 | 1 | 65 | 87 | 76 | 85 | 136 | 212 |
| Haralson | 0 | 2 | 3 | 0 | 1 | 0 | 0 | 0 | 1 | 1 | 0 | 0 | 1 | 0 |
| Heard | 0 | 0 | 1 | 0 | 1 | 0 | 0 | 0 | 1 | 0 | 0 | 0 | 1 | 0 |
| Henry | 16 | 11 | 2 | 2 | 27 | 6 | 17 | 15 | 13 | 24 | 4 | 0 | 0 | 20 |
| Jasper | 2 | 0 | 1 | 0 | 1 | 1 | 0 | 0 | 0 | 2 | 0 | 0 | 0 | 1 |
| Lamar | 1 | 2 | 2 | 0 | 2 | 0 | 2 | 0 | 1 | 1 | 2 | 1 | 1 | 1 |
| Meriwether | 2 | 4 | 1 | 1 | 1 | 0 | 0 | 0 | 0 | 4 | 1 | 0 | 0 | 0 |
| Morgan | 2 | 0 | 1 | 0 | 2 | 0 | 0 | 0 | 0 | 0 | 2 | 0 | 0 | 0 |
| Newton | 9 | 9 | 3 | 4 | 7 | 5 | 8 | 0 | 7 | 11 | 9 | 3 | 4 | 5 |
| Paulding | 4 | 4 | 0 | 4 | 3 | 8 | 3 | 1 | 6 | 6 | 4 | 13 | 1 | 4 |
| Pickens | 0 | 0 | 0 | 0 | 1 | 1 | 0 | 1 | 0 | 5 | 0 | 0 | 0 | 1 |
| Pike | 3 | 0 | 1 | 1 | 3 | 0 | 1 | 0 | 0 | 0 | 0 | 1 | 0 | 0 |
| Rockdale | 12 | 9 | 1 | 3 | 7 | 7 | 3 | 0 | 5 | 9 | 3 | 11 | 2 | 3 |
| Spalding | 2 | 7 | 1 | 0 | 4 | 4 | 45 | 1 | 7 | 4 | 4 | 5 | 4 | 1 |
| Walton | 2 | 2 | 0 | 10 | 8 | 11 | 5 | 0 | 3 | 13 | 8 | 5 | 3 | 3 |

**Table S4.** Daily number of COVID-19 cases reported in the state of Georgia, Dougherty County and Metro Atlanta counties, May 3—26, 2020.

| Location | May |  |  |  |  |  |  |  |  |  |  |  |  |  |  |  |  |  |  |  |  |  |  |  |
| --- | --- | --- | --- | --- | --- | --- | --- | --- | --- | --- | --- | --- | --- | --- | --- | --- | --- | --- | --- | --- | --- | --- | --- | --- |
|  | 3 | 4 | 5 | 6 | 7 | 8 | 9 | 10 | 11 | 12 | 13 | 14 | 15 | 16 | 17 | 18 | 19 | 20 | 21 | 22 | 23 | 24 | 25 | 26 |
| Georgia | 350 | 732 | 526 | 848 | 800 | 565 | 392 | 786 | 181 | 863 | 555 | 556 | 820 | 413 | 299 | 680 | 580 | 948 | 807 | 765 | 674 | 581 | 425 | 652 |
| Dougherty | 2 | 7 | 7 | 5 | 11 | 17 | 4 | 6 | 16 | 34 | 1 | 0 | 18 | 0 | 0 | 2 | 4 | 4 | 44 | 7 | 2 | 2 | 3 | 0 |
| Metro Atlanta | 236 | 478 | 384 | 620 | 264 | 349 | 359 | 407 | 162 | 282 | 207 | 267 | 316 | 131 | 38 | 224 | 378 | 365 | 372 | 363 | 277 | 400 | 521 | 695 |
| Bartow | 1 | 16 | 10 | 8 | 0 | 2 | 0 | 2 | 1 | 4 | 9 | 5 | 3 | 0 | 0 | 11 | 14 | 9 | 5 | 3 | 17 | 0 | 0 | 8 |
| Butts | 0 | 0 | 5 | 5 | 4 | 4 | 1 | 8 | 9 | 2 | 1 | 4 | 0 | 1 | 0 | 2 | 0 | 6 | 0 | 1 | 2 | 1 | 2 | 3 |
| Carroll | 0 | 4 | 1 | 2 | 0 | 1 | 3 | 2 | 0 | 2 | 0 | 3 | 8 | 0 | 0 | 0 | 10 | 7 | 12 | 2 | 1 | 5 | 27 | 18 |
| Cherokee | 9 | 18 | 10 | 25 | 25 | 9 | 15 | 16 | 12 | 14 | 20 | 21 | 19 | 5 | 14 | 9 | 22 | 13 | 11 | 21 | 17 | 3 | 21 | 19 |
| Clayton | 16 | 11 | 9 | 36 | 29 | 10 | 13 | 20 | 23 | 7 | 4 | 28 | 16 | 3 | 3 | 4 | 1 | 24 | 13 | 24 | 18 | 30 | 13 | 41 |
| Cobb | 19 | 71 | 48 | 111 | 26 | 48 | 56 | 47 | 11 | 67 | 27 | 83 | 32 | 11 | 0 | 23 | 74 | 81 | 41 | 30 | 6 | 41 | 93 | 59 |
| Coweta | 17 | 12 | 2 | 2 | 11 | 0 | 8 | 11 | 2 | 2 | 0 | 0 | 2 | 3 | 0 | 0 | 1 | 0 | 17 | 8 | 5 | 15 | 45 | 7 |
| Dawson | 0 | 1 | 0 | 5 | 1 | 2 | 0 | 0 | 0 | 1 | 5 | 3 | 0 | 0 | 0 | 0 | 2 | 9 | 4 | 2 | 0 | 0 | 2 | 1 |
| DeKalb | 36 | 71 | 29 | 73 | 39 | 49 | 44 | 47 | 19 | 50 | 27 | 32 | 87 | 49 | 2 | 54 | 62 | 61 | 59 | 42 | 66 | 111 | 48 | 116 |
| Douglas | 6 | 2 | 10 | 10 | 3 | 32 | 2 | 14 | 3 | 0 | 5 | 7 | 4 | 5 | 0 | 1 | 6 | 7 | 3 | 23 | 9 | 3 | 14 | 13 |
| Fayette | 2 | 4 | 2 | 0 | 1 | 0 | 5 | 4 | 2 | 1 | 0 | 2 | 1 | 1 | 1 | 3 | 2 | 0 | 0 | 3 | 4 | 1 | 1 | 11 |
| Forsyth | 1 | 9 | 6 | 9 | 5 | 3 | 2 | 22 | 0 | 0 | 5 | 2 | 2 | 1 | 1 | 6 | 7 | 22 | 9 | 1 | 2 | 3 | 31 | 12 |
| Fulton | 53 | 76 | 108 | 85 | 10 | 57 | 68 | 124 | 7 | 79 | 19 | 11 | 78 | 46 | 10 | 0 | 36 | 0 | 77 | 59 | 46 | 77 | 26 | 150 |
| Gwinnett | 46 | 77 | 45 | 120 | 32 | 68 | 81 | 44 | 28 | 20 | 15 | 42 | 45 | 0 | 0 | 83 | 91 | 49 | 62 | 79 | 36 | 59 | 142 | 109 |
| Hall | 6 | 76 | 70 | 68 | 42 | 41 | 5 | 10 | 27 | 21 | 52 | 13 | 15 | 6 | 4 | 14 | 27 | 39 | 18 | 21 | 36 | 17 | 5 | 51 |
| Haralson | 0 | 0 | 1 | 1 | 1 | 0 | 0 | 0 | 0 | 1 | 1 | 0 | 0 | 0 | 0 | 0 | 1 | 0 | 1 | 0 | 0 | 0 | 0 | 4 |
| Heard | 3 | 0 | 0 | 0 | 0 | 0 | 0 | 0 | 1 | 2 | 0 | 0 | 0 | 0 | 0 | 0 | 3 | 3 | 1 | 2 | 0 | 0 | 2 | 0 |
| Henry | 8 | 0 | 7 | 19 | 12 | 8 | 27 | 11 | 6 | 0 | 1 | 0 | 0 | 0 | 0 | 0 | 0 | 4 | 8 | 4 | 0 | 5 | 9 | 14 |
| Jasper | 1 | 1 | 0 | 1 | 0 | 0 | 0 | 0 | 0 | 0 | 0 | 0 | 0 | 0 | 0 | 0 | 0 | 0 | 0 | 5 | 0 | 0 | 0 | 1 |
| Lamar | 0 | 0 | 0 | 0 | 0 | 0 | 0 | 1 | 0 | 0 | 0 | 1 | 0 | 0 | 0 | 0 | 3 | 0 | 2 | 4 | 0 | 1 | 2 | 4 |
| Meriwether | 0 | 2 | 1 | 2 | 2 | 1 | 3 | 0 | 1 | 0 | 2 | 4 | 0 | 0 | 0 | 0 | 0 | 1 | 3 | 2 | 0 | 0 | 0 | 0 |
| Morgan | 0 | 1 | 1 | 0 | 0 | 0 | 2 | 1 | 0 | 0 | 0 | 0 | 0 | 0 | 0 | 1 | 0 | 0 | 0 | 1 | 1 | 0 | 1 | 0 |
| Newton | 3 | 7 | 7 | 10 | 4 | 4 | 12 | 6 | 2 | 1 | 0 | 0 | 0 | 0 | 3 | 0 | 2 | 9 | 5 | 6 | 3 | 15 | 7 | 24 |
| Paulding | 1 | 12 | 2 | 3 | 6 | 3 | 1 | 1 | 3 | 4 | 4 | 2 | 4 | 0 | 0 | 1 | 6 | 14 | 3 | 9 | 2 | 2 | 14 | 3 |
| Pickens | 2 | 0 | 0 | 0 | 2 | 1 | 1 | 0 | 0 | 2 | 1 | 0 | 0 | 0 | 0 | 2 | 1 | 0 | 1 | 1 | 2 | 0 | 0 | 0 |
| Pike | 0 | 0 | 0 | 0 | 1 | 0 | 0 | 3 | 0 | 0 | 0 | 0 | 0 | 0 | 0 | 0 | 0 | 1 | 0 | 3 | 0 | 1 | 1 | 2 |
| Rockdale | 1 | 4 | 5 | 19 | 3 | 2 | 2 | 9 | 4 | 2 | 3 | 1 | 0 | 0 | 0 | 1 | 1 | 0 | 3 | 3 | 1 | 7 | 2 | 1 |
| Spalding | 4 | 0 | 3 | 1 | 1 | 3 | 5 | 3 | 1 | 0 | 0 | 3 | 0 | 0 | 0 | 3 | 4 | 0 | 0 | 4 | 3 | 0 | 6 | 3 |
| Walton | 1 | 3 | 2 | 5 | 4 | 1 | 3 | 1 | 0 | 0 | 6 | 0 | 0 | 0 | 0 | 6 | 2 | 6 | 14 | 0 | 0 | 3 | 7 | 21 |

**Table S5.** Daily number of COVID-19 cases reported in the state of Georgia, Dougherty County and Metro Atlanta counties, May 27—June 14, 2020.

| Location | May |  |  |  |  | June |  |  |  |  |  |  |  |  |  |  |  |  |  |
| --- | --- | --- | --- | --- | --- | --- | --- | --- | --- | --- | --- | --- | --- | --- | --- | --- | --- | --- | --- |
|  | 27 | 28 | 29 | 30 | 31 | 1 | 2 | 3 | 4 | 5 | 6 | 7 | 8 | 9 | 10 | 11 | 12 | 13 | 14 |
| Georgia | 647 | 650 | 525 | 448 | 715 | 706 | 306 | 648 | 817 | 752 | 663 | 428 | 624 | 774 | 696 | 867 | 869 | 977 | 696 |
| Dougherty | 0 | 40 | 0 | 0 | 0 | 10 | 4 | 6 | 4 | 2 | 7 | 0 | 3 | 12 | 2 | 7 | 1 | 5 | 4 |
| Metro Atlanta | 675 | 417 | 513 | 118 | 91 | 374 | 105 | 508 | 576 | 463 | 233 | 32 | 439 | 385 | 404 | 509 | 405 | 236 | 115 |
| Bartow | 6 | 11 | 8 | 1 | 0 | 3 | 4 | 11 | 8 | 10 | 7 | 1 | 1 | 4 | 3 | 2 | 7 | 3 | 1 |
| Butts | 5 | 4 | 6 | 0 | 0 | 0 | 5 | 3 | 3 | 2 | 1 | 0 | 0 | 0 | 0 | 0 | 0 | 0 | 2 |
| Carroll | 22 | 4 | 1 | 1 | 6 | 0 | 0 | 11 | 33 | 10 | 2 | 0 | 5 | 2 | 4 | 2 | 9 | 9 | 3 |
| Cherokee | 11 | 8 | 16 | 5 | 19 | 1 | 1 | 12 | 11 | 10 | 14 | 1 | 16 | 3 | 12 | 10 | 14 | 3 | 12 |
| Clayton | 16 | 15 | 13 | 0 | 5 | 13 | 4 | 22 | 22 | 28 | 13 | 5 | 15 | 19 | 16 | 21 | 14 | 14 | 4 |
| Cobb | 53 | 41 | 39 | 22 | 18 | 25 | 4 | 41 | 68 | 58 | 19 | 7 | 49 | 39 | 33 | 101 | 51 | 35 | 24 |
| Coweta | 14 | 6 | 11 | 1 | 2 | 4 | 1 | 10 | 8 | 18 | 5 | 0 | 5 | 6 | 2 | 12 | 19 | 2 | 2 |
| Dawson | 4 | 0 | 1 | 0 | 0 | 0 | 0 | 1 | 0 | 2 | 1 | 0 | 1 | 0 | 2 | 2 | 0 | 0 | 0 |
| DeKalb | 152 | 62 | 83 | 13 | 3 | 61 | 8 | 71 | 65 | 47 | 22 | 4 | 42 | 66 | 66 | 43 | 53 | 21 | 8 |
| Douglas | 4 | 7 | 11 | 0 | 4 | 3 | 3 | 7 | 15 | 12 | 9 | 0 | 9 | 11 | 12 | 10 | 7 | 10 | 4 |
| Fayette | 3 | 0 | 0 | 0 | 0 | 0 | 2 | 2 | 2 | 3 | 1 | 0 | 2 | 7 | 0 | 4 | 1 | 1 | 1 |
| Forsyth | 10 | 5 | 5 | 0 | 0 | 11 | 9 | 2 | 1 | 6 | 5 | 0 | 10 | 19 | 4 | 0 | 10 | 5 | 1 |
| Fulton | 79 | 58 | 123 | 17 | 17 | 97 | 17 | 50 | 36 | 66 | 32 | 1 | 64 | 38 | 64 | 42 | 38 | 31 | 6 |
| Gwinnett | 173 | 139 | 114 | 36 | 11 | 90 | 21 | 172 | 204 | 105 | 61 | 7 | 158 | 94 | 116 | 172 | 109 | 52 | 31 |
| Hall | 44 | 28 | 15 | 2 | 0 | 40 | 1 | 31 | 27 | 30 | 19 | 1 | 26 | 22 | 22 | 13 | 28 | 13 | 7 |
| Haralson | 0 | 2 | 1 | 0 | 0 | 2 | 2 | 2 | 1 | 1 | 1 | 0 | 0 | 2 | 2 | 0 | 1 | 0 | 0 |
| Heard | 2 | 2 | 0 | 1 | 1 | 0 | 1 | 0 | 1 | 0 | 0 | 0 | 0 | 1 | 2 | 2 | 1 | 0 | 1 |
| Henry | 17 | 0 | 14 | 2 | 2 | 3 | 2 | 21 | 26 | 9 | 5 | 0 | 11 | 12 | 8 | 23 | 19 | 10 | 2 |
| Jasper | 0 | 0 | 1 | 3 | 0 | 4 | 0 | 0 | 5 | 1 | 1 | 1 | 1 | 2 | 0 | 4 | 1 | 0 | 0 |
| Lamar | 3 | 6 | 1 | 0 | 0 | 4 | 0 | 2 | 2 | 1 | 0 | 0 | 0 | 0 | 2 | 1 | 0 | 0 | 0 |
| Meriwether | 3 | 1 | 2 | 1 | 0 | 1 | 1 | 8 | 4 | 6 | 1 | 0 | 3 | 1 | 4 | 5 | 4 | 2 | 0 |
| Morgan | 0 | 0 | 0 | 0 | 0 | 1 | 0 | 0 | 0 | 1 | 0 | 0 | 0 | 1 | 0 | 0 | 0 | 0 | 0 |
| Newton | 6 | 3 | 7 | 1 | 0 | 9 | 9 | 6 | 6 | 6 | 1 | 0 | 6 | 1 | 6 | 5 | 5 | 3 | 0 |
| Paulding | 11 | 4 | 11 | 6 | 2 | 0 | 2 | 4 | 6 | 7 | 4 | 1 | 2 | 17 | 11 | 15 | 4 | 1 | 3 |
| Pickens | 0 | 2 | 2 | 0 | 1 | 0 | 0 | 2 | 0 | 0 | 1 | 1 | 0 | 1 | 4 | 2 | 0 | 6 | 0 |
| Pike | 2 | 0 | 0 | 0 | 0 | 0 | 0 | 2 | 1 | 2 | 1 | 1 | 1 | 2 | 1 | 0 | 1 | 3 | 0 |
| Rockdale | 8 | 2 | 5 | 2 | 0 | 0 | 4 | 8 | 5 | 2 | 4 | 0 | 6 | 6 | 5 | 4 | 4 | 5 | 2 |
| Spalding | 2 | 3 | 8 | 0 | 0 | 2 | 0 | 3 | 1 | 11 | 2 | 0 | 2 | 5 | 1 | 6 | 1 | 4 | 1 |
| Walton | 25 | 4 | 15 | 4 | 0 | 0 | 4 | 4 | 15 | 9 | 1 | 1 | 4 | 4 | 2 | 8 | 4 | 3 | 0 |

**Table S6.**
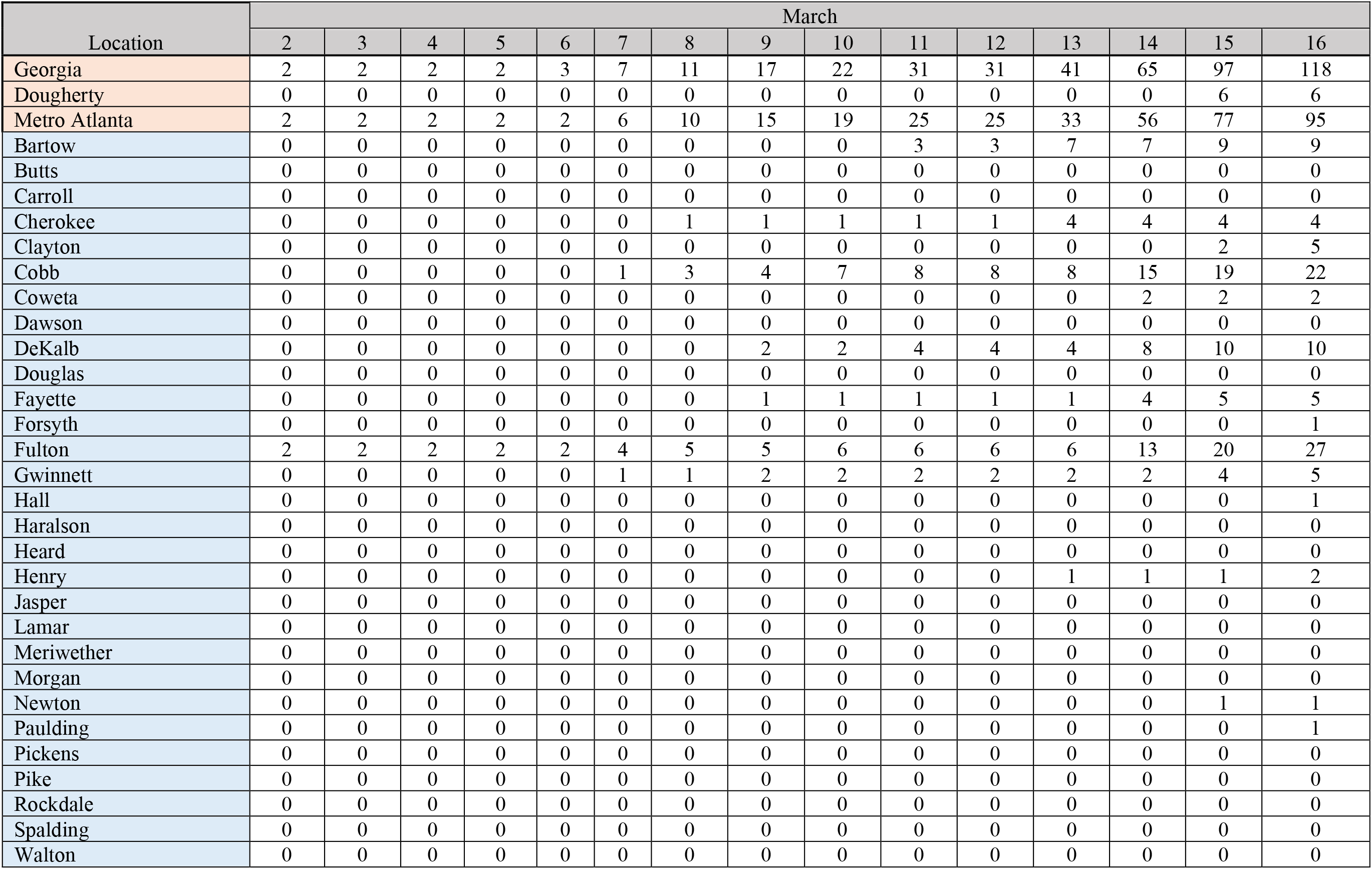
Cumulative number of reported COVID-19 cases in the state of Georgia, Dougherty County and Metro Atlanta counties, March 2—16, 2020.

**Table S7.** Cumulative number of reported COVID-19 cases in the state of Georgia, Dougherty County and Metro Atlanta counties, March 17—31, 2020.

| Date | March |  |  |  |  |  |  |  |  |  |  |  |  |  |  |
| --- | --- | --- | --- | --- | --- | --- | --- | --- | --- | --- | --- | --- | --- | --- | --- |
|  | 17 | 18 | 19 | 20 | 21 | 22 | 23 | 24 | 25 | 26 | 27 | 28 | 29 | 30 | 31 |
| Georgia | 142 | 193 | 282 | 482 | 552 | 620 | 800 | 1097 | 1387 | 1643 | 2198 | 2447 | 2683 | 3032 | 4117 |
| Dougherty | 6 | 7 | 22 | 45 | 47 | 52 | 69 | 101 | 123 | 164 | 203 | 224 | 247 | 278 | 466 |
| Metro Atlanta | 114 | 158 | 215 | 348 | 379 | 425 | 547 | 698 | 844 | 969 | 1366 | 1507 | 1689 | 1972 | 2455 |
| Bartow | 11 | 20 | 27 | 54 | 55 | 56 | 61 | 76 | 82 | 93 | 107 | 117 | 119 | 125 | 137 |
| Butts | 0 | 0 | 0 | 0 | 0 | 2 | 2 | 2 | 3 | 3 | 3 | 5 | 5 | 6 | 8 |
| Carroll | 0 | 0 | 0 | 14 | 16 | 16 | 21 | 26 | 50 | 52 | 61 | 64 | 70 | 97 | 123 |
| Cherokee | 4 | 6 | 13 | 16 | 17 | 18 | 24 | 30 | 36 | 44 | 50 | 54 | 60 | 69 | 78 |
| Clayton | 5 | 6 | 6 | 10 | 12 | 13 | 19 | 21 | 29 | 37 | 53 | 57 | 59 | 62 | 112 |
| Cobb | 25 | 29 | 38 | 51 | 56 | 67 | 79 | 90 | 109 | 119 | 163 | 185 | 228 | 250 | 287 |
| Coweta | 3 | 3 | 3 | 6 | 8 | 9 | 9 | 10 | 10 | 14 | 19 | 20 | 23 | 37 | 41 |
| Dawson | 0 | 0 | 0 | 1 | 1 | 1 | 1 | 1 | 1 | 3 | 3 | 3 | 4 | 7 | 10 |
| DeKalb | 15 | 18 | 22 | 40 | 42 | 53 | 74 | 107 | 125 | 137 | 219 | 246 | 273 | 294 | 360 |
| Douglas | 0 | 0 | 0 | 1 | 4 | 4 | 7 | 12 | 18 | 18 | 32 | 35 | 38 | 43 | 51 |
| Fayette | 5 | 7 | 8 | 8 | 9 | 9 | 11 | 12 | 12 | 14 | 22 | 26 | 27 | 42 | 48 |
| Forsyth | 1 | 2 | 3 | 3 | 4 | 5 | 7 | 8 | 10 | 15 | 21 | 22 | 28 | 36 | 47 |
| Fulton | 33 | 49 | 68 | 95 | 100 | 111 | 152 | 191 | 204 | 231 | 347 | 378 | 425 | 503 | 599 |
| Gwinnett | 7 | 7 | 12 | 23 | 23 | 27 | 35 | 46 | 69 | 79 | 121 | 131 | 145 | 178 | 242 |
| Hall | 1 | 3 | 5 | 6 | 9 | 9 | 10 | 16 | 19 | 22 | 30 | 31 | 33 | 34 | 67 |
| Haralson | 0 | 0 | 0 | 0 | 0 | 0 | 0 | 0 | 0 | 1 | 1 | 2 | 2 | 4 | 5 |
| Heard | 0 | 0 | 0 | 1 | 1 | 1 | 1 | 1 | 1 | 1 | 1 | 1 | 1 | 1 | 1 |
| Henry | 2 | 2 | 3 | 6 | 7 | 7 | 9 | 13 | 20 | 29 | 44 | 50 | 56 | 68 | 86 |
| Jasper | 0 | 0 | 0 | 0 | 0 | 0 | 0 | 1 | 2 | 2 | 2 | 2 | 2 | 2 | 2 |
| Lamar | 0 | 0 | 0 | 1 | 1 | 3 | 3 | 3 | 3 | 3 | 3 | 3 | 3 | 3 | 3 |
| Meriwether | 0 | 0 | 0 | 0 | 0 | 0 | 0 | 1 | 1 | 3 | 3 | 3 | 5 | 6 | 6 |
| Morgan | 0 | 0 | 0 | 0 | 0 | 0 | 1 | 1 | 1 | 1 | 1 | 1 | 1 | 1 | 2 |
| Newton | 1 | 3 | 3 | 4 | 4 | 4 | 4 | 6 | 8 | 12 | 15 | 15 | 18 | 22 | 31 |
| Paulding | 1 | 3 | 3 | 3 | 4 | 4 | 5 | 6 | 6 | 7 | 13 | 20 | 23 | 26 | 31 |
| Pickens | 0 | 0 | 0 | 2 | 2 | 2 | 3 | 4 | 4 | 4 | 4 | 5 | 5 | 6 | 7 |
| Pike | 0 | 0 | 0 | 0 | 0 | 0 | 0 | 0 | 0 | 0 | 0 | 1 | 2 | 2 | 2 |
| Rockdale | 0 | 0 | 1 | 1 | 2 | 2 | 5 | 8 | 10 | 13 | 16 | 17 | 18 | 29 | 45 |
| Spalding | 0 | 0 | 0 | 2 | 2 | 2 | 4 | 5 | 10 | 11 | 11 | 11 | 12 | 14 | 15 |
| Walton | 0 | 0 | 0 | 0 | 0 | 0 | 0 | 1 | 1 | 1 | 1 | 2 | 4 | 5 | 9 |

**Table S8.** Cumulative number of reported COVID-19 cases in the state of Georgia, Dougherty County and Metro Atlanta counties, April 1—18, 2020.

| Location | April |  |  |  |  |  |  |  |  |  |  |  |  |  |  |  |  |  |
| --- | --- | --- | --- | --- | --- | --- | --- | --- | --- | --- | --- | --- | --- | --- | --- | --- | --- | --- |
|  | 1 | 2 | 3 | 4 | 5 | 6 | 7 | 8 | 9 | 10 | 11 | 12 | 13 | 14 | 15 | 16 | 17 | 18 |
| Georgia | 4748 | 5444 | 5967 | 6383 | 6742 | 7558 | 9156 | 10204 | 10885 | 11859 | 12261 | 12452 | 13621 | 13913 | 14583 | 15644 | 16658 | 17841 |
| Dougherty | 490 | 521 | 607 | 685 | 688 | 722 | 973 | 1001 | 1042 | 1072 | 1102 | 1178 | 1245 | 1297 | 1320 | 1358 | 1385 | 1409 |
| Metro Atlanta | 2609 | 2979 | 3535 | 3890 | 3974 | 4404 | 4980 | 5294 | 5596 | 5976 | 6143 | 6384 | 7026 | 7700 | 8183 | 8610 | 9252 | 9443 |
| Bartow | 147 | 153 | 159 | 160 | 160 | 182 | 191 | 195 | 203 | 208 | 211 | 213 | 223 | 230 | 237 | 240 | 245 | 246 |
| Butts | 8 | 9 | 9 | 10 | 12 | 17 | 18 | 23 | 25 | 27 | 28 | 29 | 30 | 35 | 36 | 38 | 40 | 41 |
| Carroll | 133 | 139 | 147 | 158 | 158 | 163 | 183 | 184 | 195 | 200 | 204 | 210 | 229 | 235 | 242 | 258 | 261 | 271 |
| Cherokee | 85 | 94 | 114 | 120 | 124 | 141 | 147 | 155 | 167 | 177 | 186 | 187 | 204 | 219 | 232 | 245 | 274 | 285 |
| Clayton | 128 | 165 | 206 | 235 | 238 | 254 | 278 | 289 | 298 | 328 | 339 | 372 | 396 | 435 | 462 | 473 | 499 | 505 |
| Cobb | 304 | 341 | 422 | 453 | 474 | 517 | 566 | 608 | 653 | 681 | 705 | 728 | 816 | 895 | 950 | 1014 | 1085 | 1104 |
| Coweta | 42 | 48 | 56 | 64 | 67 | 76 | 80 | 82 | 87 | 100 | 101 | 106 | 121 | 135 | 138 | 142 | 153 | 160 |
| Dawson | 11 | 11 | 13 | 16 | 16 | 18 | 20 | 23 | 24 | 25 | 25 | 26 | 32 | 36 | 37 | 39 | 42 | 43 |
| DeKalb | 360 | 409 | 483 | 539 | 549 | 600 | 673 | 732 | 766 | 826 | 848 | 893 | 1006 | 1144 | 1225 | 1260 | 1366 | 1408 |
| Douglas | 56 | 66 | 78 | 84 | 91 | 105 | 117 | 124 | 134 | 148 | 151 | 159 | 177 | 189 | 201 | 209 | 222 | 229 |
| Fayette | 48 | 55 | 61 | 66 | 67 | 74 | 81 | 90 | 93 | 93 | 99 | 100 | 106 | 115 | 122 | 126 | 134 | 136 |
| Forsyth | 50 | 53 | 61 | 71 | 73 | 85 | 99 | 99 | 109 | 119 | 128 | 133 | 143 | 153 | 157 | 169 | 185 | 197 |
| Fulton | 638 | 747 | 910 | 959 | 970 | 1053 | 1185 | 1261 | 1336 | 1417 | 1446 | 1495 | 1635 | 1812 | 1902 | 1945 | 2037 | 2065 |
| Gwinnett | 257 | 303 | 353 | 400 | 410 | 455 | 540 | 586 | 618 | 669 | 681 | 701 | 766 | 815 | 885 | 917 | 1037 | 1059 |
| Hall | 71 | 72 | 81 | 117 | 117 | 138 | 215 | 233 | 247 | 273 | 276 | 282 | 319 | 363 | 402 | 526 | 605 | 607 |
| Haralson | 6 | 6 | 10 | 10 | 10 | 14 | 14 | 15 | 15 | 15 | 16 | 16 | 18 | 19 | 19 | 19 | 20 | 20 |
| Heard | 2 | 2 | 2 | 2 | 2 | 2 | 3 | 3 | 3 | 3 | 4 | 4 | 4 | 5 | 5 | 5 | 5 | 7 |
| Henry | 95 | 115 | 143 | 167 | 168 | 181 | 208 | 219 | 233 | 249 | 254 | 263 | 289 | 306 | 328 | 330 | 350 | 355 |
| Jasper | 2 | 2 | 2 | 4 | 4 | 4 | 6 | 6 | 6 | 6 | 6 | 8 | 9 | 9 | 10 | 10 | 14 | 15 |
| Lamar | 4 | 6 | 6 | 7 | 7 | 15 | 15 | 15 | 15 | 15 | 15 | 17 | 18 | 20 | 21 | 21 | 22 | 24 |
| Meriwether | 7 | 9 | 10 | 11 | 11 | 15 | 18 | 18 | 18 | 19 | 23 | 23 | 23 | 24 | 28 | 35 | 40 | 40 |
| Morgan | 2 | 2 | 2 | 2 | 2 | 6 | 10 | 12 | 13 | 14 | 14 | 14 | 15 | 15 | 17 | 19 | 19 | 20 |
| Newton | 34 | 37 | 42 | 52 | 55 | 65 | 67 | 67 | 70 | 77 | 78 | 82 | 93 | 100 | 113 | 115 | 121 | 121 |
| Paulding | 33 | 35 | 46 | 50 | 51 | 57 | 62 | 66 | 69 | 74 | 79 | 84 | 100 | 115 | 118 | 123 | 132 | 133 |
| Pickens | 7 | 9 | 9 | 9 | 9 | 9 | 10 | 10 | 10 | 10 | 11 | 11 | 11 | 13 | 14 | 15 | 16 | 18 |
| Pike | 2 | 2 | 4 | 5 | 5 | 9 | 10 | 11 | 12 | 15 | 16 | 16 | 19 | 22 | 25 | 28 | 29 | 30 |
| Rockdale | 47 | 57 | 67 | 75 | 77 | 82 | 85 | 85 | 85 | 89 | 92 | 97 | 101 | 106 | 107 | 111 | 117 | 119 |
| Spalding | 17 | 17 | 23 | 26 | 26 | 44 | 54 | 56 | 60 | 65 | 69 | 73 | 78 | 83 | 96 | 122 | 124 | 126 |
| Walton | 13 | 15 | 16 | 18 | 21 | 23 | 25 | 27 | 32 | 34 | 38 | 42 | 45 | 52 | 54 | 56 | 58 | 59 |

**Table S9.** Cumulative number of reported COVID-19 cases in the state of Georgia, Dougherty County and Metro Atlanta counties, April 19—30, 2020.

| Location | April |  |  |  |  |  |  |  |  |  |  |  |
| --- | --- | --- | --- | --- | --- | --- | --- | --- | --- | --- | --- | --- |
|  | 19 | 20 | 21 | 22 | 23 | 24 | 25 | 26 | 27 | 28 | 29 | 30 |
| Georgia | 17619 | 18447 | 19189 | 20099 | 20905 | 21575 | 22225 | 22459 | 23229 | 23607 | 24300 | 25431 |
| Dougherty | 1425 | 1436 | 1456 | 1456 | 1456 | 1465 | 1470 | 1470 | 1480 | 1493 | 1500 | 1506 |
| Metro Atlanta | 9827 | 10297 | 10543 | 10975 | 11643 | 11965 | 12314 | 12435 | 12904 | 13464 | 13937 | 14305 |
| Bartow | 250 | 255 | 256 | 259 | 264 | 273 | 283 | 286 | 289 | 306 | 309 | 315 |
| Butts | 50 | 80 | 81 | 125 | 125 | 125 | 125 | 126 | 126 | 130 | 134 | 145 |
| Carroll | 282 | 305 | 313 | 313 | 314 | 320 | 325 | 326 | 329 | 342 | 348 | 358 |
| Cherokee | 300 | 317 | 324 | 340 | 365 | 367 | 368 | 374 | 393 | 436 | 457 | 467 |
| Clayton | 520 | 540 | 547 | 549 | 616 | 629 | 641 | 641 | 649 | 672 | 693 | 716 |
| Cobb | 1148 | 1196 | 1230 | 1272 | 1326 | 1368 | 1395 | 1428 | 1483 | 1539 | 1581 | 1621 |
| Coweta | 164 | 169 | 169 | 169 | 177 | 187 | 187 | 188 | 190 | 191 | 195 | 195 |
| Dawson | 44 | 45 | 45 | 49 | 50 | 51 | 51 | 51 | 53 | 58 | 60 | 61 |
| DeKalb | 1473 | 1520 | 1563 | 1609 | 1689 | 1721 | 1788 | 1800 | 1855 | 1907 | 1980 | 2027 |
| Douglas | 233 | 243 | 246 | 259 | 273 | 276 | 276 | 278 | 284 | 300 | 311 | 319 |
| Fayette | 139 | 143 | 144 | 146 | 156 | 158 | 161 | 161 | 169 | 171 | 176 | 176 |
| Forsyth | 202 | 211 | 216 | 227 | 235 | 241 | 249 | 252 | 271 | 284 | 292 | 299 |
| Fulton | 2131 | 2198 | 2206 | 2255 | 2436 | 2500 | 2543 | 2545 | 2680 | 2753 | 2785 | 2821 |
| Gwinnett | 1124 | 1181 | 1238 | 1273 | 1351 | 1382 | 1460 | 1504 | 1545 | 1620 | 1744 | 1793 |
| Hall | 625 | 702 | 756 | 907 | 963 | 1022 | 1032 | 1033 | 1098 | 1185 | 1261 | 1346 |
| Haralson | 20 | 22 | 25 | 25 | 26 | 26 | 26 | 26 | 27 | 28 | 28 | 28 |
| Heard | 7 | 7 | 8 | 8 | 9 | 9 | 9 | 9 | 10 | 10 | 10 | 10 |
| Henry | 371 | 382 | 384 | 386 | 413 | 419 | 436 | 451 | 464 | 488 | 492 | 492 |
| Jasper | 17 | 17 | 18 | 18 | 19 | 20 | 20 | 20 | 20 | 22 | 22 | 22 |
| Lamar | 25 | 27 | 29 | 29 | 31 | 31 | 33 | 33 | 34 | 35 | 37 | 38 |
| Meriwether | 42 | 46 | 47 | 48 | 49 | 49 | 49 | 49 | 49 | 53 | 54 | 54 |
| Morgan | 22 | 22 | 23 | 23 | 25 | 25 | 25 | 25 | 25 | 25 | 27 | 27 |
| Newton | 130 | 139 | 142 | 146 | 153 | 158 | 166 | 166 | 173 | 184 | 193 | 196 |
| Paulding | 137 | 141 | 141 | 145 | 148 | 156 | 159 | 160 | 166 | 172 | 176 | 189 |
| Pickens | 18 | 18 | 18 | 18 | 19 | 20 | 20 | 21 | 21 | 26 | 26 | 26 |
| Pike | 33 | 33 | 34 | 35 | 38 | 38 | 39 | 39 | 39 | 39 | 39 | 40 |
| Rockdale | 131 | 140 | 141 | 144 | 151 | 158 | 161 | 161 | 166 | 175 | 178 | 189 |
| Spalding | 128 | 135 | 136 | 136 | 140 | 144 | 189 | 190 | 197 | 201 | 205 | 210 |
| Walton | 61 | 63 | 63 | 73 | 81 | 92 | 97 | 97 | 100 | 113 | 121 | 126 |

**Table S10.** Cumulative number of COVID-19 cases reported in the state of Georgia, Dougherty County and Metro Atlanta counties, May 1—15, 2020.

| Location | May |  |  |  |  |  |  |  |  |  |  |  |  |  |  |
| --- | --- | --- | --- | --- | --- | --- | --- | --- | --- | --- | --- | --- | --- | --- | --- |
|  | 1 | 2 | 3 | 4 | 5 | 6 | 7 | 8 | 9 | 10 | 11 | 12 | 13 | 14 | 15 |
| Georgia | 26436 | 27268 | 27618 | 28350 | 28876 | 29724 | 30524 | 31089 | 31481 | 32267 | 32448 | 33311 | 33866 | 34422 | 35242 |
| Dougherty | 1531 | 1534 | 1536 | 1543 | 1550 | 1555 | 1566 | 1583 | 1587 | 1593 | 1609 | 1643 | 1644 | 1643 | 1662 |
| Metro Atlanta | 14771 | 15399 | 15638 | 16108 | 16497 | 17119 | 17382 | 17731 | 18089 | 18502 | 18668 | 18948 | 19155 | 19426 | 19749 |
| Bartow | 323 | 324 | 325 | 341 | 351 | 359 | 359 | 361 | 361 | 363 | 364 | 368 | 377 | 382 | 385 |
| Butts | 151 | 151 | 150 | 152 | 158 | 163 | 167 | 171 | 172 | 180 | 189 | 191 | 192 | 196 | 196 |
| Carroll | 375 | 383 | 386 | 382 | 387 | 391 | 389 | 390 | 393 | 395 | 395 | 398 | 397 | 400 | 408 |
| Cherokee | 474 | 482 | 491 | 509 | 519 | 544 | 569 | 578 | 593 | 609 | 621 | 635 | 655 | 676 | 695 |
| Clayton | 733 | 777 | 793 | 804 | 813 | 849 | 878 | 888 | 901 | 921 | 944 | 951 | 955 | 983 | 999 |
| Cobb | 1679 | 1749 | 1768 | 1839 | 1887 | 1998 | 2024 | 2072 | 2128 | 2175 | 2186 | 2253 | 2280 | 2363 | 2395 |
| Coweta | 211 | 213 | 230 | 242 | 244 | 246 | 257 | 257 | 265 | 276 | 278 | 280 | 280 | 280 | 282 |
| Dawson | 61 | 66 | 66 | 67 | 67 | 72 | 73 | 75 | 75 | 81 | 81 | 76 | 81 | 84 | 84 |
| DeKalb | 2073 | 2148 | 2184 | 2255 | 2284 | 2357 | 2396 | 2445 | 2489 | 2536 | 2555 | 2605 | 2632 | 2664 | 2751 |
| Douglas | 329 | 335 | 341 | 343 | 353 | 363 | 366 | 398 | 400 | 414 | 417 | 417 | 422 | 429 | 433 |
| Fayette | 178 | 181 | 183 | 187 | 189 | 188 | 191 | 190 | 195 | 199 | 201 | 202 | 202 | 204 | 205 |
| Forsyth | 309 | 331 | 332 | 341 | 347 | 356 | 361 | 364 | 366 | 388 | 391 | 386 | 393 | 395 | 397 |
| Fulton | 2880 | 2928 | 2981 | 3057 | 3165 | 3250 | 3260 | 3317 | 3385 | 3509 | 3516 | 3595 | 3614 | 3625 | 3703 |
| Gwinnett | 1848 | 1934 | 1980 | 2057 | 2102 | 2222 | 2254 | 2322 | 2403 | 2447 | 2475 | 2495 | 2510 | 2552 | 2597 |
| Hall | 1482 | 1694 | 1700 | 1776 | 1846 | 1914 | 1956 | 1997 | 2002 | 2012 | 2039 | 2060 | 2112 | 2125 | 2140 |
| Haralson | 29 | 29 | 29 | 29 | 30 | 31 | 32 | 32 | 32 | 32 | 32 | 33 | 34 | 34 | 34 |
| Heard | 11 | 11 | 14 | 13 | 14 | 14 | 14 | 14 | 14 | 14 | 15 | 17 | 17 | 17 | 17 |
| Henry | 492 | 512 | 520 | 520 | 527 | 546 | 558 | 566 | 593 | 604 | 610 | 614 | 615 | 621 | 628 |
| Jasper | 22 | 23 | 24 | 25 | 25 | 26 | 26 | 26 | 25 | 25 | 25 | 26 | 26 | 26 | 26 |
| Lamar | 39 | 40 | 40 | 39 | 39 | 39 | 38 | 39 | 39 | 40 | 40 | 40 | 40 | 41 | 41 |
| Meriwether | 54 | 54 | 54 | 56 | 57 | 59 | 61 | 62 | 65 | 65 | 66 | 66 | 68 | 72 | 73 |
| Morgan | 27 | 27 | 28 | 29 | 30 | 30 | 30 | 30 | 32 | 33 | 33 | 33 | 33 | 33 | 33 |
| Newton | 200 | 205 | 208 | 215 | 222 | 232 | 236 | 240 | 252 | 258 | 260 | 261 | 262 | 263 | 262 |
| Paulding | 190 | 194 | 195 | 207 | 209 | 212 | 218 | 221 | 222 | 223 | 226 | 230 | 234 | 236 | 240 |
| Pickens | 26 | 27 | 29 | 29 | 28 | 29 | 31 | 32 | 33 | 33 | 33 | 35 | 36 | 35 | 35 |
| Pike | 40 | 40 | 40 | 40 | 40 | 40 | 41 | 41 | 41 | 44 | 44 | 47 | 45 | 44 | 44 |
| Rockdale | 191 | 194 | 195 | 199 | 204 | 223 | 226 | 228 | 230 | 239 | 243 | 245 | 248 | 249 | 249 |
| Spalding | 214 | 215 | 219 | 219 | 222 | 223 | 224 | 227 | 232 | 235 | 237 | 237 | 236 | 239 | 239 |
| Walton | 129 | 132 | 133 | 136 | 138 | 143 | 147 | 148 | 151 | 152 | 152 | 152 | 159 | 158 | 158 |

**Table S11.** Cumulative number of COVID-19 cases reported in the state of Georgia, Dougherty County and Metro Atlanta counties, May 16—31, 2020.

| Location | May |  |  |  |  |  |  |  |  |  |  |  |  |  |  |  |
| --- | --- | --- | --- | --- | --- | --- | --- | --- | --- | --- | --- | --- | --- | --- | --- | --- |
|  | 16 | 17 | 18 | 19 | 20 | 21 | 22 | 23 | 24 | 25 | 26 | 27 | 28 | 29 | 30 | 31 |
| Georgia | 35655 | 35954 | 36634 | 37214 | 38162 | 38969 | 39734 | 40408 | 40989 | 41414 | 42066 | 42713 | 43363 | 43888 | 44336 | 45051 |
| Dougherty | 1662 | 1661 | 1664 | 1668 | 1672 | 1716 | 1723 | 1725 | 1727 | 1730 | 1774 | 1784 | 1764 | 1763 | 1770 | 1770 |
| Metro Atlanta | 19884 | 19899 | 20130 | 20502 | 20873 | 21239 | 21602 | 21878 | 22278 | 22799 | 23495 | 24170 | 24583 | 25099 | 25218 | 25308 |
| Bartow | 385 | 384 | 396 | 410 | 419 | 424 | 427 | 444 | 443 | 443 | 452 | 458 | 469 | 477 | 478 | 478 |
| Butts | 197 | 197 | 199 | 199 | 205 | 205 | 206 | 208 | 209 | 211 | 214 | 219 | 223 | 229 | 229 | 228 |
| Carroll | 409 | 406 | 408 | 418 | 425 | 437 | 439 | 440 | 445 | 472 | 490 | 512 | 516 | 517 | 518 | 525 |
| Cherokee | 700 | 714 | 723 | 745 | 758 | 769 | 790 | 807 | 810 | 831 | 850 | 861 | 869 | 885 | 890 | 909 |
| Clayton | 1002 | 1005 | 1009 | 1010 | 1034 | 1047 | 1071 | 1089 | 1119 | 1132 | 1173 | 1189 | 1204 | 1217 | 1217 | 1221 |
| Cobb | 2407 | 2406 | 2429 | 2503 | 2584 | 2625 | 2655 | 2661 | 2702 | 2795 | 2854 | 2907 | 2948 | 2987 | 3009 | 3027 |
| Coweta | 285 | 284 | 285 | 287 | 286 | 303 | 311 | 316 | 331 | 376 | 383 | 397 | 403 | 414 | 415 | 417 |
| Dawson | 84 | 83 | 84 | 86 | 95 | 99 | 101 | 101 | 101 | 103 | 104 | 108 | 108 | 109 | 109 | 109 |
| DeKalb | 2800 | 2802 | 2856 | 2918 | 2979 | 3038 | 3080 | 3146 | 3257 | 3305 | 3421 | 3573 | 3635 | 3718 | 3731 | 3734 |
| Douglas | 438 | 437 | 439 | 445 | 452 | 455 | 478 | 487 | 490 | 504 | 517 | 521 | 528 | 539 | 539 | 543 |
| Fayette | 206 | 207 | 210 | 212 | 215 | 212 | 215 | 219 | 220 | 221 | 232 | 235 | 231 | 234 | 235 | 235 |
| Forsyth | 398 | 399 | 405 | 412 | 434 | 443 | 444 | 446 | 449 | 480 | 492 | 502 | 507 | 512 | 512 | 512 |
| Fulton | 3749 | 3759 | 3751 | 3795 | 3793 | 3872 | 3931 | 3977 | 4054 | 4080 | 4230 | 4309 | 4367 | 4490 | 4507 | 4524 |
| Gwinnett | 2597 | 2594 | 2680 | 2771 | 2820 | 2882 | 2961 | 2997 | 3056 | 3198 | 3307 | 3480 | 3619 | 3733 | 3769 | 3780 |
| Hall | 2146 | 2150 | 2164 | 2191 | 2230 | 2248 | 2269 | 2305 | 2322 | 2327 | 2378 | 2422 | 2450 | 2465 | 2467 | 2467 |
| Haralson | 34 | 34 | 34 | 35 | 35 | 36 | 36 | 36 | 36 | 36 | 40 | 40 | 42 | 43 | 43 | 43 |
| Heard | 17 | 17 | 17 | 20 | 23 | 24 | 26 | 26 | 26 | 28 | 28 | 30 | 32 | 32 | 33 | 34 |
| Henry | 630 | 627 | 627 | 611 | 615 | 623 | 627 | 626 | 632 | 641 | 655 | 672 | 672 | 686 | 688 | 690 |
| Jasper | 26 | 26 | 26 | 26 | 26 | 26 | 31 | 31 | 31 | 31 | 32 | 32 | 32 | 33 | 36 | 36 |
| Lamar | 41 | 41 | 41 | 44 | 44 | 46 | 50 | 50 | 51 | 53 | 57 | 60 | 66 | 67 | 67 | 67 |
| Meriwether | 72 | 72 | 70 | 70 | 73 | 76 | 78 | 78 | 78 | 78 | 78 | 81 | 82 | 84 | 85 | 85 |
| Morgan | 33 | 33 | 34 | 34 | 34 | 34 | 35 | 36 | 36 | 37 | 37 | 37 | 37 | 37 | 37 | 37 |
| Newton | 261 | 257 | 264 | 266 | 275 | 280 | 286 | 289 | 304 | 311 | 335 | 341 | 344 | 351 | 352 | 352 |
| Paulding | 242 | 240 | 241 | 247 | 261 | 264 | 273 | 275 | 277 | 291 | 294 | 305 | 309 | 320 | 326 | 328 |
| Pickens | 35 | 35 | 38 | 39 | 39 | 40 | 41 | 43 | 43 | 43 | 43 | 43 | 45 | 47 | 47 | 48 |
| Pike | 44 | 44 | 44 | 44 | 45 | 45 | 48 | 48 | 49 | 50 | 52 | 54 | 54 | 54 | 54 | 54 |
| Rockdale | 249 | 249 | 250 | 252 | 251 | 254 | 257 | 258 | 265 | 267 | 268 | 276 | 278 | 283 | 285 | 285 |
| Spalding | 239 | 239 | 242 | 246 | 251 | 246 | 250 | 253 | 253 | 259 | 262 | 264 | 267 | 275 | 275 | 275 |
| Walton | 158 | 158 | 164 | 166 | 172 | 186 | 186 | 186 | 189 | 196 | 217 | 242 | 246 | 261 | 265 | 265 |

**Table S12.**
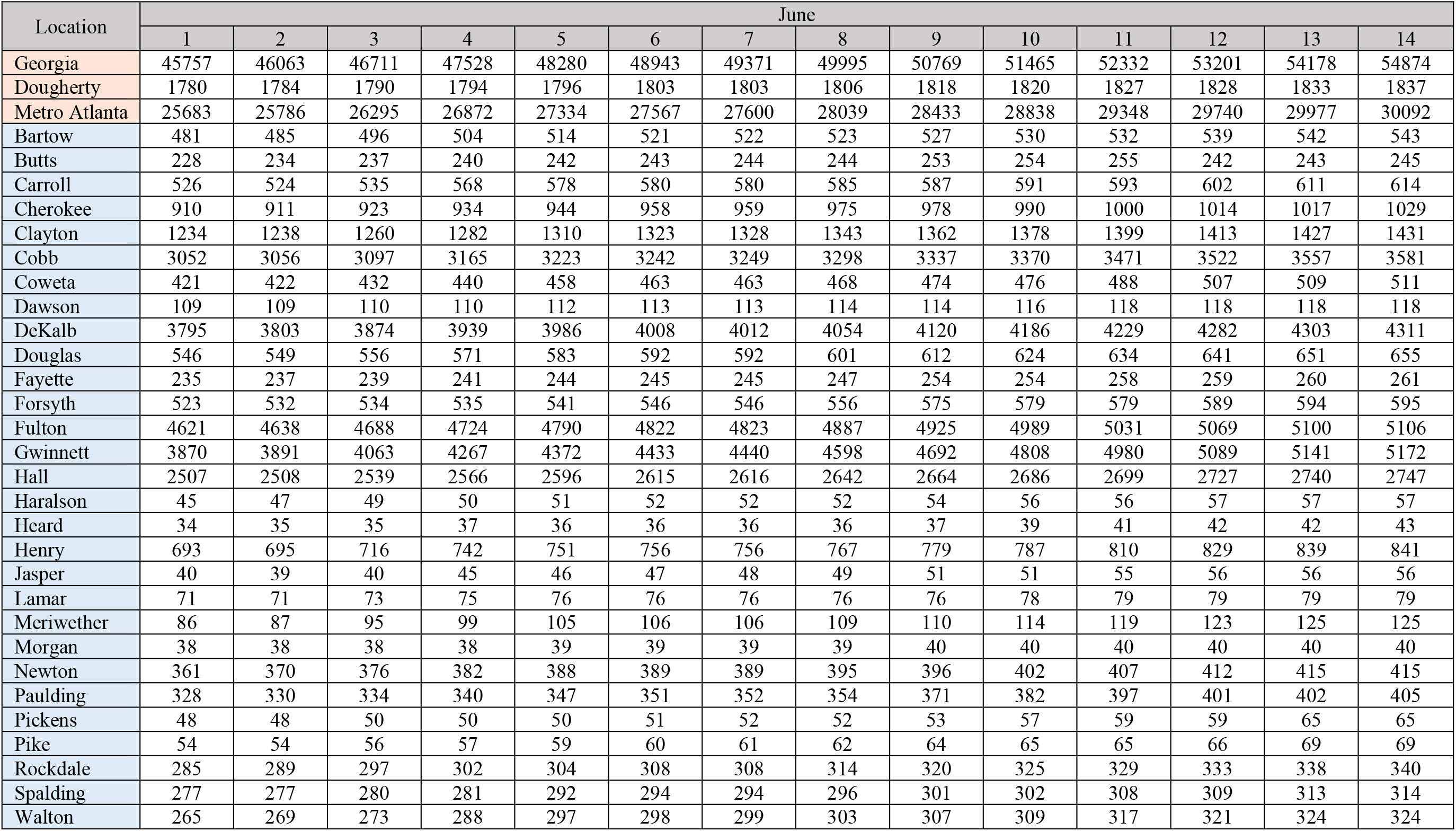
Cumulative number of COVID-19 cases reported in the state of Georgia, Dougherty County and Metro Atlanta counties, June 1—14, 2020.

### Legends of Supplementary Figures: Results for Counties in Metro Atlanta, GA, USA

**Figure S1.**
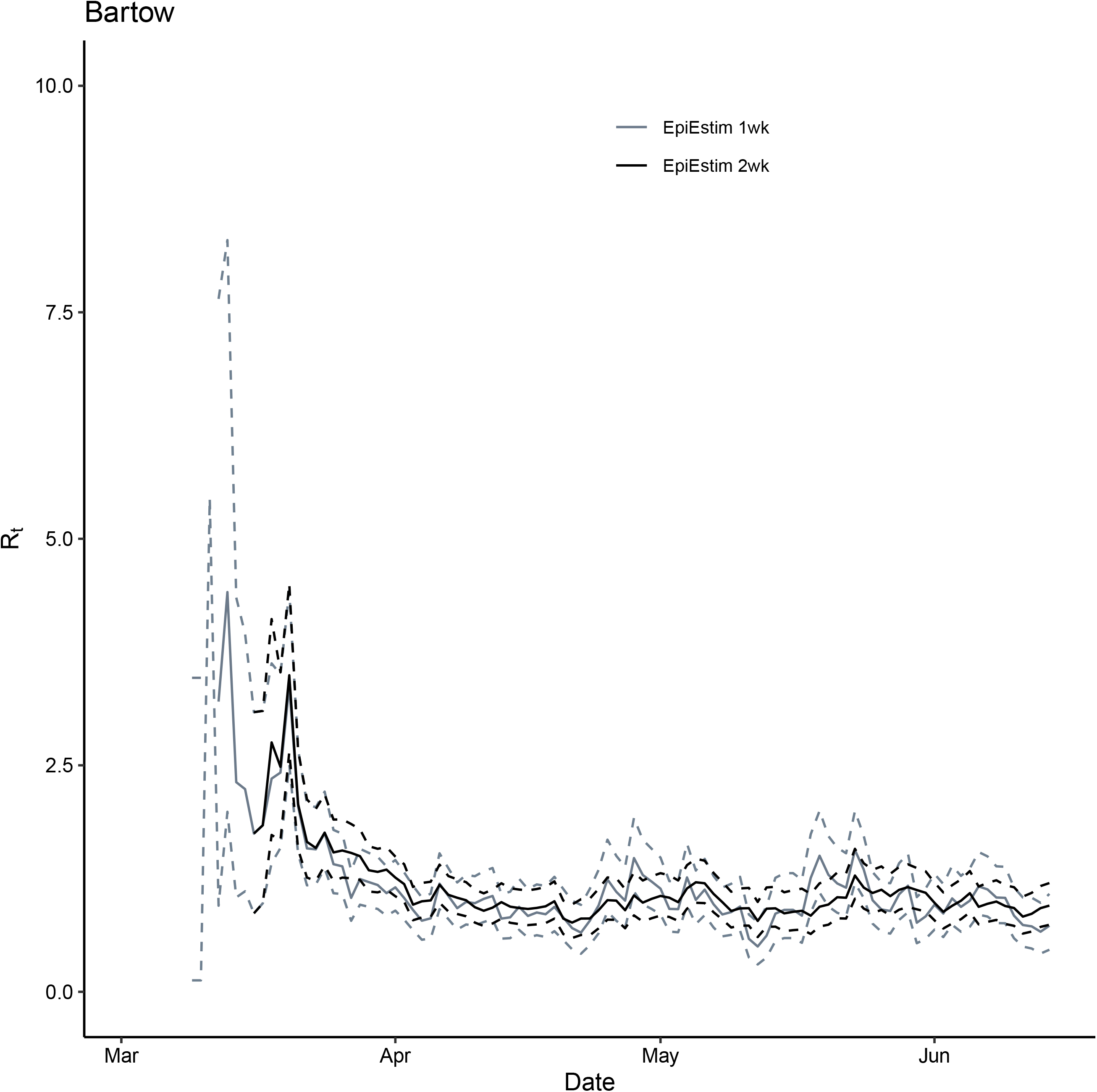
Comparison between *R*_*t*_ for Bartow County, GA, March 2—June 14, 2020, estimated using the instantaneous reproduction number method implemented in EpiEstim package (grey: 1-week window and black: 2-week window). Solid lines represent the median estimates and the dashed line represent the 2.5 and 97.5 quantiles.

**Figure S2.**
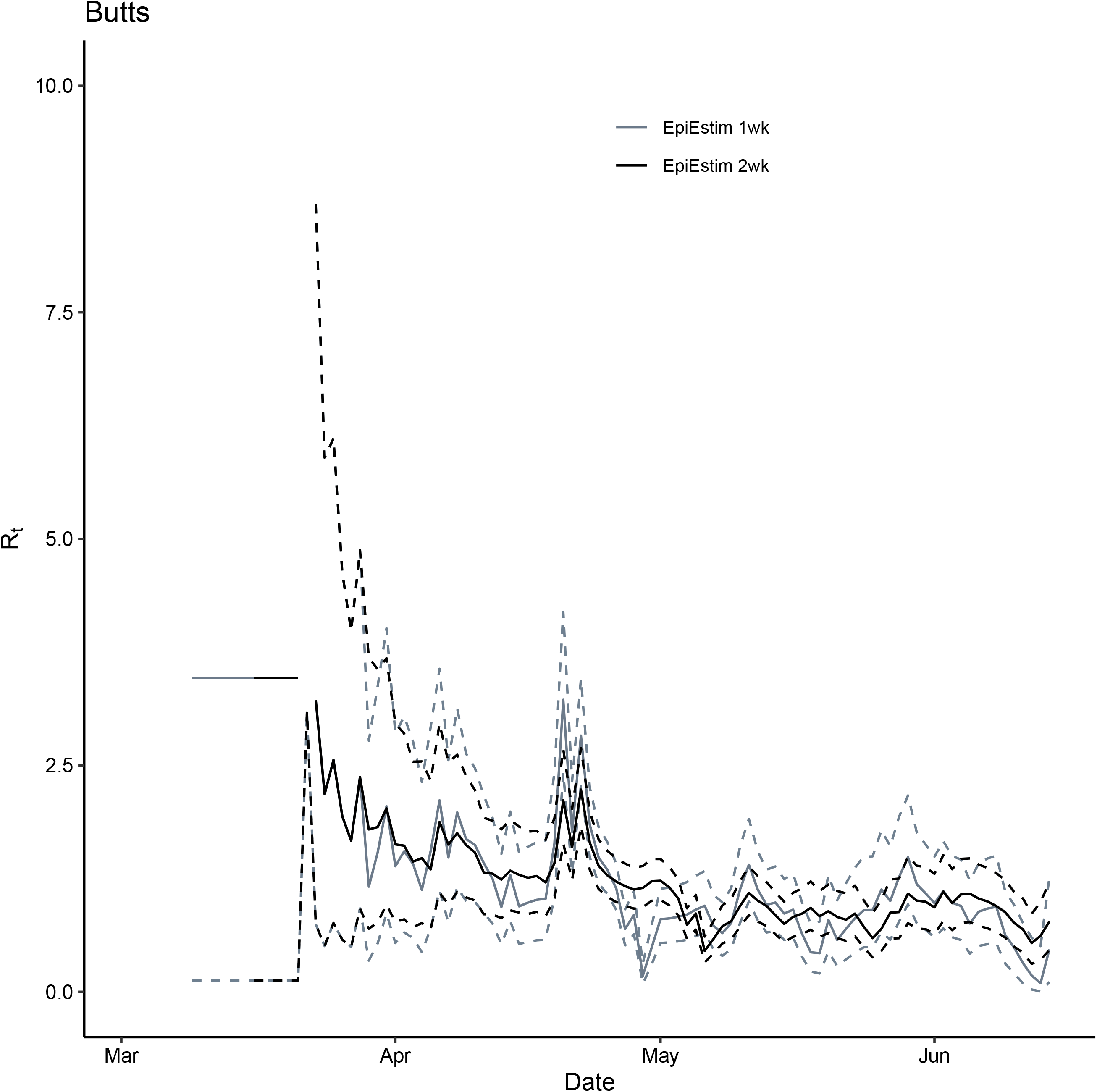
Comparison between *R*_*t*_ for Butts County, GA, March 2—June 14, 2020, estimated using the instantaneous reproduction number method implemented in EpiEstim package (grey: 1-week window and black: 2-week window). Solid lines represent the median estimates and the dashed line represent the 2.5 and 97.5 quantiles.

**Figure S3.**
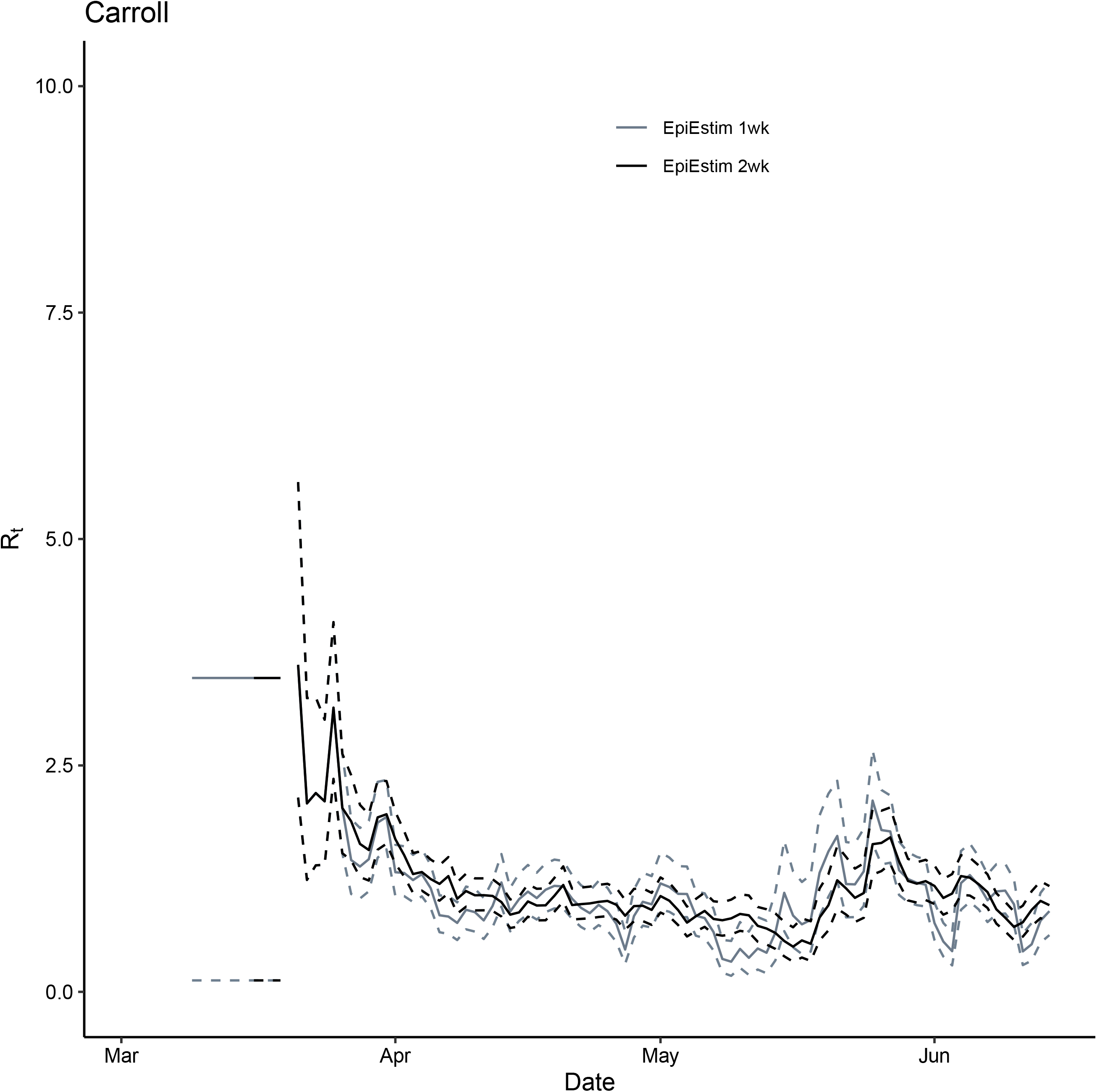
Comparison between *R*_*t*_ for Carroll County, GA, March 2—June 14, 2020, estimated using the instantaneous reproduction number method implemented in EpiEstim package (grey: 1-week window and black: 2-week window). Solid lines represent the median estimates and the dashed line represent the 2.5 and 97.5 quantiles.

**Figure S4.**
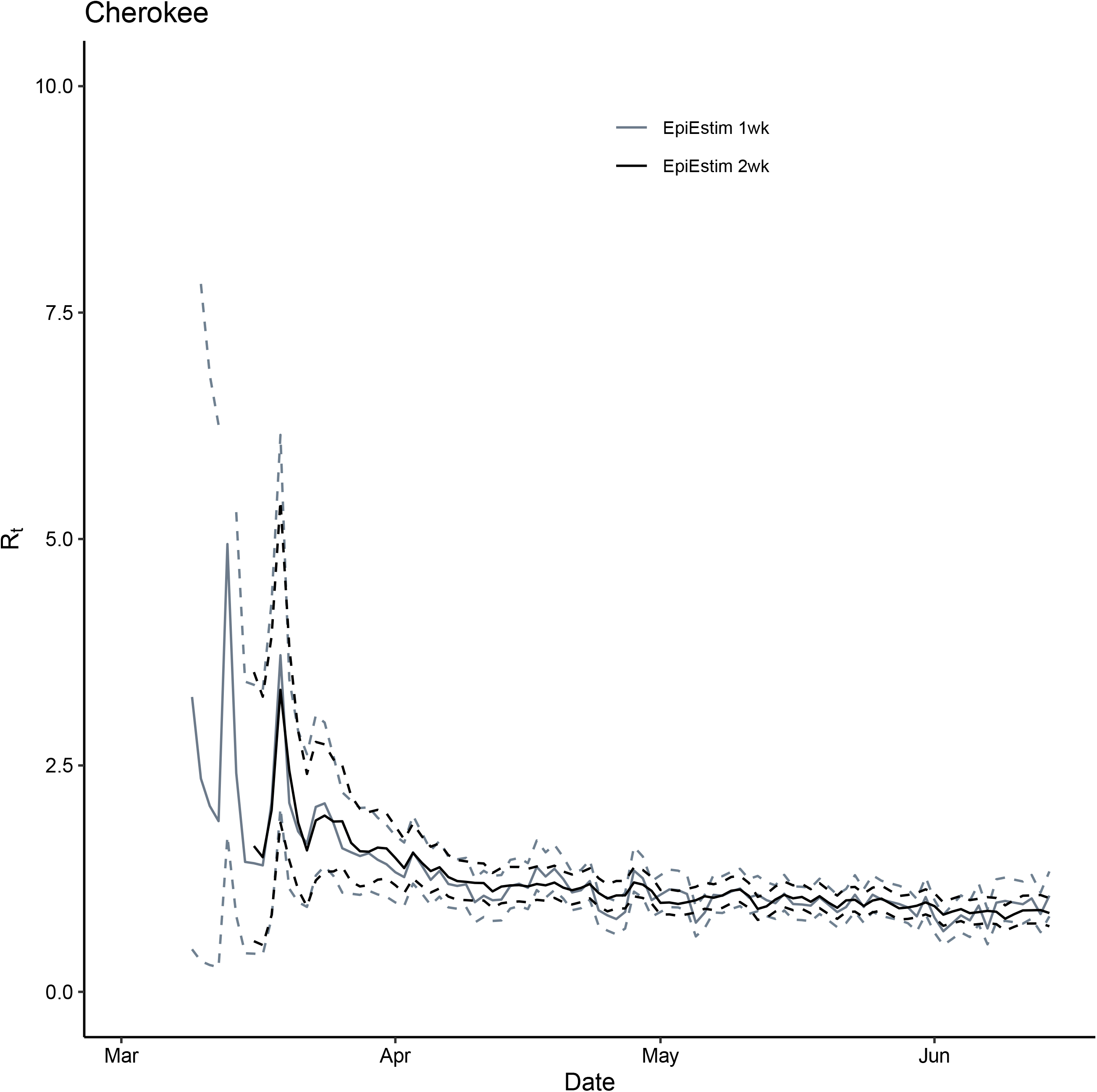
Comparison between *R*_*t*_ for Cherokee County, GA, March 2—June 14, 2020, estimated using the instantaneous reproduction number method implemented in EpiEstim package (grey: 1-week window and black: 2-week window). Solid lines represent the median estimates and the dashed line represent the 2.5 and 97.5 quantiles.

**Figure S5.**
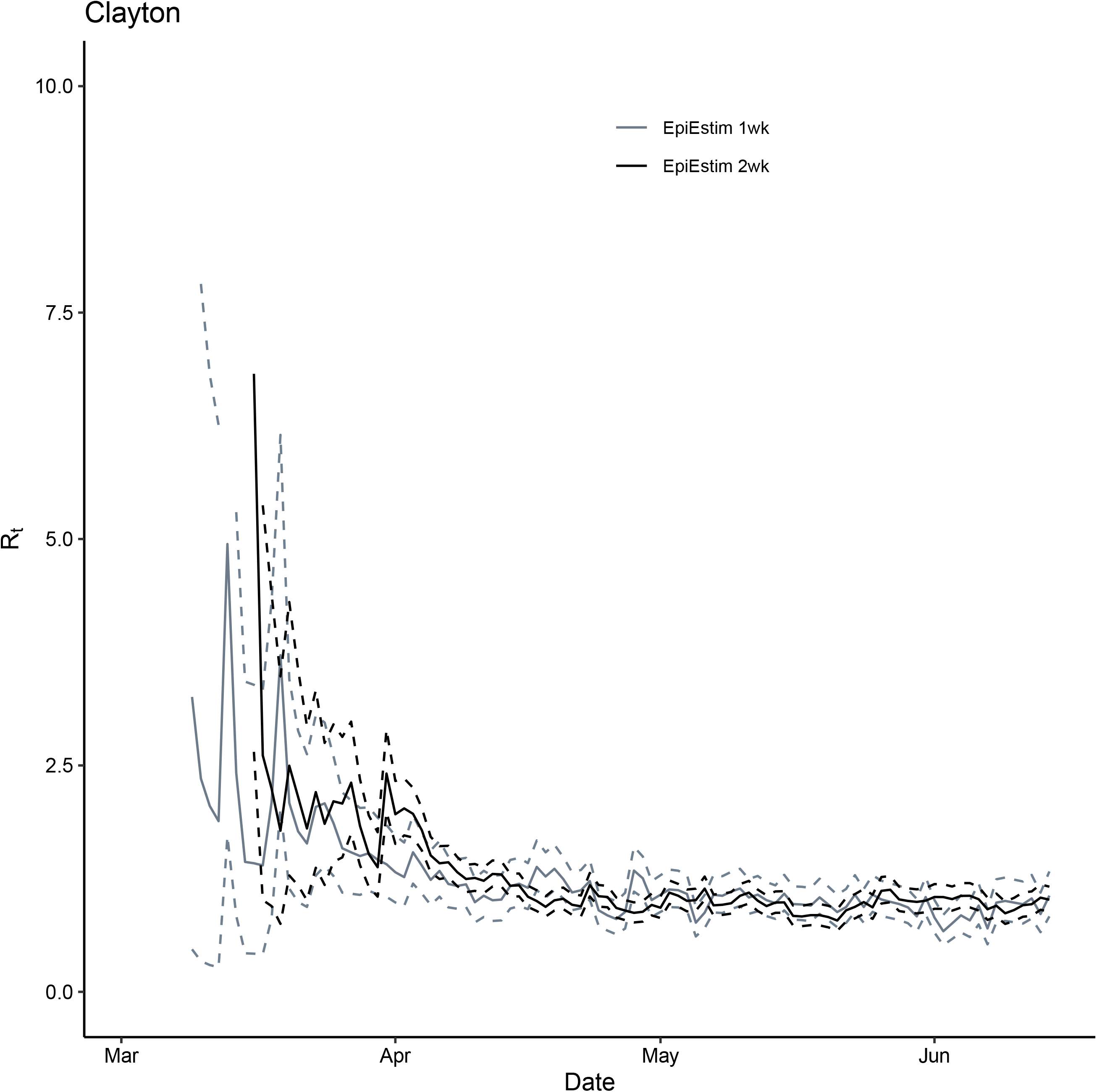
Comparison between *R*_*t*_ for Clayton County, GA, March 2—June 14, 2020, estimated using the instantaneous reproduction number method implemented in EpiEstim package (grey: 1-week window and black: 2-week window). Solid lines represent the median estimates and the dashed line represent the 2.5 and 97.5 quantiles.

**Figure S6.**
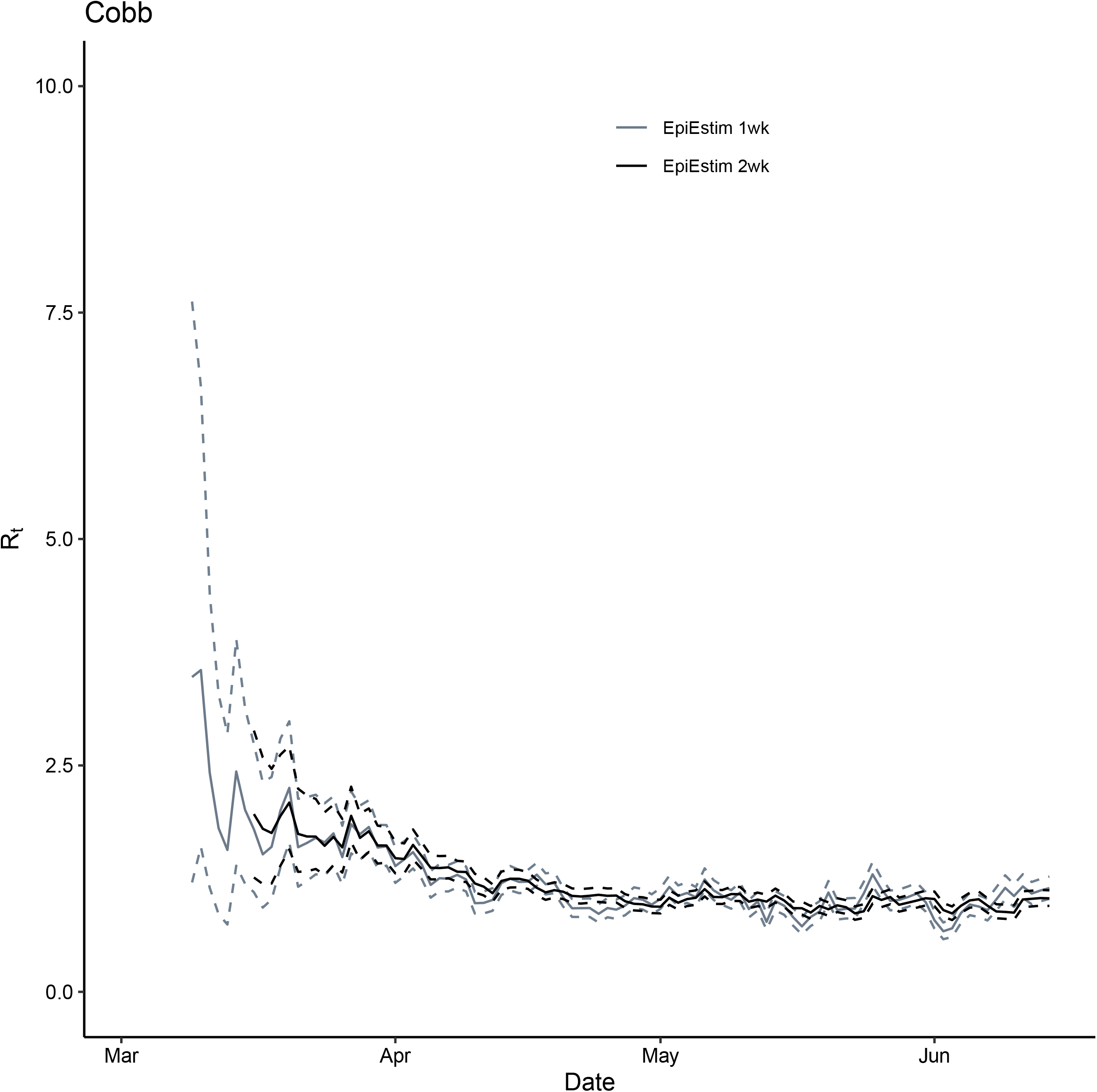
Comparison between *R*_*t*_ for Cobb County, GA, March 2—June 14, 2020, estimated using the instantaneous reproduction number method implemented in EpiEstim package (grey: 1-week window and black: 2-week window). Solid lines represent the median estimates and the dashed line represent the 2.5 and 97.5 quantiles.

**Figure S7.**
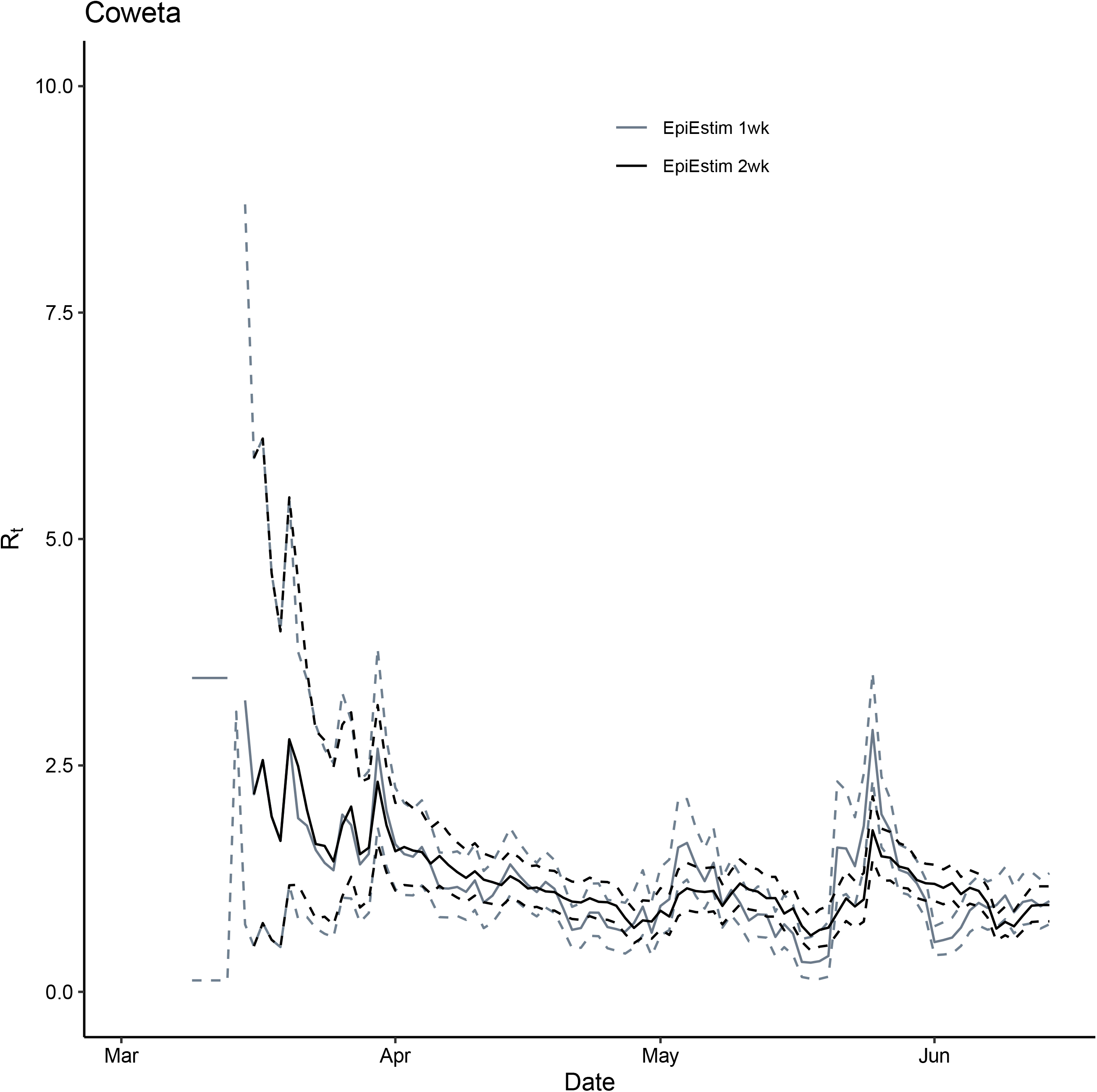
Comparison between *R*_*t*_ for Coweta County, GA, March 2—June 14, 2020, estimated using the instantaneous reproduction number method implemented in EpiEstim package (grey: 1-week window and black: 2-week window). Solid lines represent the median estimates and the dashed line represent the 2.5 and 97.5 quantiles.

**Figure S8.**
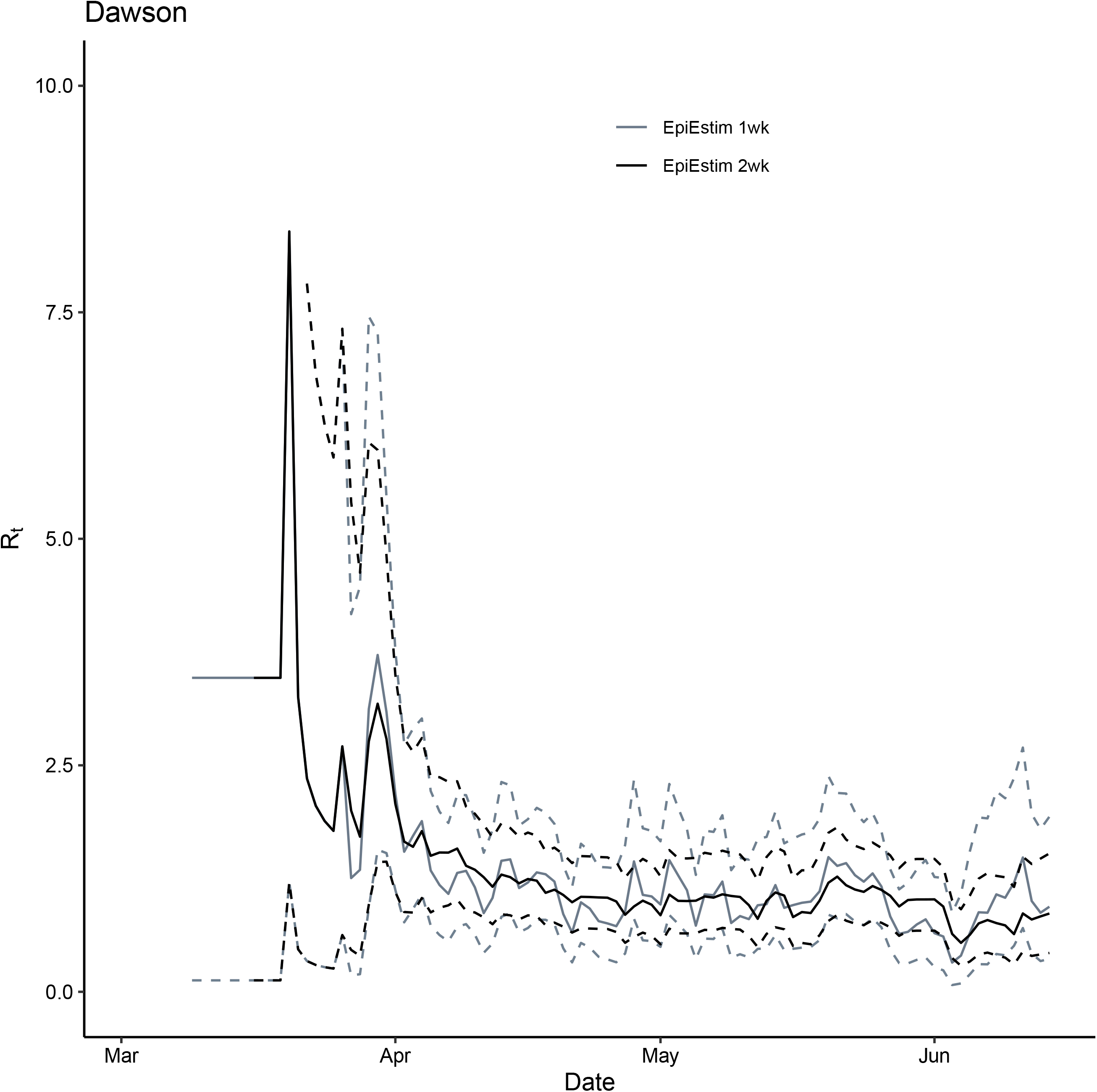
Comparison between *R*_*t*_ for Dawson County, GA, March 2—June 14, 2020, estimated using the instantaneous reproduction number method implemented in EpiEstim package (grey: 1-week window and black: 2-week window). Solid lines represent the median estimates and the dashed line represent the 2.5 and 97.5 quantiles.

**Figure S9.**
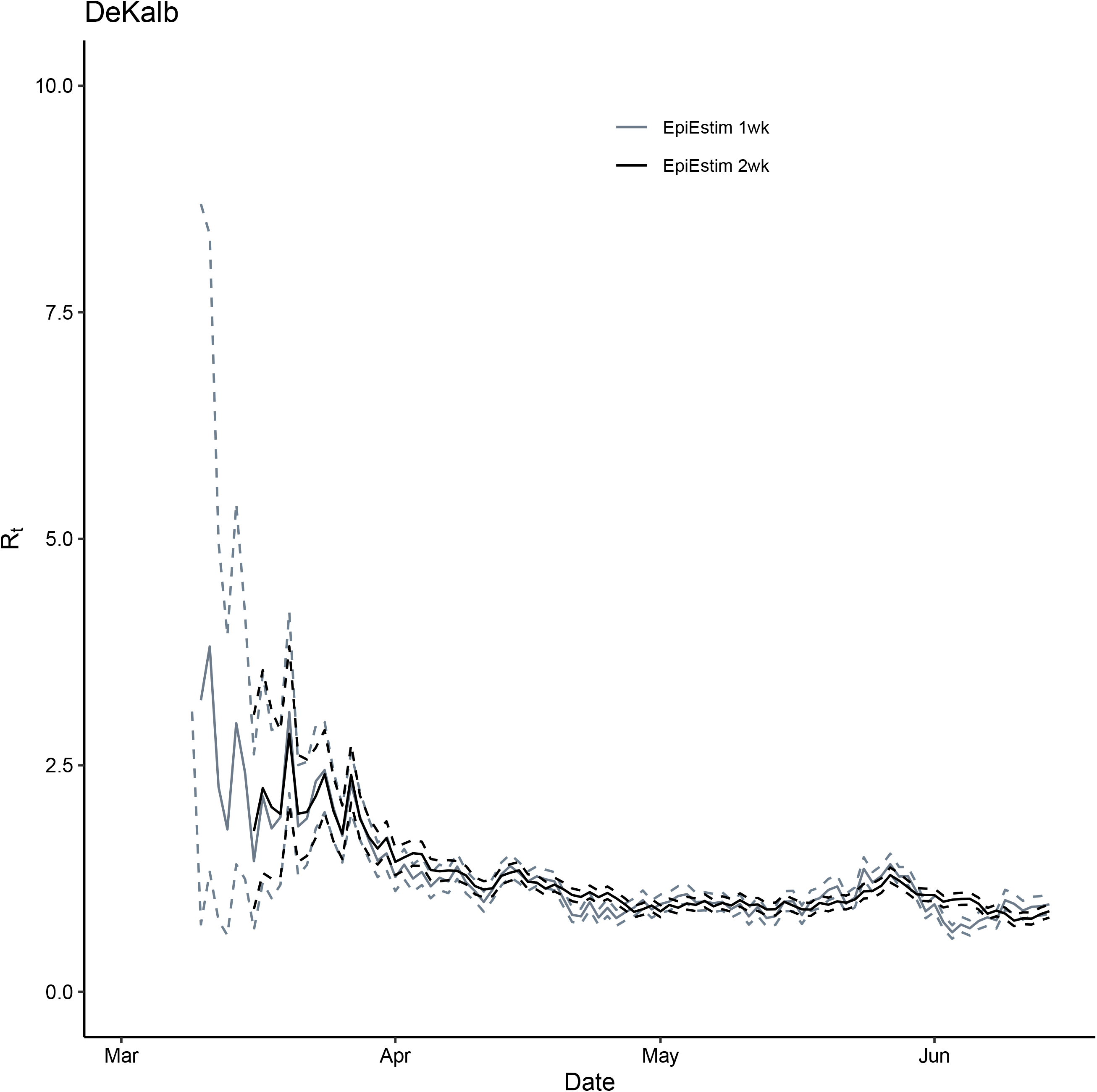
Comparison between *R*_*t*_ for DeKalb County, GA, March 2—June 14, 2020, estimated using the instantaneous reproduction number method implemented in EpiEstim package (grey: 1-week window and black: 2-week window). Solid lines represent the median estimates and the dashed line represent the 2.5 and 97.5 quantiles.

**Figure S10.**
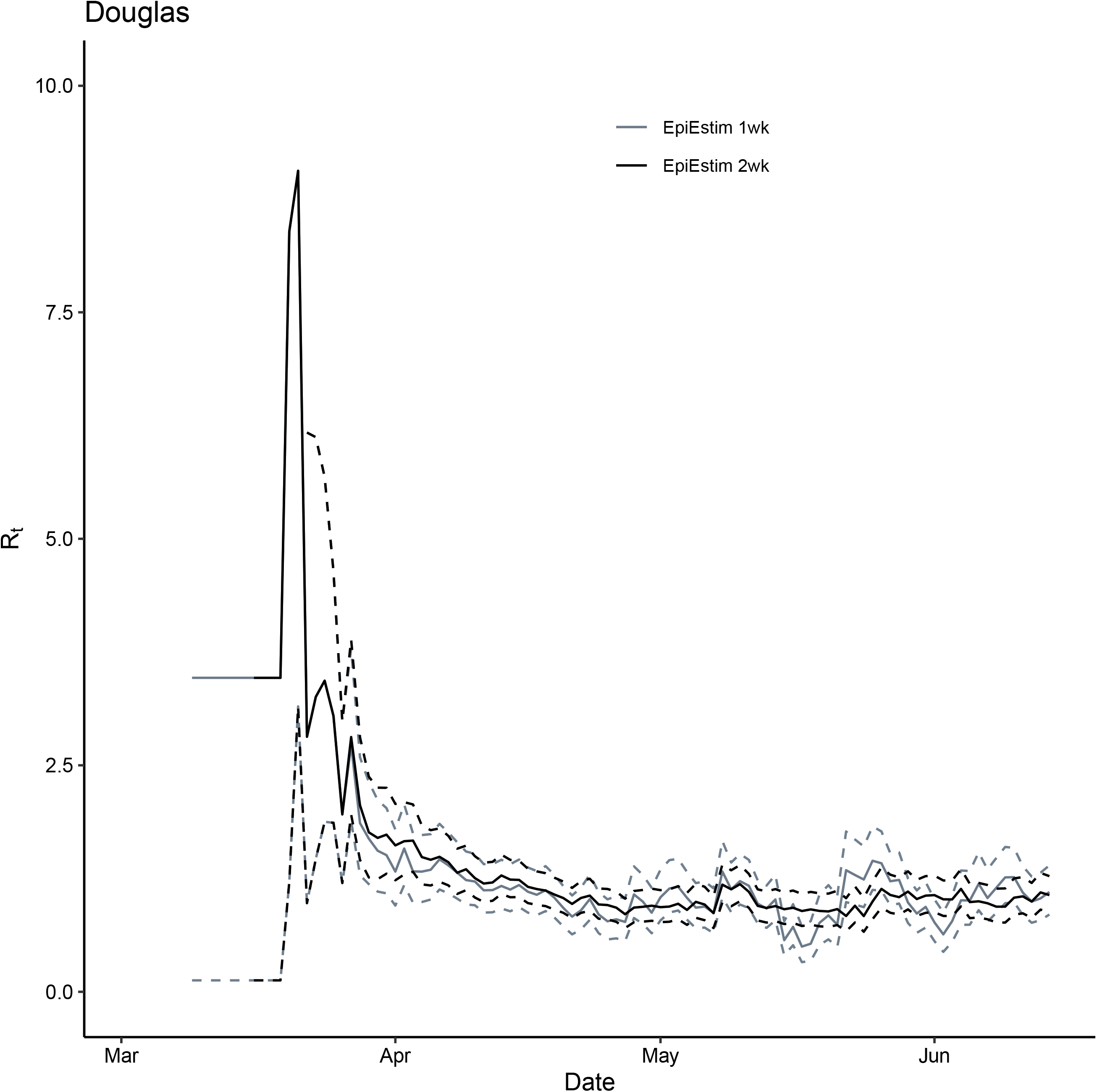
Comparison between *R*_*t*_ for Douglas County, GA, March 2—June 14, 2020, estimated using the instantaneous reproduction number method implemented in EpiEstim package (grey: 1-week window and black: 2-week window). Solid lines represent the median estimates and the dashed line represent the 2.5 and 97.5 quantiles.

**Figure S11.**
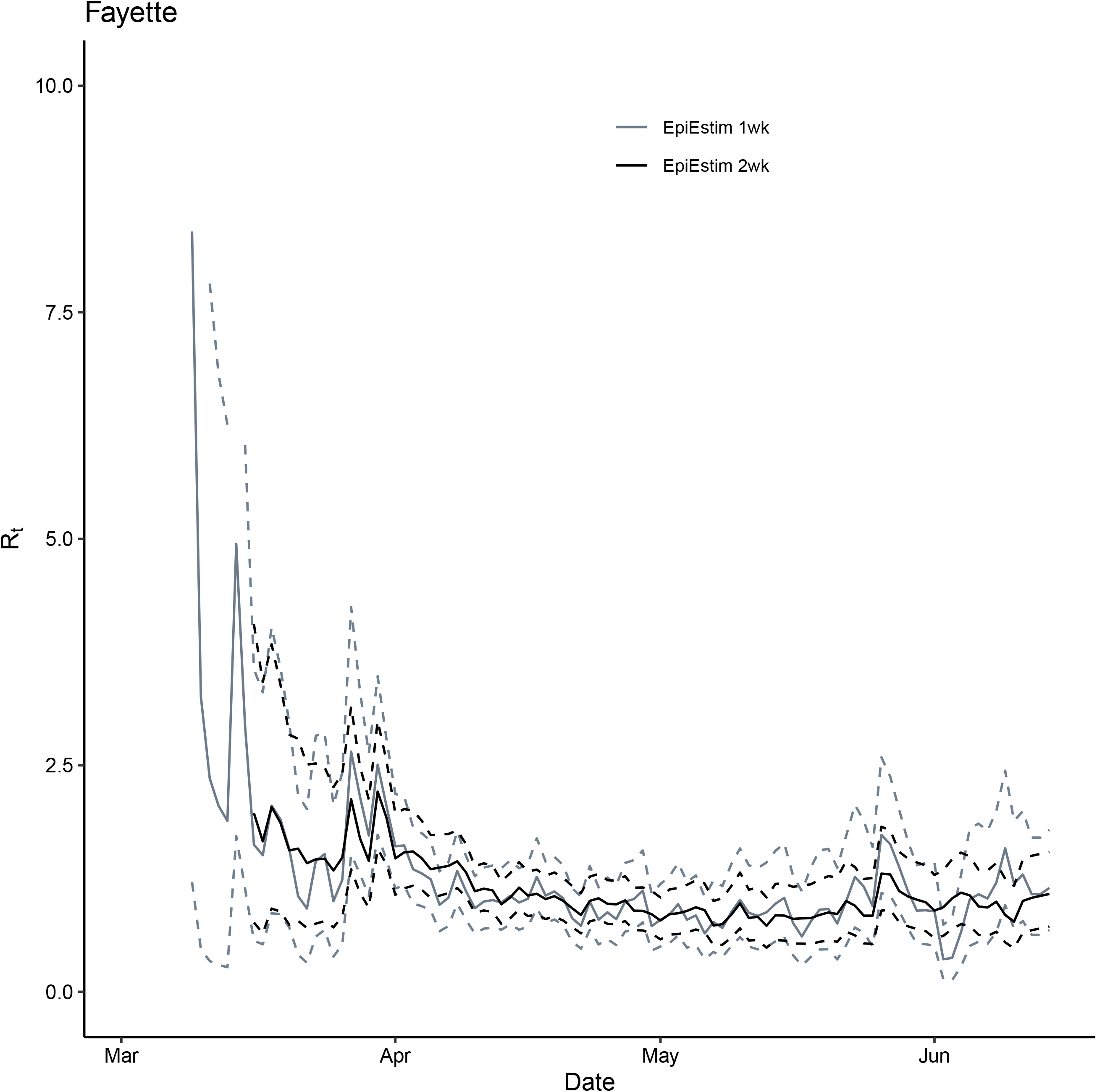
Comparison between *R*_*t*_ for Fayette County, GA, March 2—June 14, 2020, estimated using the instantaneous reproduction number method implemented in EpiEstim package (grey: 1-week window and black: 2-week window). Solid lines represent the median estimates and the dashed line represent the 2.5 and 97.5 quantiles.

**Figure S12.**
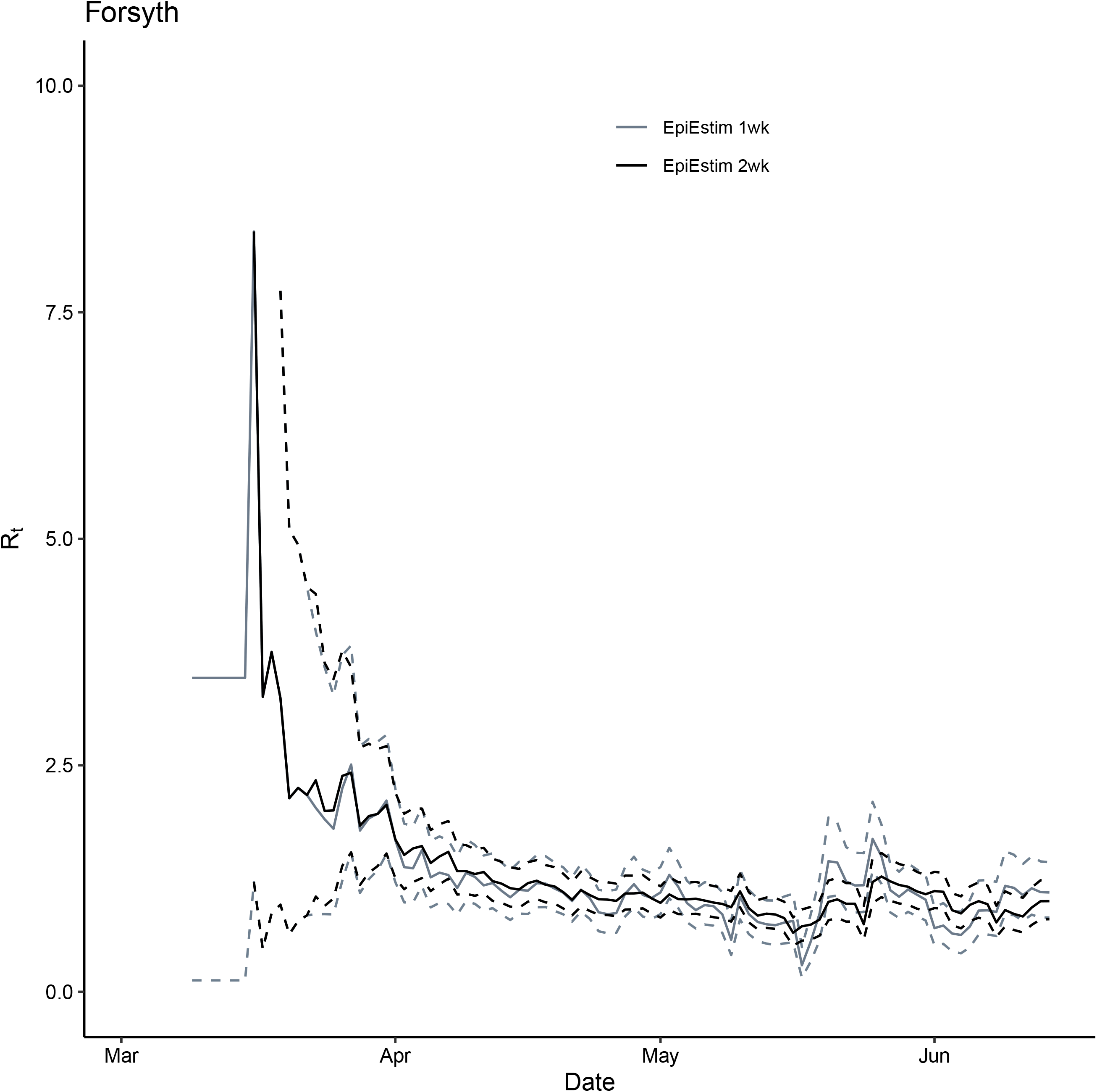
Comparison between *R*_*t*_ for Forsyth County, GA, March 2—June 14, 2020, estimated using the instantaneous reproduction number method implemented in EpiEstim package (grey: 1-week window and black: 2-week window). Solid lines represent the median estimates and the dashed line represent the 2.5 and 97.5 quantiles.

**Figure S13.**
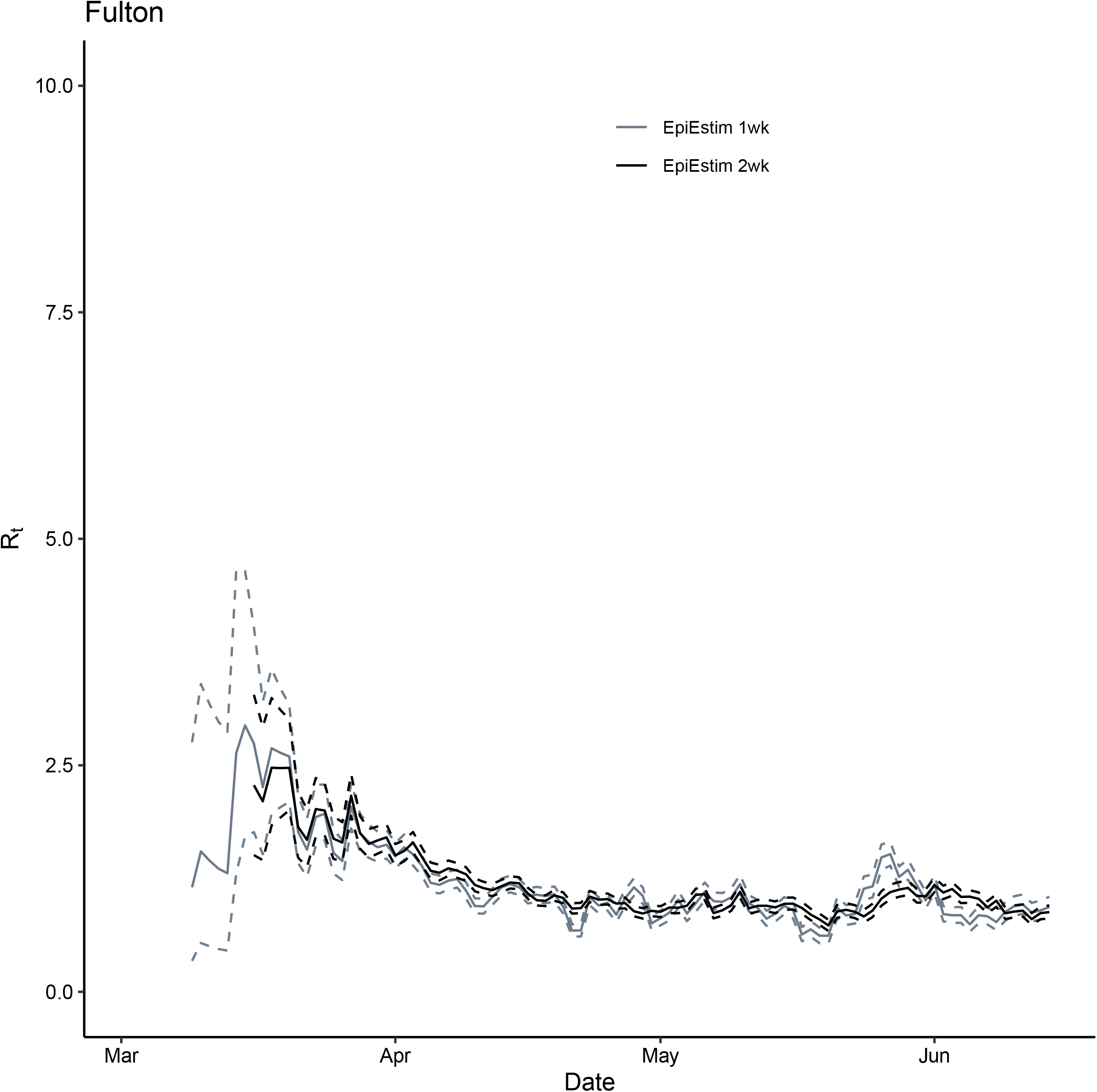
Comparison between *R*_*t*_ for Fulton County, GA, March 2—June 14, 2020, estimated using the instantaneous reproduction number method implemented in EpiEstim package (grey: 1-week window and black: 2-week window). Solid lines represent the median estimates and the dashed line represent the 2.5 and 97.5 quantiles.

**Figure S14.**
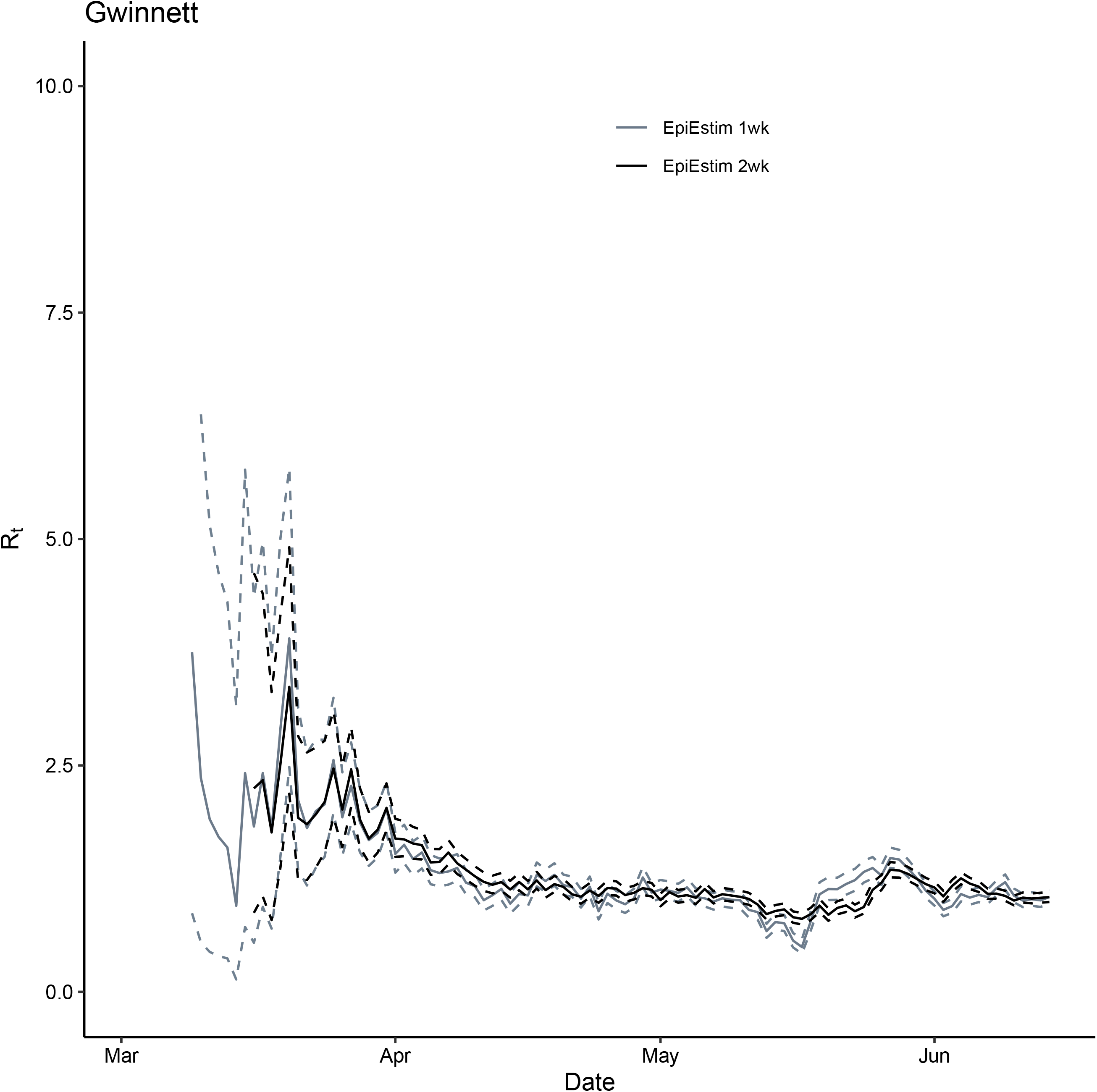
Comparison between *R*_*t*_ for Gwinnett County, GA, March 2—June 14, 2020, estimated using the instantaneous reproduction number method implemented in EpiEstim package (grey: 1-week window and black: 2-week window). Solid lines represent the median estimates and the dashed line represent the 2.5 and 97.5 quantiles.

**Figure S15.**
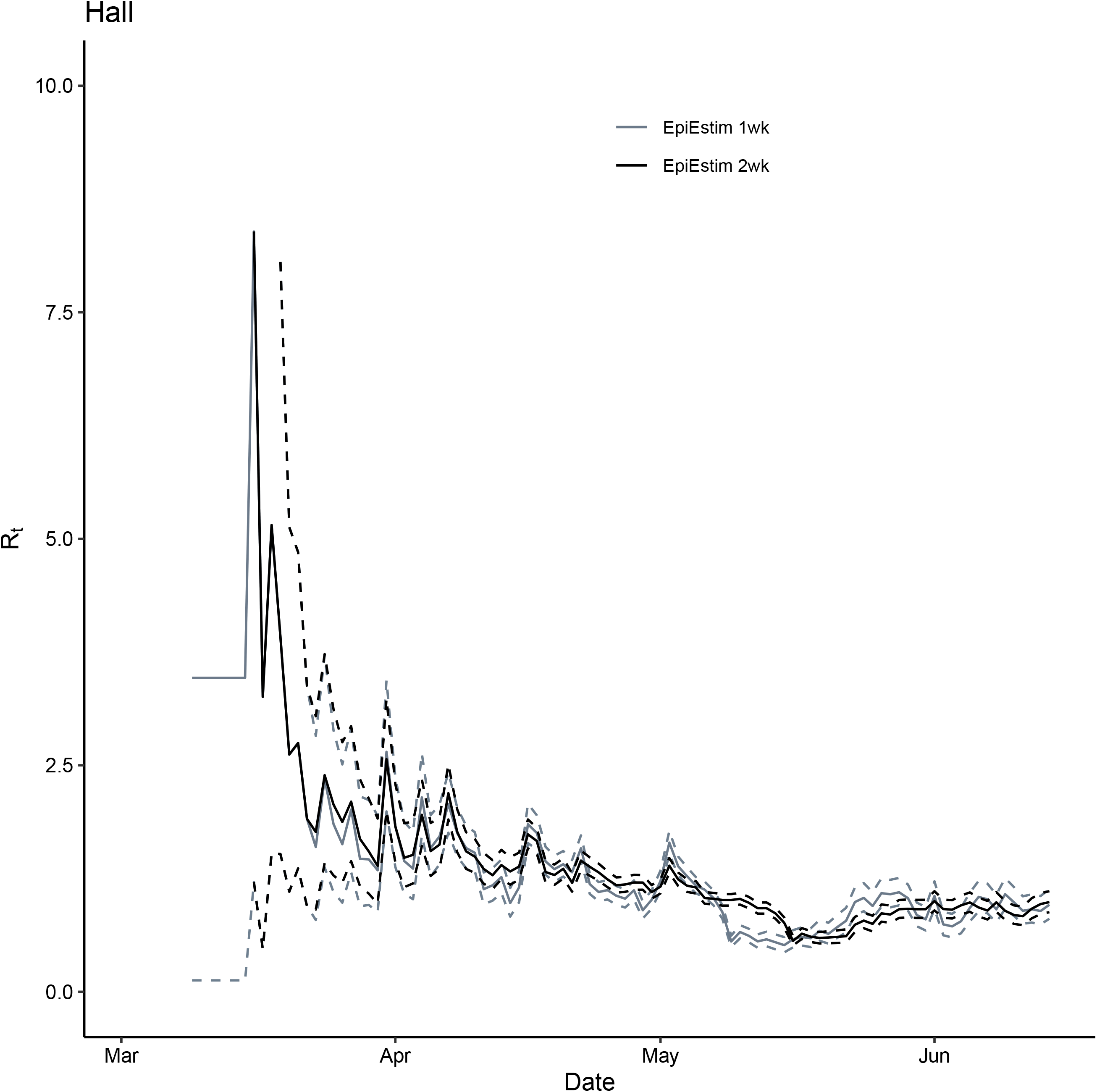
Comparison between *R*_*t*_ for Hall County, GA, March 2—June 14, 2020, estimated using the instantaneous reproduction number method implemented in EpiEstim package (grey: 1-week window and black: 2-week window). Solid lines represent the median estimates and the dashed line represent the 2.5 and 97.5 quantiles.

**Figure S16.**
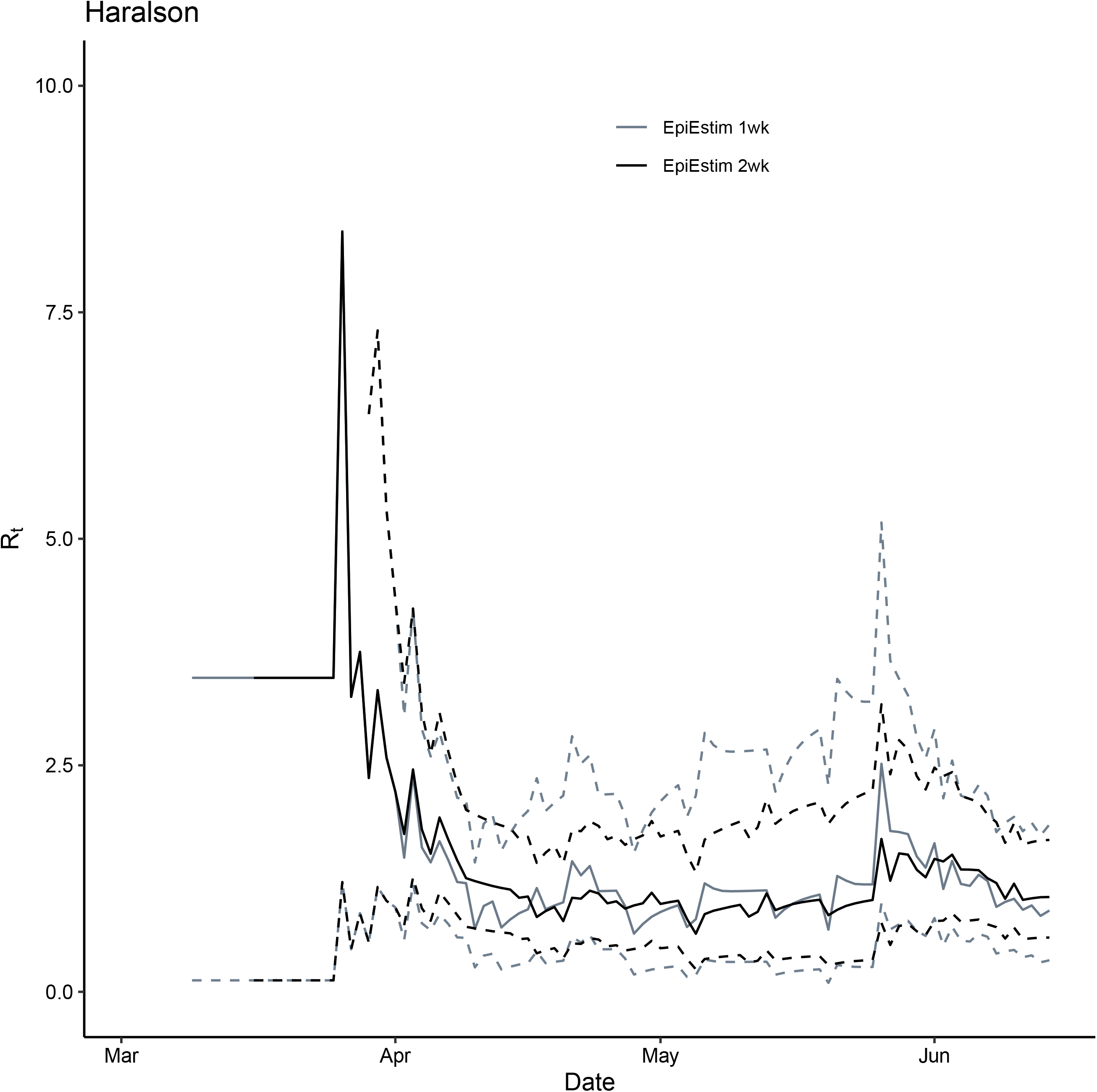
Comparison between *R*_*t*_ for Haralson County, GA, March 2—June 14, 2020, estimated using the instantaneous reproduction number method implemented in EpiEstim package (grey: 1-week window and black: 2-week window). Solid lines represent the median estimates and the dashed line represent the 2.5 and 97.5 quantiles.

**Figure S17.**
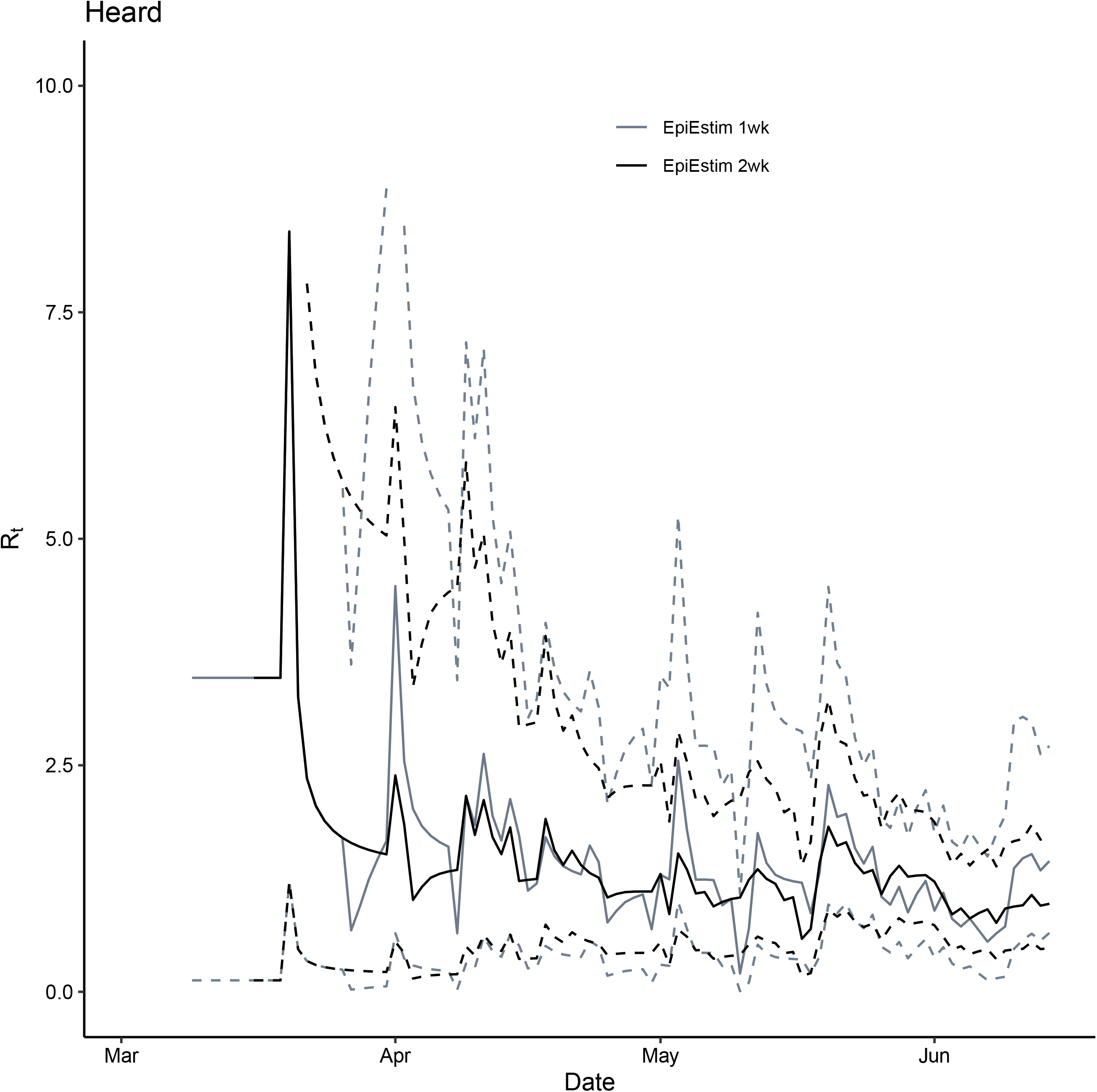
Comparison between *R*_*t*_ for Heard County, GA, March 2—June 14, 2020, estimated using the instantaneous reproduction number method implemented in EpiEstim package (grey: 1-week window and black: 2-week window). Solid lines represent the median estimates and the dashed line represent the 2.5 and 97.5 quantiles.

**Figure S18.**
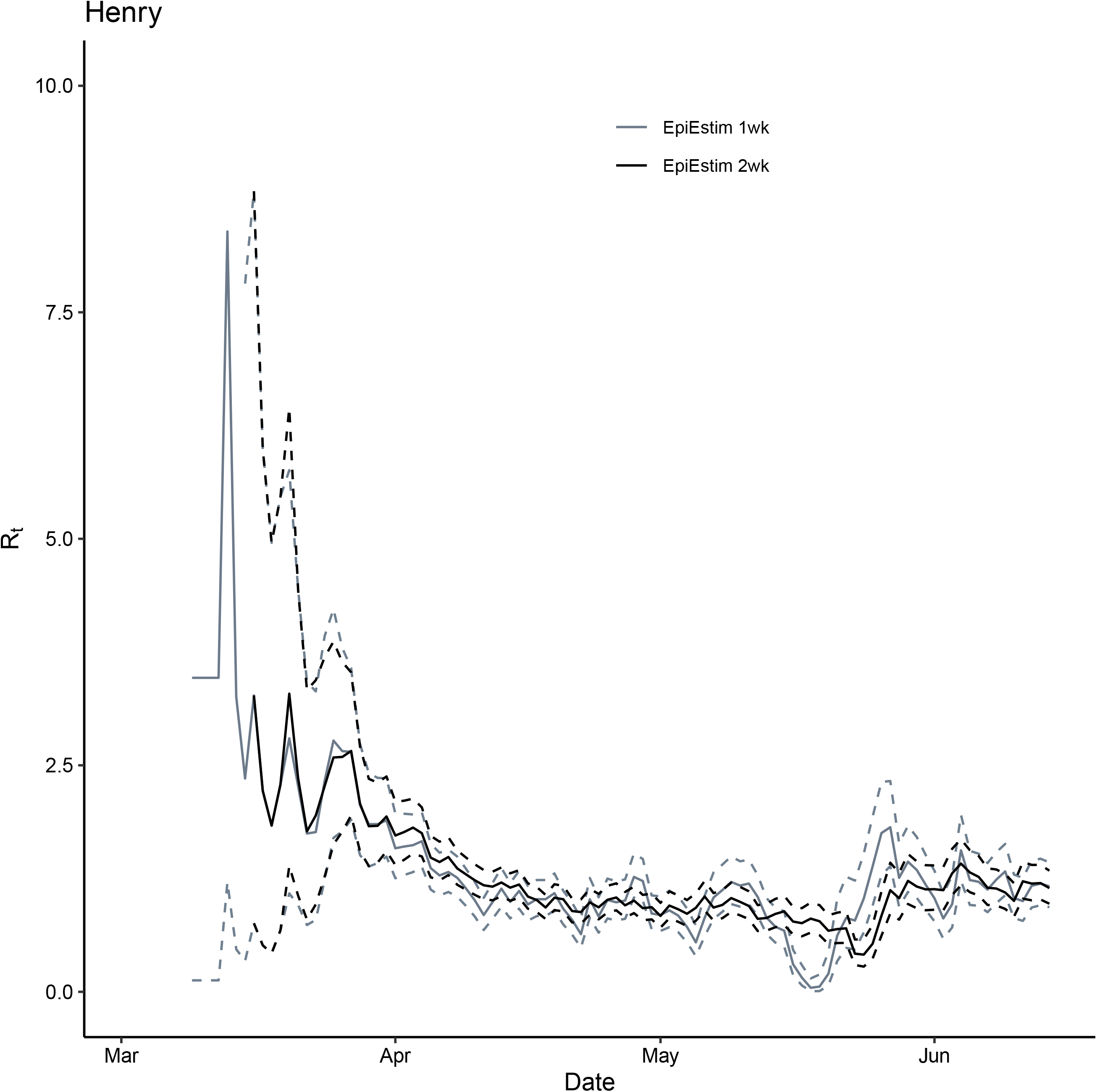
Comparison between *R*_*t*_ for Henry County, GA, March 2—June 14, 2020, estimated using the instantaneous reproduction number method implemented in EpiEstim package (grey: 1-week window and black: 2-week window). Solid lines represent the median estimates and the dashed line represent the 2.5 and 97.5 quantiles.

**Figure S19.**
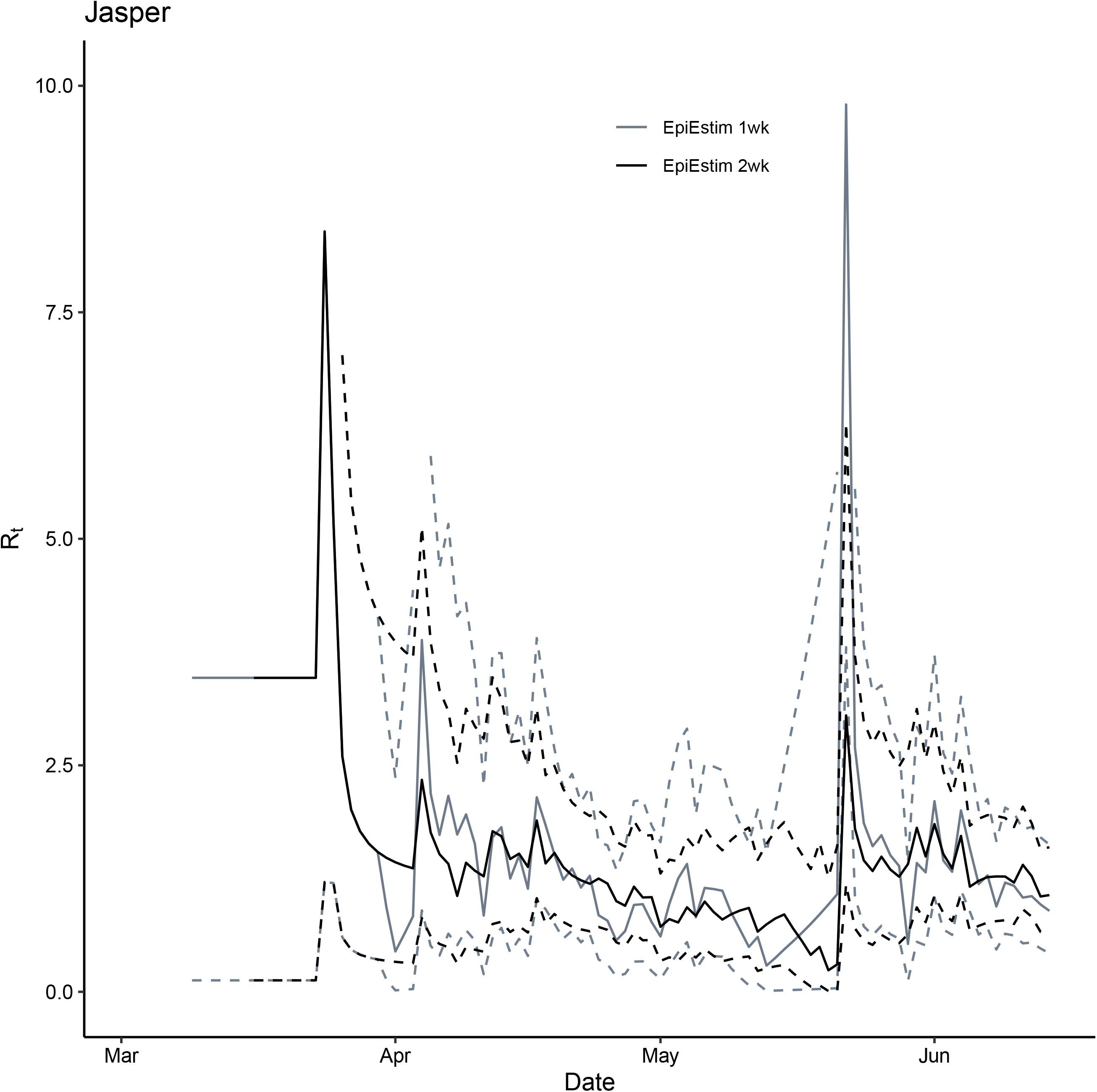
Comparison between *R*_*t*_ for Jasper County, GA, March 2—June 14, 2020, estimated using the instantaneous reproduction number method implemented in EpiEstim package (grey: 1-week window and black: 2-week window). Solid lines represent the median estimates and the dashed line represent the 2.5 and 97.5 quantiles.

**Figure S20.**
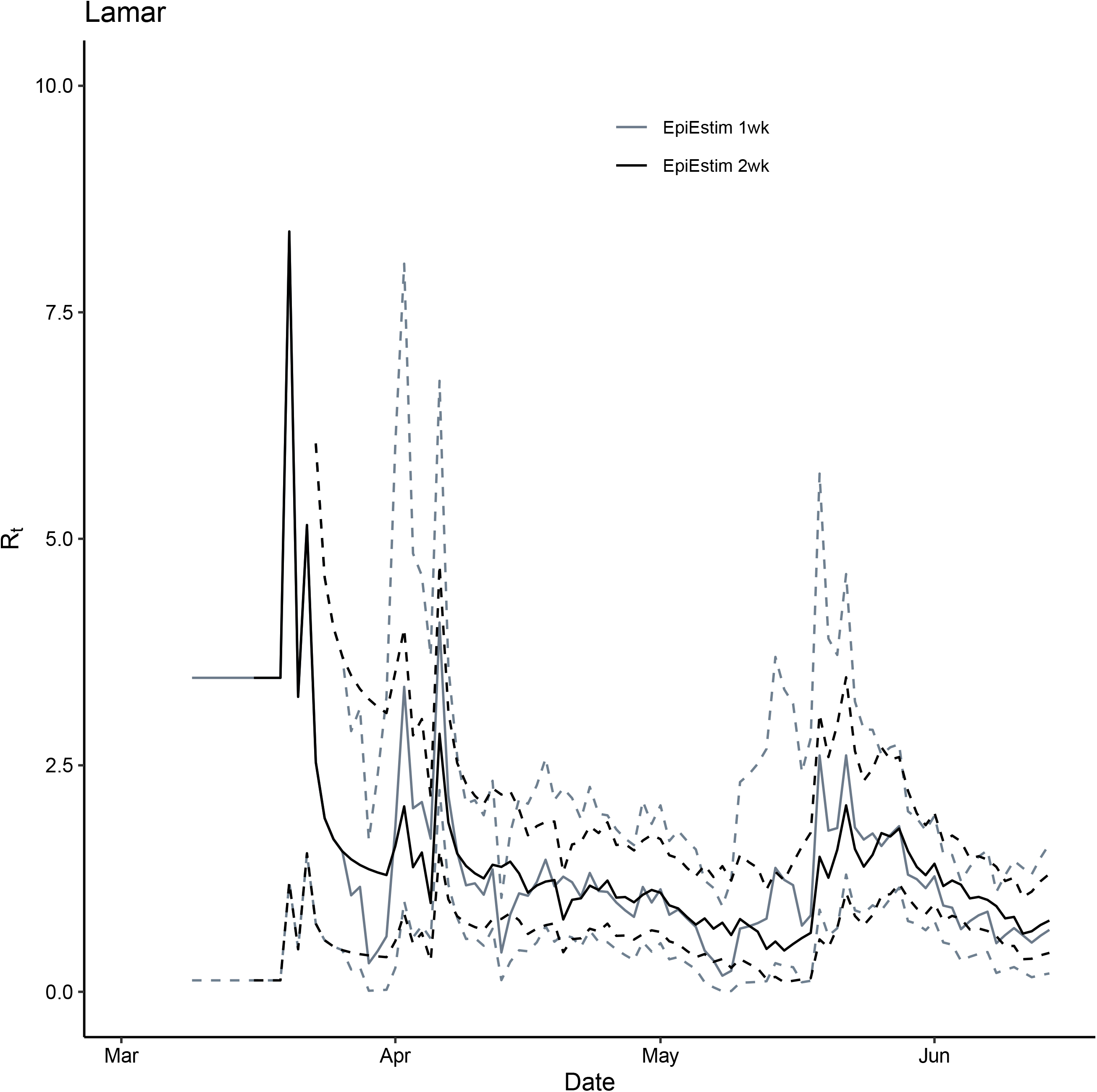
Comparison between *R*_*t*_ for Lamar County, GA, March 2—June 14, 2020, estimated using the instantaneous reproduction number method implemented in EpiEstim package (grey: 1-week window and black: 2-week window). Solid lines represent the median estimates and the dashed line represent the 2.5 and 97.5 quantiles.

**Figure S21.**
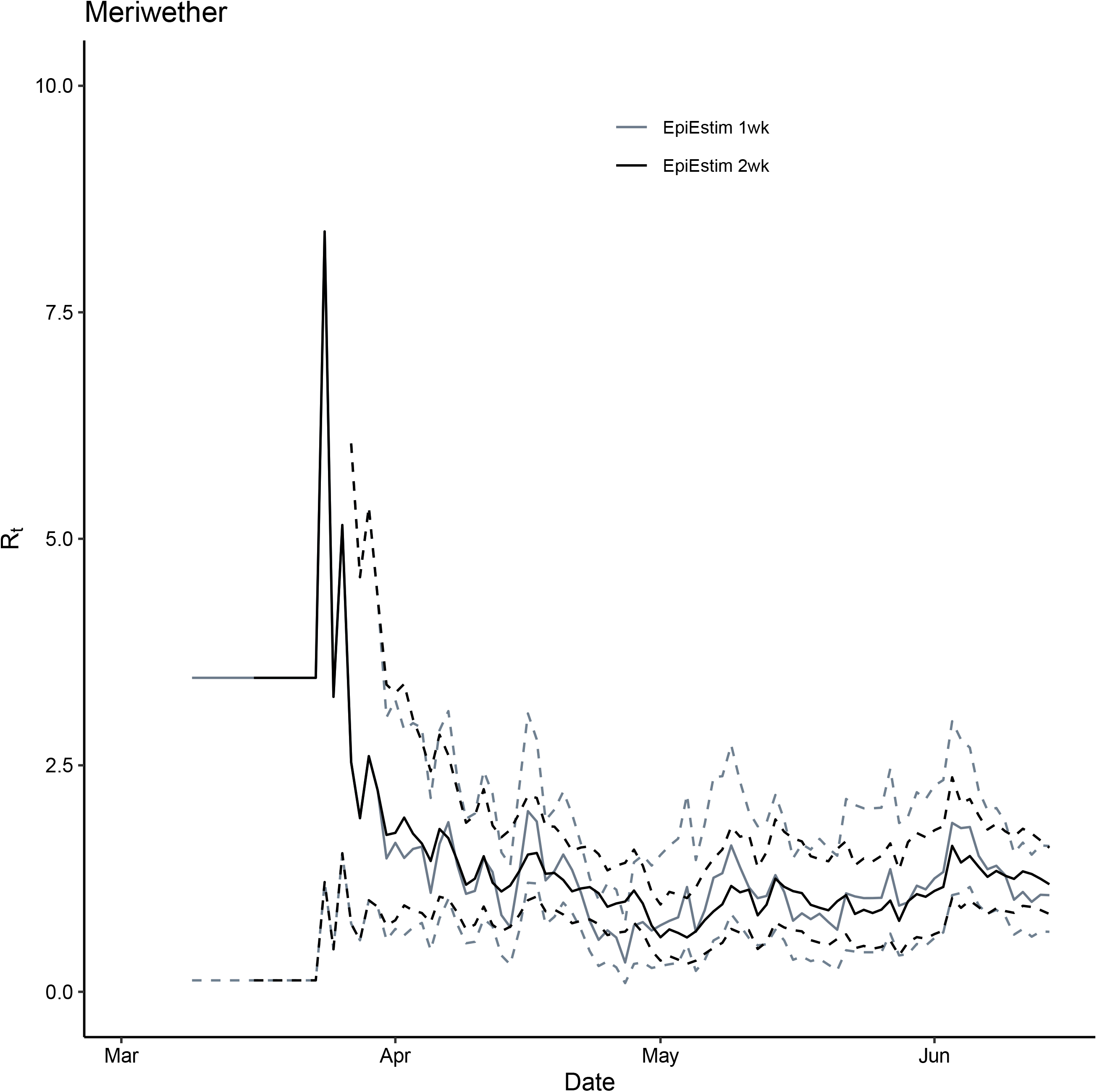
Comparison between *R*_*t*_ for Meriwether County, GA, March 2—June 14, 2020, estimated using the instantaneous reproduction number method implemented in EpiEstim package (grey: 1-week window and black: 2-week window). Solid lines represent the median estimates and the dashed line represent the 2.5 and 97.5 quantiles.

**Figure S22.**
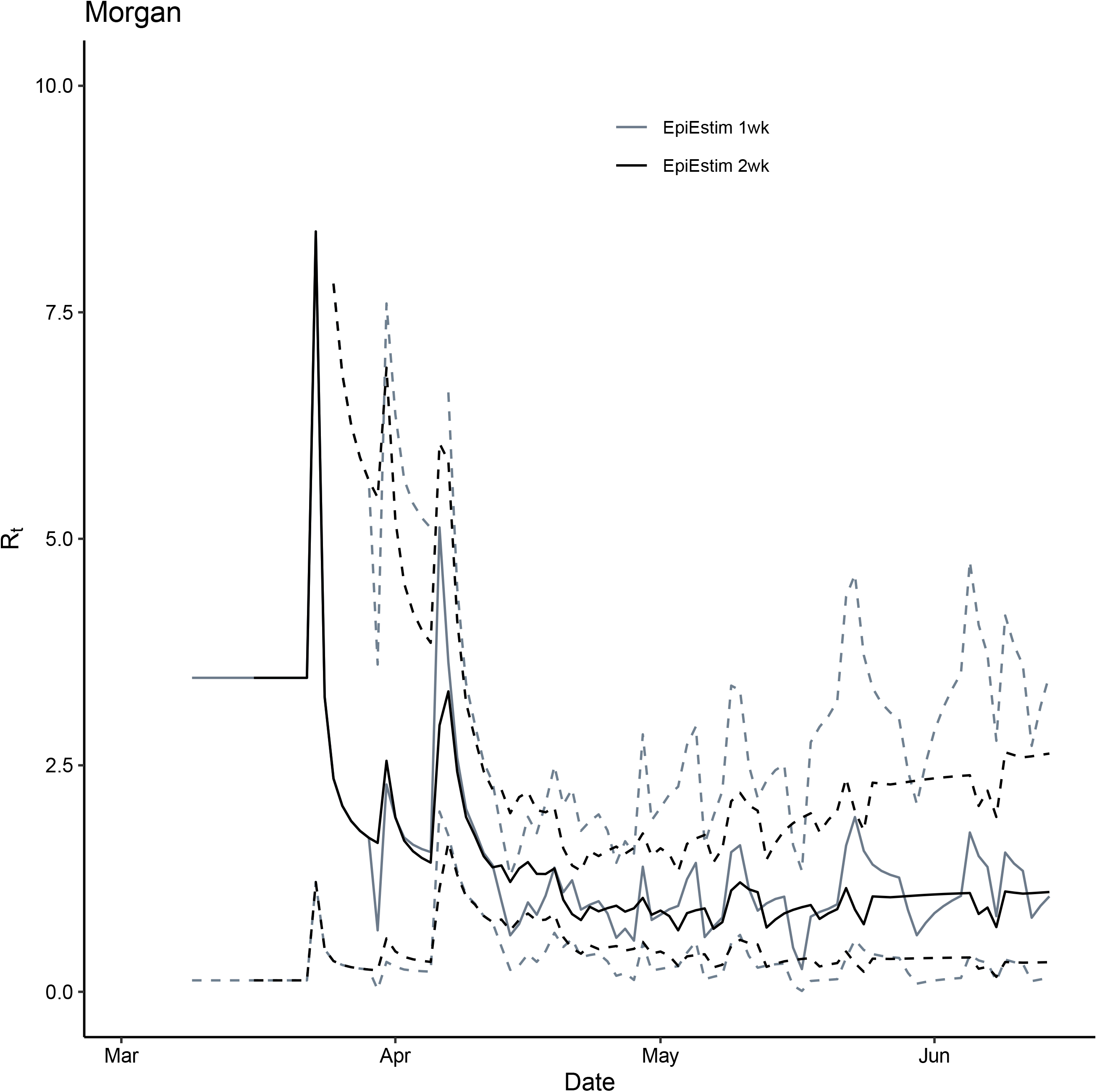
Comparison between *R*_*t*_ for Morgan County, GA, March 2—June 14, 2020, estimated using the instantaneous reproduction number method implemented in EpiEstim package (grey: 1-week window and black: 2-week window). Solid lines represent the median estimates and the dashed line represent the 2.5 and 97.5 quantiles.

**Figure S23.**
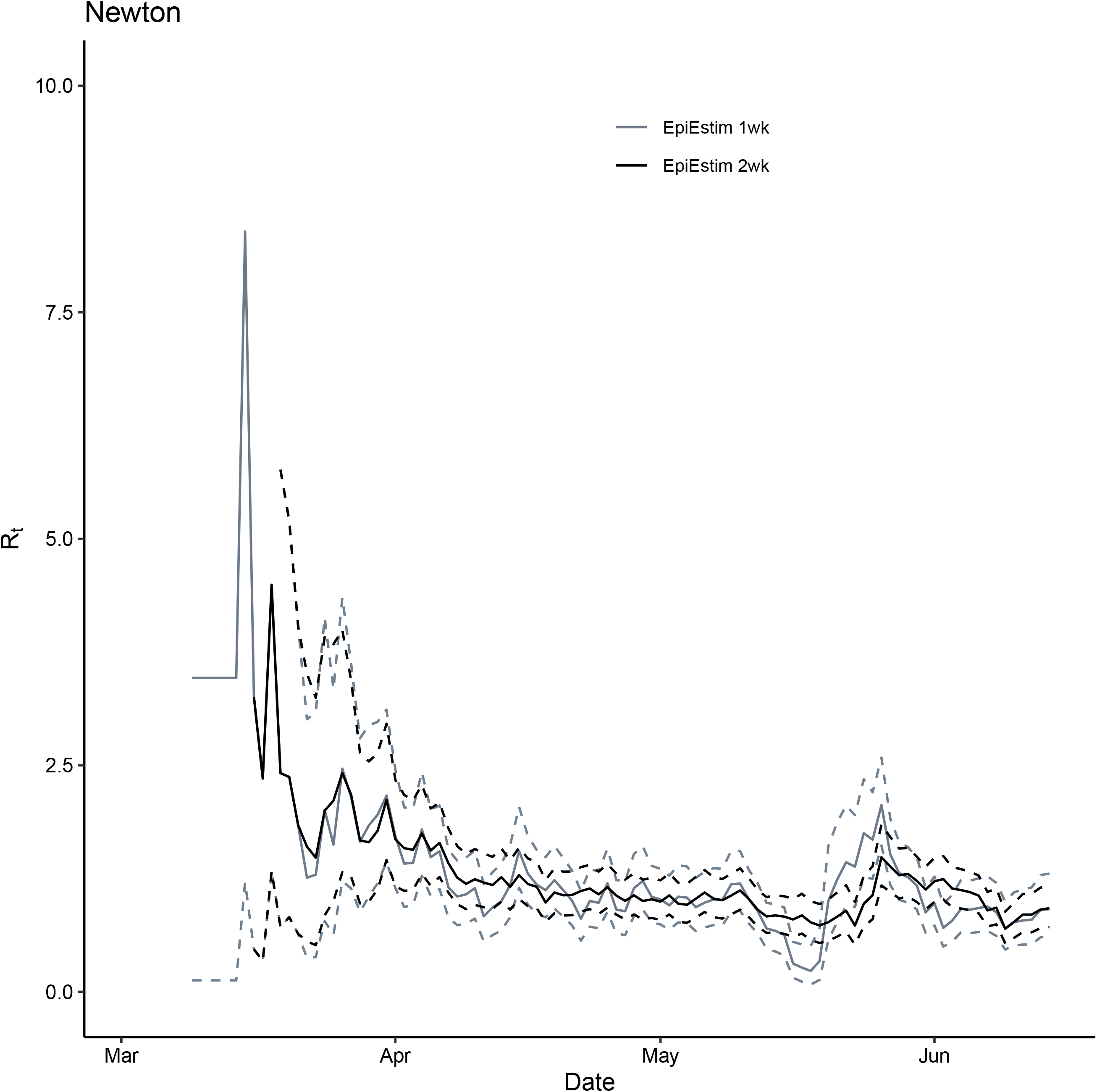
Comparison between *R*_*t*_ for Newton County, GA, March 2—June 14, 2020, estimated using the instantaneous reproduction number method implemented in EpiEstim package (grey: 1-week window and black: 2-week window). Solid lines represent the median estimates and the dashed line represent the 2.5 and 97.5 quantiles.

**Figure S24.**
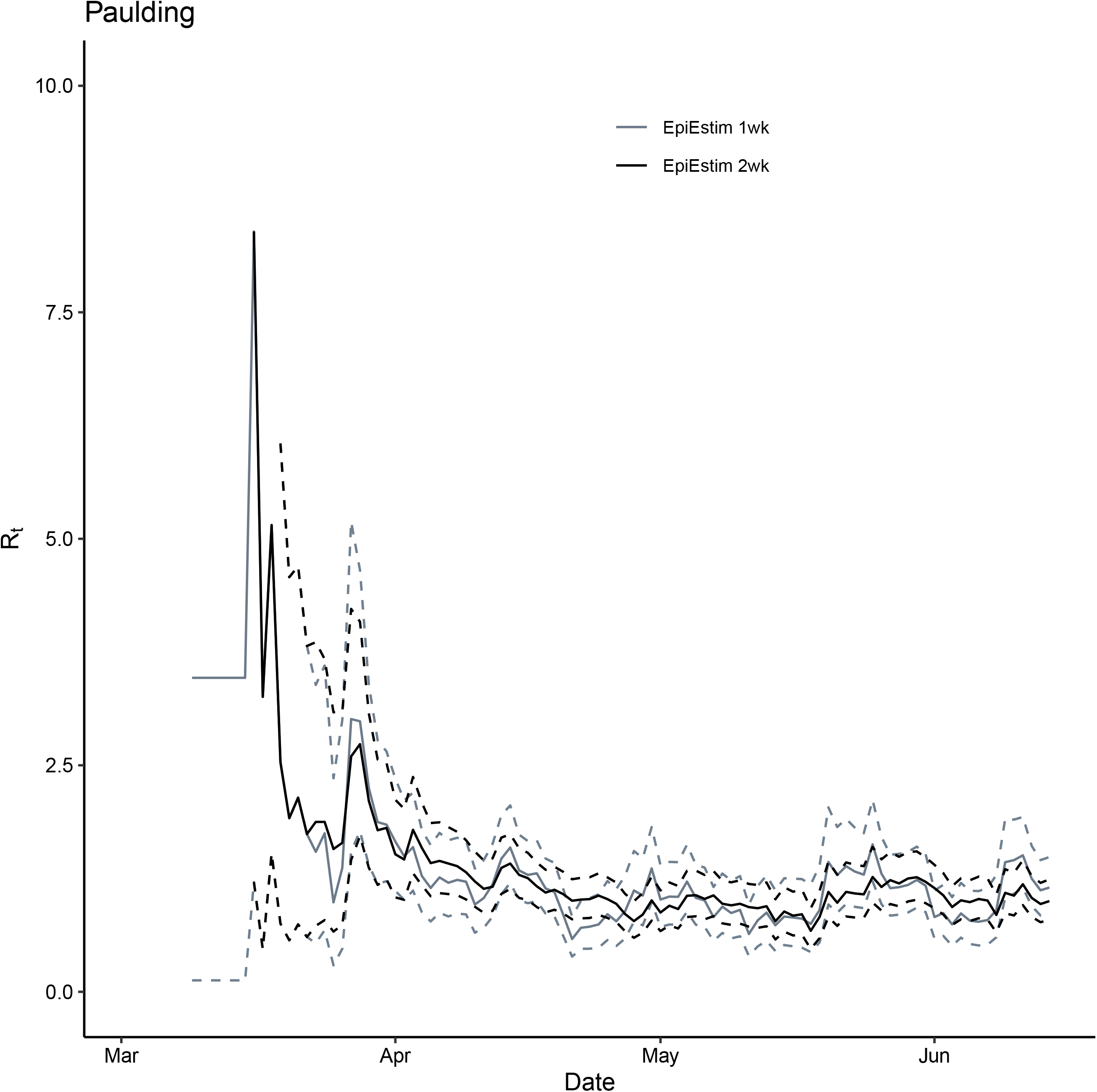
Comparison between *R*_*t*_ for Paulding County, GA, March 2—June 14, 2020, estimated using the instantaneous reproduction number method implemented in EpiEstim package (grey: 1-week window and black: 2-week window). Solid lines represent the median estimates and the dashed line represent the 2.5 and 97.5 quantiles.

**Figure S25.**
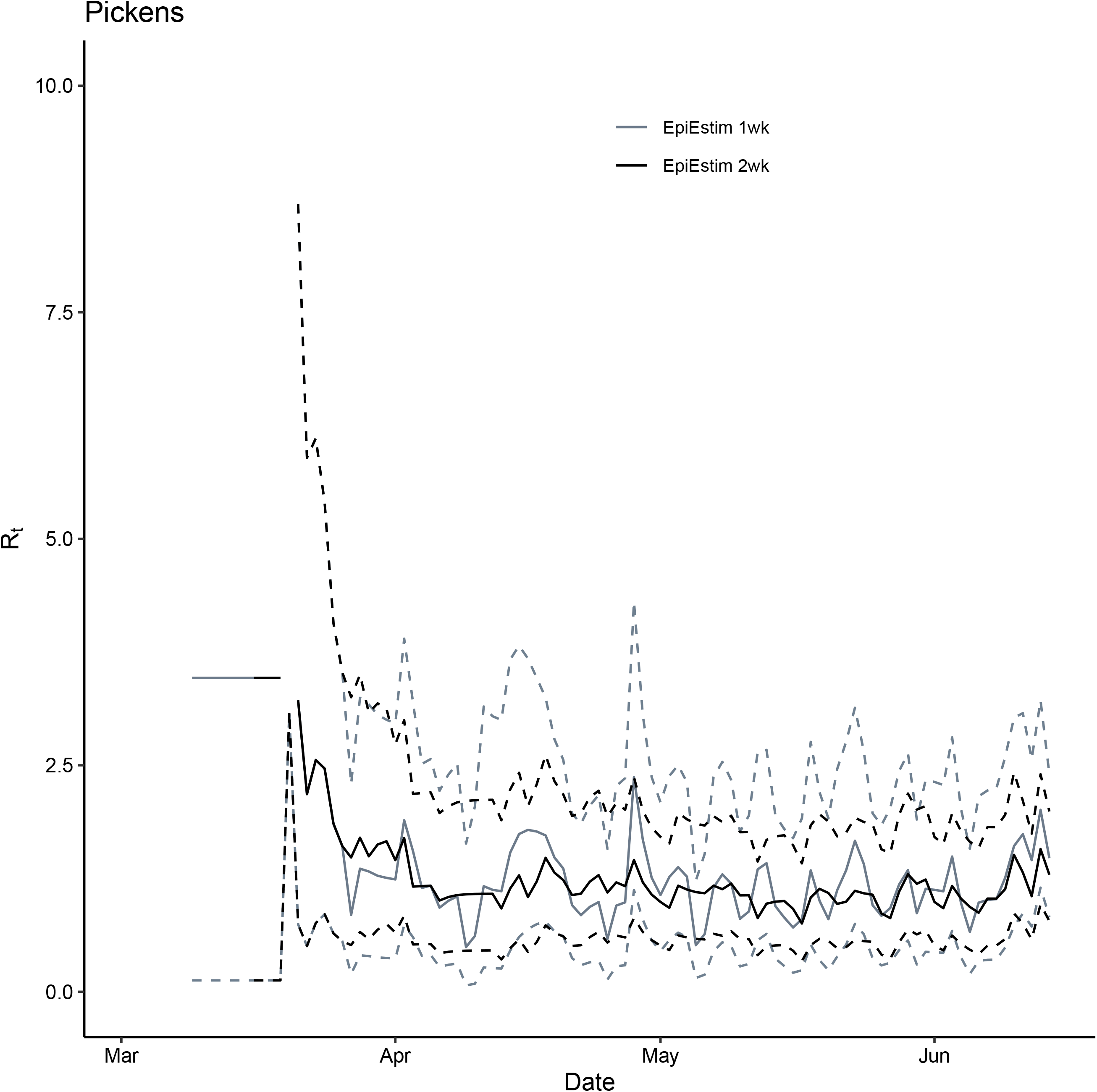
Comparison between *R*_*t*_ for Pickens County, GA, March 2—June 14, 2020, estimated using the instantaneous reproduction number method implemented in EpiEstim package (grey: 1-week window and black: 2-week window). Solid lines represent the median estimates and the dashed line represent the 2.5 and 97.5 quantiles.

**Figure S26.**
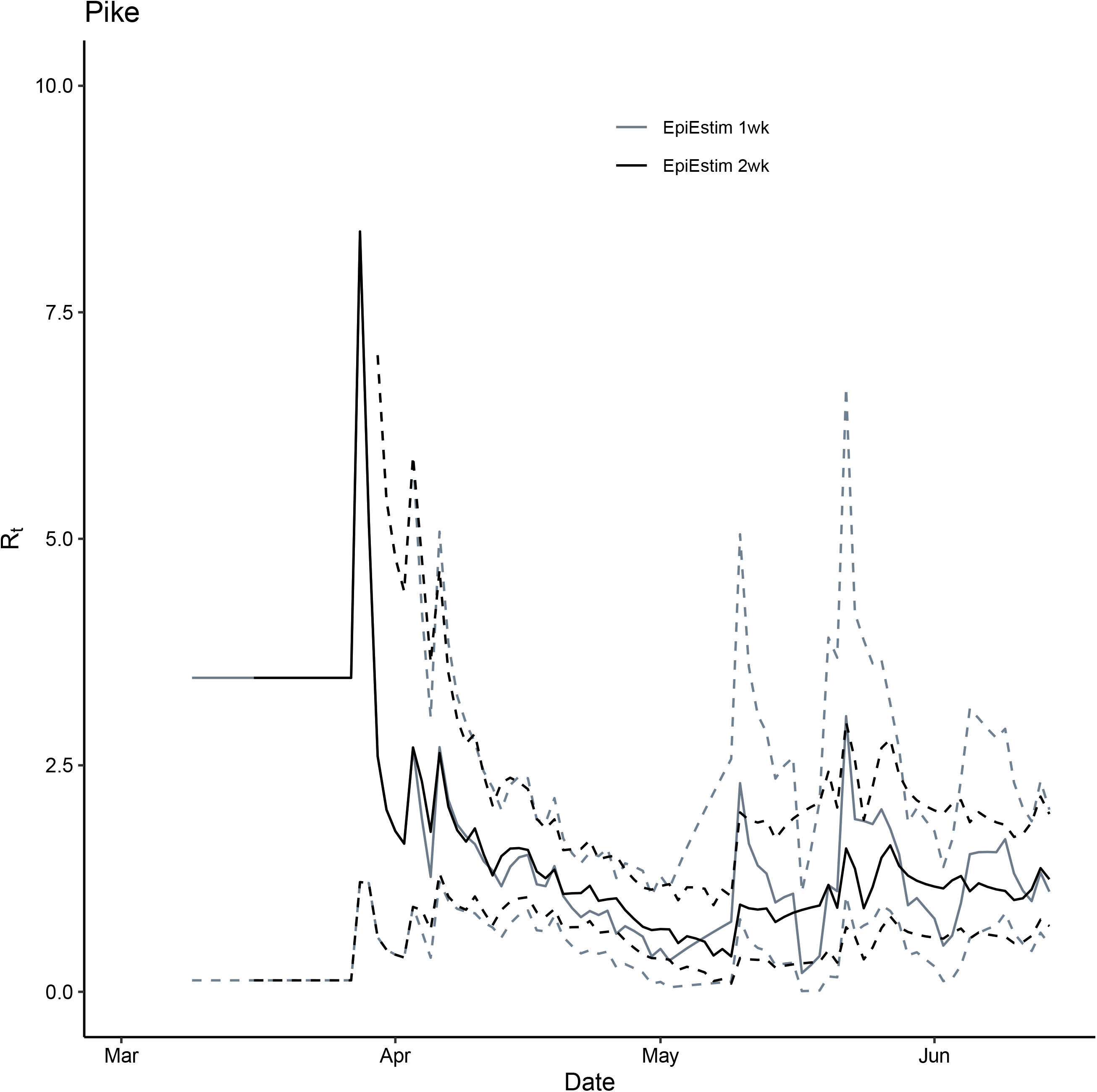
Comparison between *R*_*t*_ for Pike County, GA, March 2—June 14, 2020, estimated using the instantaneous reproduction number method implemented in EpiEstim package (grey: 1-week window and black: 2-week window). Solid lines represent the median estimates and the dashed line represent the 2.5 and 97.5 quantiles.

**Figure S27.**
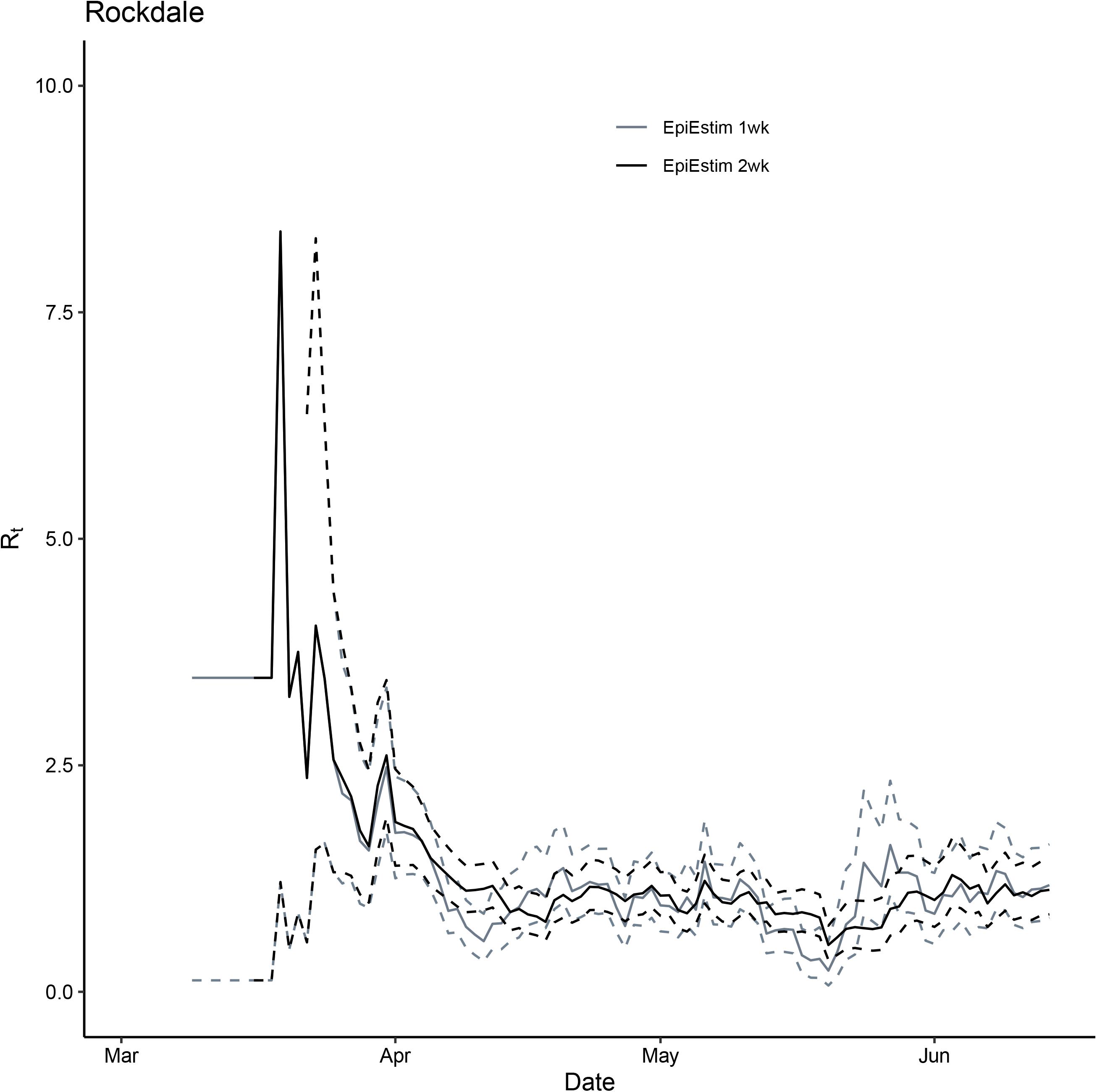
Comparison between *R*_*t*_ for Rockdale County, GA, March 2—June 14, 2020, estimated using the instantaneous reproduction number method implemented in EpiEstim package (grey: 1-week window and black: 2-week window). Solid lines represent the median estimates and the dashed line represent the 2.5 and 97.5 quantiles.

**Figure S28.**
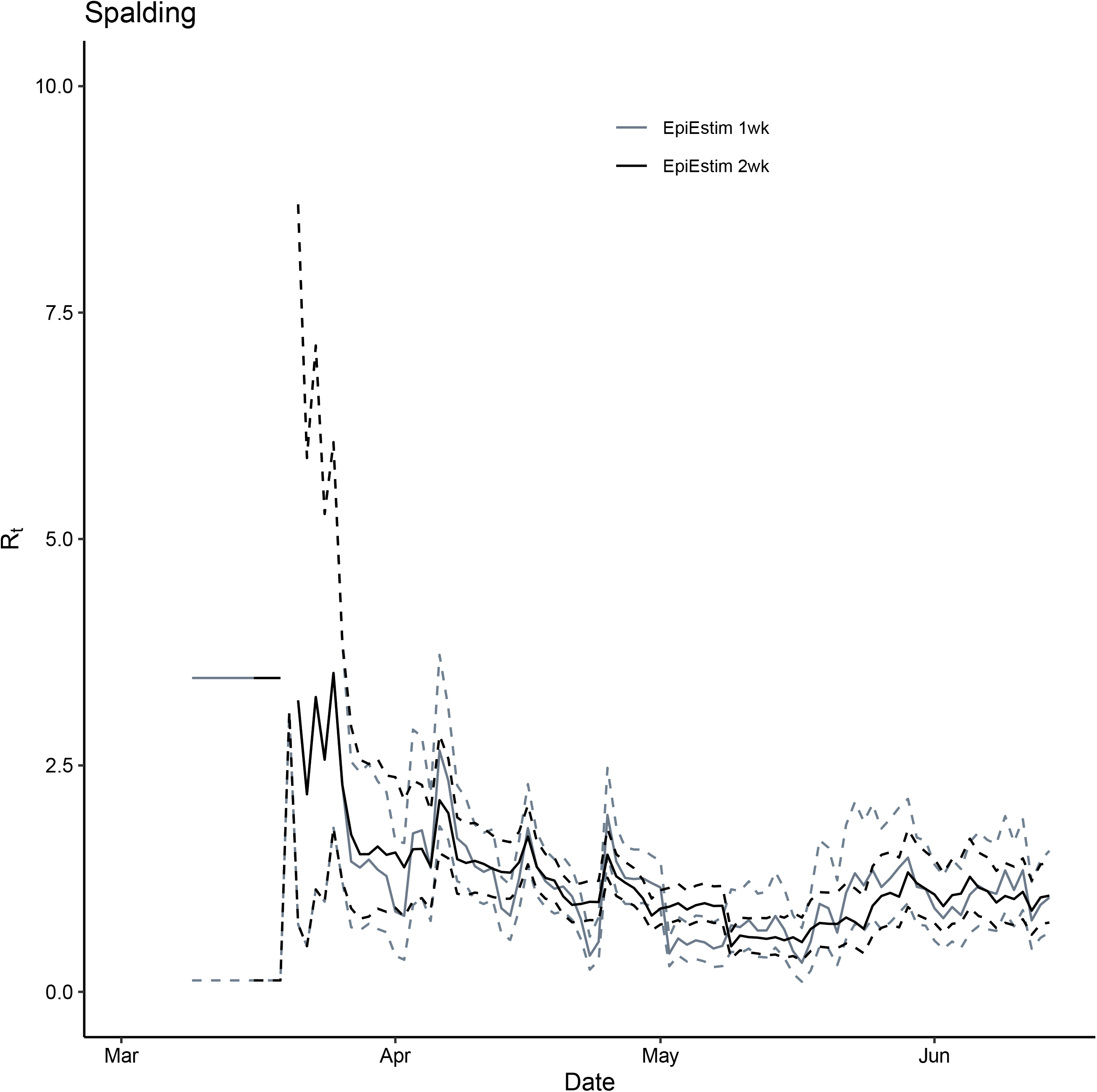
Comparison between *R*_*t*_ for Spalding County, GA, March 2—June 14, 2020, estimated using the instantaneous reproduction number method implemented in EpiEstim package (grey: 1-week window and black: 2-week window). Solid lines represent the median estimates and the dashed line represent the 2.5 and 97.5 quantiles.

**Figure S29.**
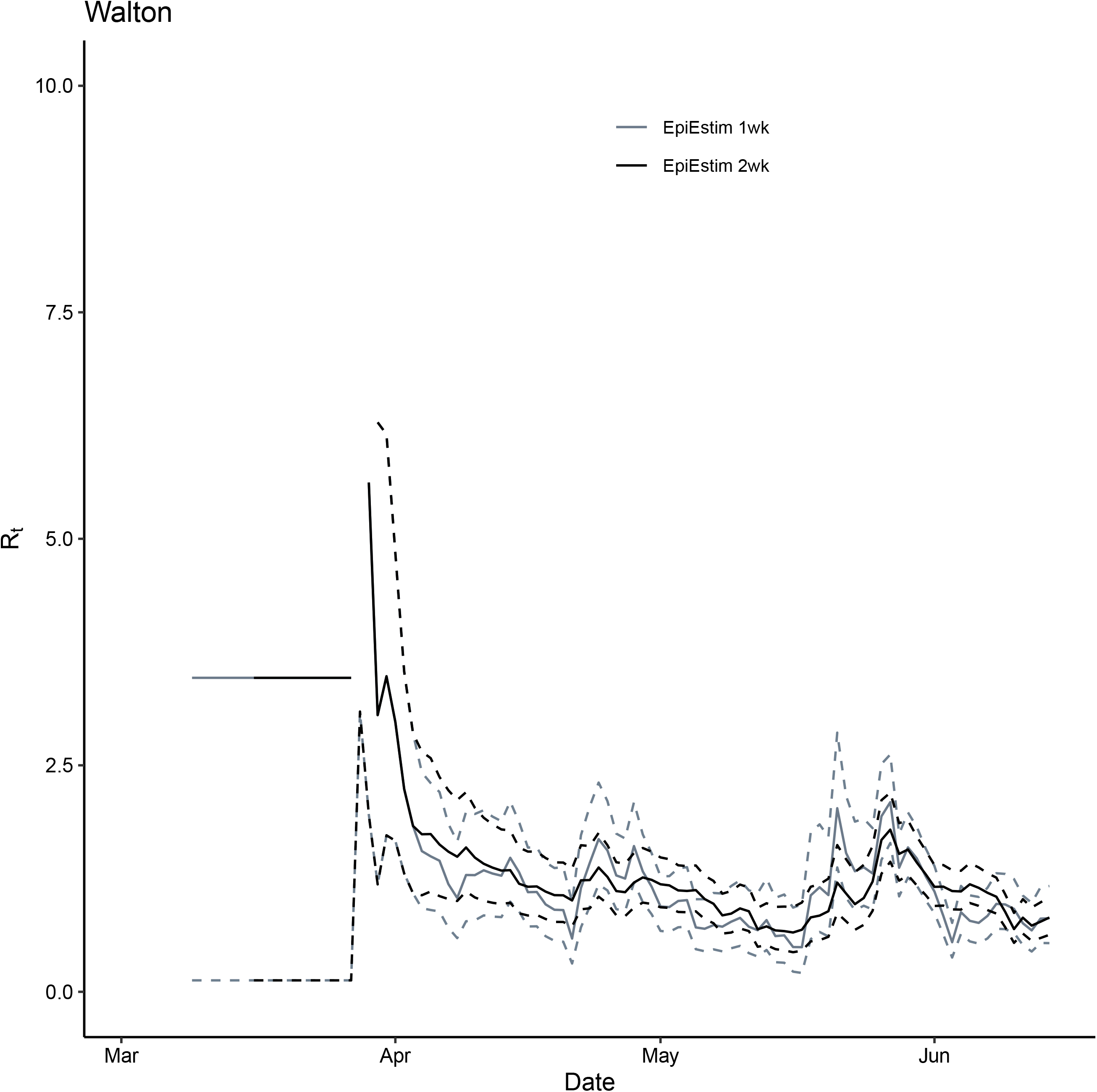
Comparison between *R*_*t*_ for Walton County, GA, March 2—June 14, 2020, estimated using the instantaneous reproduction number method implemented in EpiEstim package (grey: 1-week window and black: 2-week window). Solid lines represent the median estimates and the dashed line represent the 2.5 and 97.5 quantiles.

## Notes

### Competing Interest Statement

The authors have declared no competing interest.

### Funding Statement

GC acknowledges support from NSF grant 1414374 as part of the joint NSF-NIH-USDA Ecology and Evolution of Infectious Diseases program. ICHF acknowledges salary support from the National Center for Emerging and Zoonotic Infectious Diseases, Centers for Disease Control and Prevention (19IPA1908208). This article is not part of ICHF's CDC-sponsored projects.

### Author Declarations

The Georgia Southern University Institutional Review Board made a nonhuman subjects determination for this project (H20364), under the G8 exemption category.

